# Results reporting for clinical trials led by medical universities and university hospitals in the Nordic countries was often missing or delayed

**DOI:** 10.1101/2024.02.04.24301363

**Authors:** Gustav Nilsonne, Susanne Wieschowski, Nicholas J. DeVito, Maia Salholz-Hillel, Love Ahnström, Till Bruckner, Katarzyna Klas, Tarik Suljic, Samruddhi Yerunkar, Natasha Olsson, Carolina Cruz, Karolina Strzebonska, Lars Småbrekke, Mateusz T. Wasylewski, Johan Bengtsson, Martin Ringsten, Aminul Schuster, Tomasz Krawczyk, Themistoklis Paraskevas, Eero Raittio, Luca Herczeg, Jan-Ole Hesselberg, Sofia Karlsson, Ronak Borana, Matteo Bruschettini, Shai Mulinari, Karely Lizárraga, Maximilian Siebert, Nicole Hildebrand, Shreya Ramakrishnan, Perrine Janiaud, Emmanuel Zavalis, Delwen Franzen, Kim Boesen, Lars G. Hemkens, Florian Naudet, Sofie Possmark, Rebecca M. Willén, John P. Ioannidis, Daniel Strech, Cathrine Axfors

## Abstract

**Objective:** To systematically evaluate timely reporting of clinical trial results at medical universities and university hospitals in the Nordic countries.

**Study Design and Setting:** In this cross-sectional study, we included trials (regardless of intervention) registered in the EU Clinical Trials Registry and/or ClinicalTrials.gov, completed 2016-2019, and led by a university with medical faculty or university hospital in Denmark, Finland, Iceland, Norway, or Sweden. We identified summary results posted at the trial registries, and conducted systematic manual searches for results publications (e.g., journal articles, preprints). We present proportions with 95% confidence intervals (CI), and medians with interquartile range (IQR). Protocol: https://osf.io/wua3r

**Results:** Among 2,112 included clinical trials, 1,650 (78.1%, 95%CI 76.3-79.8%) reported any results during our follow-up; 1,097 (51.9%, 95%CI 49.8-54.1%) reported any results within 2 years of the global completion date; and 48 (2.3%, 95%CI 1.7-3.0%) posted summary results in the registry within 1 year. Median time from global completion date to results reporting was 690 days (IQR 1,103). 856/1,681 (50.9%) of ClinicalTrials.gov- registrations were prospective. Denmark contributed approximately half of all trials. Reporting performance varied widely between institutions.

**Conclusion:** Missing and delayed results reporting of academically led clinical trials is a pervasive problem in the Nordic countries. We relied on trial registry information, which can be incomplete. Institutions, funders, and policy makers need to support trial teams, ensure regulation adherence, and secure trial reporting before results are permanently lost.

**Plain language summary:** Reporting of results from clinical trials is necessary for evidence-based clinical decision making. We followed up reporting of clinical trials in the Nordic countries sponsored by medical universities and university hospitals. Of 2,112 studies completed 2016-2019 in two major trials registries, about half reported results in any form within 24 months and more than one in five did not report results at all. These results show that there is need for improvement in the reporting of Nordic clinical trials.

**What is new?:**

- Many Nordic registered clinical trials were reported late or not at all.
- Almost one in four trials remained unreported at the end of our search period.
- About half of registered trials had reported results two years after completion.
- Only 2.3% of trials posted summary results in the registry one year after completion.
- Concerted action is needed to improve reporting of Nordic clinical trials.

## Background

Clinical trials are a cornerstone of evidence-based medicine. Completing trials is hard work and requires a joint investment of time and resources by multiple stakeholders. Patients agree to participate, and to contribute their sensitive information, with the premise that results may inform decision-making. Incomplete or delayed reporting impedes this goal and is a preventable research waste (1). The large extent of incomplete and delayed reporting has prompted guidelines and mandates for trial registration and public disclosure of results at official registries (e.g., the European Union Clinical Trials Register (EUCTR) and ClinicalTrials.gov in the United States), by the World Health Organization (WHO), authorities, and funders (2–6). Still, compliance is modest, and many registered studies never have their results uploaded in registries (7–9).

Medicinal products trials in the European Union (EU) are legally mandated by the EU clinical trials regulation to be registered in the EUCTR (10). From 2023 onwards, EUCTR is replaced with the Clinical Trial Information System (CTIS); https://euclinicaltrials.eu. In parallel, applications should be made to the relevant national medicines agency. Approval for marketing in the United States necessitates registration also at ClinicalTrials.gov.

Furthermore, EU clinical trials are legally mandated to submit summary results to the registry as soon as the summary results are available and no later than 12 months after completion (10). The contents of summary results are defined by the EU clinical trials regulation and include trial characteristics and main outcomes. Besides the legal mandate codified in the EU clinical trials regulation, the WHO and the Declaration of Helsinki both require timely reporting of clinical trials (11,12). The WHO joint statement on public disclosure of results from clinical trials recommends that summary results be posted on the registry within 12 months of study completion and a journal article be published within 24 months (11).

As public sector institutions and sponsors for clinical investigations, universities and university hospitals have a particular responsibility to ensure their trials are fully and timely reported. In the United States, Germany, Poland, and Canada, benchmarking investigations have shown that institutional activity in this regard is commonly below expectations (13–19). Previous evaluations of summary results reporting of Nordic clinical trials in the EUCTR indicated worrisome performance (20–24). However, those evaluations relied on summary results of trials on medicinal products reported in the EUCTR and did not attempt to find results published in journals. In this project, we therefore sought to provide a more comprehensive benchmark for all the Nordic countries, enabling comparisons with earlier studies.

We aimed to systematically evaluate the reporting of results from registered clinical trials at all Nordic medical universities and university hospitals. Our project expands the thorough manual search approach developed and validated in the respective German IntoValue project (15,16,25,26) for ClinicalTrials.gov. We further include the EUCTR, the main mandated registry of Nordic medicinal product trials, and we incorporate methods and insights from the EU Trials Tracker (8) (as further described and expanded upon by DeVito (27)) and the Dissemination of Registered COVID-19 Clinical Trials (DIRECCT) project (28,29).

## Methods

This cross-sectional meta-research study followed a prespecified protocol (https://osf.io/wua3r, Nov 8, 2022) that can be consulted for full method details, with amendments in Appendix Table 1, and is reported according to STROBE (30).

**Table 1.** Characteristics of included trials (N=2,112).

| **Trial characteristic<sup>1</sup>** | **All trials,**<br>**N=2,112** | **Trials**<br>**registered in**<br>**the EUCTR**<br>**(with or**<br>**without**<br>**ClinicalTrials**<br>**.gov), n=431** | **Trials**<br>**registered**<br>**only at**<br>**ClinicalTrials**<br>**.gov,**<br>**n=1,681** |
| --- | --- | --- | --- |
| Country |  |  |  |
| Denmark | 1,019 (48.2) | 255 (59.2) | 764 (45.4) |
| Finland | 230 (10.9) | 55 (12.8) | 175 (10.4) |
| Iceland | 10 (0.5) | 3 (0.7) | 7 (0.4) |
| Norway | 328 (15.5) | 33 (7.7) | 295 (17.5) |
| Sweden | 525 (24.9) | 85 (19.7) | 440 (26.2) |
| Registry<sup>2</sup> |  |  |  |
| Registered in the EUCTR | 431 (20.4) | 431 (100) | N/A |
| Registered at ClinicalTrials.gov | 2,007 (95.0) | 326 (75.6) | 1,681 (100) |
| Intervention type<sup>3</sup> |  |  |  |
| Medicinal product | 675 (32.0) | 431 (100) | 244 (14.5) |
| Not medicinal product | 1,437 (68.0) | N/A | 1,437 (85.5) |
| Design (missing, n=3) |  |  |  |
| Randomized controlled | 1,515 (71.7) | 304 (70.5) | 1,211 (72) |
| Non-randomized | 594 (28.1) | 127 (29.5) | 467 (27.8) |
| Masking (missing, n=8) |  |  |  |
| Open label | 1,157 (54.8) | 245 (56.8) | 912 (54.3) |
| Any masking | 947 (44.8) | 186 (43.2) | 761 (45.3) |
| Sponsor type<sup>4</sup> |  |  |  |
| Industry or mixed | 158 (7.5) | 22 (5.1) | 136 (8.1) |
| Only non-industry | 1,954 (92.5) | 409 (94.9) | 1,545 (91.9) |
| Phase |  |  |  |
| I | 65 (3.1) | 12 (2.8) | 53 (3.2) |
| I-II | 30 (1.4) | 11 (2.6) | 19 (1.1) |
| II | 196 (9.3) | 149 (34.6) | 47 (2.8) |
| II-III | 21 (1.0) | 8 (1.9) | 13 (0.8) |
| III | 61 (2.9) | 39 (9) | 22 (1.3) |
| IV | 240 (11.4) | 196 (45.5) | 44 (2.6) |
| Not given | 1,499 (71.0) | 16 (3.7) | 1,483 (88.2) |
| Multicentric |  |  |  |
| Yes | 526 (24.9) | 136 (31.6) | 390 (23.2) |
| No (monocentric) | 1,586 (75.1) | 295 (68.4) | 1,291 (76.8) |
| Enrollment (actual or anticipated; missing, n=1) |  |  |  |
| 1 - 100 | 1,463 (69.3) | 318 (73.8) | 1145 (68.1) |
| 100 - 500 | 515 (24.4) | 100 (23.2) | 415 (24.7) |
| > 500 | 133 (6.3) | 13 (3) | 120 (7.1) |
| Completion year (actual or anticipated) |  |  |  |
| 2016 | 515 (24.4) | 107 (24.8) | 408 (24.3) |
| 2017 | 532 (25.2) | 134 (31.1) | 398 (23.7) |
| 2018 | 556 (26.3) | 91 (21.1) | 465 (27.7) |
| 2019 | 509 (24.1) | 99 (23.0) | 410 (24.4) |
| Status |  |  |  |
| Completed | 1,675 (79.3) | 349 (81.0) | 1,326 (78.9) |
| Terminated | 149 (7.1) | 61 (14.2) | 88 (5.2) |
| Unknown status | 288 (13.6) | 21 (4.9) | 267 (15.9) |

Percentages may exceed 100 because of rounding. <sup>1</sup>Total count may differ between categories because of missing data (number missing shown if >0). <sup>2</sup>Subcategories are not mutually exclusive. <sup>3</sup>Medicinal product trials included all EUCTR trials, and ClinicalTrials.gov trials with the labels drug, biological, genetic, or combination product; non-medicinal product trials included ClinicalTrials.gov trials with the labels behavioral, device, diagnostic test, dietary supplement, procedure, radiation, or other. <sup>4</sup>Among lead or collaborating sponsors.

## Eligibility Criteria

We included clinical trials that were led by any university with a medical faculty (medical university) or university hospital in the Nordic countries (Denmark, Finland, Iceland, Norway, and Sweden), and completed in 2016-2019 as per their registration in the EUCTR or at ClinicalTrials.gov. Other locally used registries not internationally recognized as trial registries were not considered (e.g., ResearchWeb, “Cancer studies in Sweden”, or Open Science Framework) (6,31,32). University hospitals may be a conglomerate of several smaller hospitals, e.g., Copenhagen University Hospital (33,34). Similarly for Stockholm, we included Danderyd Hospital and Stockholm South General Hospital (Södersjukhuset) as they are university-affiliated. Trials were included regardless of phase, population, intervention, comparator, or outcome. We considered a trial to be “led by” the medical university or university hospital if the institution was listed in the EUCTR as sponsor, or at ClinicalTrials.gov as lead sponsor and/or responsible party (35). We excluded trials led by an administrative region (e.g., “Region Skåne”) and trials withdrawn without enrollment. For eligible trials present in both registries, we preferred EUCTR information for being the mandatory registry, but used ClinicalTrials.gov data on status or completion dates if these were more recently updated.

As results publications, we included peer-reviewed scientific publications, preprints, doctoral theses, or published congress abstracts, with >500 words, as shorter abstracts would not be likely to permit comprehensive results reporting. The publication had to be matched to an eligible clinical trial regarding design, indication/population, intervention/treatment, and comparator if applicable. Also, the publication’s primary outcome measure (or first mentioned outcome) had to be listed as an outcome measure in the registration, whether as primary or not. Our goal was to identify correct matches, not to evaluate discrepancies in primary outcome reporting.

As summary results, we included summaries posted in the EUCTR or ClinicalTrials.gov directly as tabular results, or in a file (e.g., Clinical Study Report synopsis, as observed in previous work ()), but not links to external files or publications outside the registration page (this only occurred in two cases). We also included EUCTR results sections without analysis results, but with a plausible justification (e.g., low enrollment), as in previous work (27).

## Data collection

### EUCTR

We retrieved the latest dataset for the EU Trials Tracker (8,36) of EUCTR trials on Nov 27, 2022, with data from Nov 7, 2022. Trials registered in the EUCTR can have EudraCT protocols, one for each EU/EEA country where the trial is conducted, and a results section. Trials without a EudraCT protocol were not present in the dataset (i.e., not uploaded or not publicly available). We accessed automatically extracted data from EUCTR country protocols and results sections (Appendix Table 2) from a custom web scraper by the EU Trials Tracker team (36). Automatically identified results sections were manually verified for eligibility as summary results. Global completion dates were preferred from results sections for being the most recently updated.

### ClinicalTrials.gov

We downloaded the Aggregate Analysis of ClinicalTrials.gov dataset (AACT, http://aact.ctti- clinicaltrials.org<u>/</u>) on Nov 27, 2022, with data from Nov 9, 2022. We automatically excluded observational studies and duplicated entries and identified the sample with a search string for medical universities and university hospitals (Appendix Table 3). We adapted code from the IntoValue project (25,37) to extract variables (Appendix Table 2), e.g., whether a trial had summary results posted at the registry.

### Manual data extraction

Our comprehensive systematic manual search for publications, by two independent reviewers per trial, followed a stepwise process (Appendix Figure 1). First, we identified results publications linked at the registration website. Second, we searched Google with the main trial ID (i.e., EudraCT number or NCT ID; then other trial IDs if present, one at a time), and screened the first page (minimum 10 hits). Third, if no publication was so far identified, we performed another Google search with two combinations of terms from the registry (e.g., title, principal investigator, intervention, disease, or other). Publication dates were extracted automatically through PubMed (for articles with PubMed IDs) and otherwise manually from the publication, preferring the earliest date among multiple available dates or eligible publications. Discrepancies regarding the earliest eligible results publication for each trial were solved by one reviewer with access to the two independent extractions and comments, and uncertainties discussed with another reviewer. We publicly share our dataset with all identified potentially eligible results publications (https://github.com/cathrineaxfors/nordic-trial-reporting).

To pilot and refine the extraction forms, two reviewers (SW and CA) independently analyzed a random set of 10 trials. For training, all reviewers initially analyzed the same 10 trials, with personal feedback after checking agreement against SW and CA. Reviewers repeated the training on another random set of 5 trials if the agreement was <100% for the existence of any eligible results publication, or if they found fewer eligible publications for >20% of trials. This occurred for 6/31 reviewers (19%).

### Statistical Analysis

Descriptive statistics for primary outcomes are presented for the total sample, per country, and per accountable institution (medical university, university hospital, or if applicable, member hospitals of the university hospital, Appendix Table 3). Trials with multiple accountable institutions were counted for each one. The following four primary outcomes were prespecified: (a) *Any results reported within 2 years of completion*: number and percent, with 95% confidence interval using Wilson score, of registered clinical trials that shared results either in a publication or as summary results in the trials registry within 2 years after global completion, as the upper limit of what is mandated or ethically expected (given the WHO joint statement on reporting of clinical trials in journals) (2,4). (b) *Summary results reported within 1 year of completion:* number and percent, with 95% confidence interval using Wilson score, of registered clinical trials that have summary results posted within 12 months after completion (corresponding to the time frame mandated by the EU clinical trials regulation to upload summary results in the EUCTR). For trials at ClinicalTrials.gov, we apply the same timeline for consistency and to illustrate adherence to best practices (2). (c) *No results reported:* Number and percent (with 95% confidence interval using Wilson score) of registered clinical trials that remained unreported. (d) *Time to reported results:* We calculated median time (and interquartile range) from trial completion (last patient’s last visit for all outcomes) to the reporting of summary results or results publication, whichever occurred first, taking into account differential follow-up. This is also illustrated with a Kaplan-Meier curve. For unpublished trials, we censored the timeline at the latest date that searches were performed for that trial.

We report the following three secondary outcomes: (a) Participants in trials that are unreported (planned number) and reported (actual number, or if missing, planned number) according to registry information. (b) Prospective registration: percent (with 95% confidence interval using Wilson score) of clinical trials that are registered before the start date of the study. This outcome is also illustrated with a Kaplan-Meier curve (time to registration after trial start). It was only calculated for trials registered at ClinicalTrials.gov, since EUCTR entries are made by national regulators (27). (c) Number and percent of medicinal products trials identified at ClinicalTrials.gov, excluding intervention category “dietary supplements” and phase I trials, which do not have a cross-registration in the EUCTR.

We also explored subgroup analyses for binary primary outcomes: (a) country (Denmark, Finland, Iceland, Norway, Sweden); (b) intervention type (medicinal products vs. other interventions); (c) single-center vs. multicenter trials; (d) industry-sponsored vs. other trials; and (e) enrollment (smallest, medium, and largest tertile), as given in registry, whether actual or planned. We applied post-hoc statistical significance testing (Pearson’s chi-squared test), Bonferroni-corrected for the number of tests (n=15, i.e., p<0.05/15).

Sensitivity analyses were performed to explore the robustness of results for the binary primary outcomes: (a) randomized controlled trials only; (b) trials registered in the EUCTR (with or without cross-registration); (c) trials registered only at ClinicalTrials.gov; (d) excluding trials with missing status or status “Unknown”; (e) recalculating institutional results only counting trials for an institution if it was the sole sponsor or listed *first* among sponsors; (f) using primary completion date instead of global; and (g) trials registered only at ClinicalTrials.gov that had an actual completion date (not anticipated).

## Results

We included 2,112 registered trials led by 54 Nordic medical universities and university hospitals (Figure 1). Twenty-two trials were co-led by two different eligible institutions. Most trials were on non-drug interventions, for which phase allocation was generally not applicable (Table 1). Predominant characteristics were academic-only sponsorship, small size (n<100), and single-center organization. Denmark was the major contributor of clinical trials with 48% of the total Nordic output. In the manual searches for results publications (Nov 21, 2022 to March 23, 2023), for 75% of trials, both reviewers found no publication or identified the same earliest publication; for 13%, both found at least one publication, but the earliest among these was not the same; and for 11%, one reviewer found a publication and not the other. Among trials with a results publication, the earliest publication was most often a journal article (1,574/1,601, 98.3%), and rarely a preprint (18/1,601, 1.1%) or other type (9/1,601, <1%; letter to the editor, n=4, doctoral thesis, n=3, conference abstract, n=2). Half of the earliest publications were linked at a registry (790/1,601, 49.3%). The other half were found only through our systematic Google search (768/1,601 48%) or cross-referenced in a linked publication (43/1,601, 2.7%).

**Figure 1.**
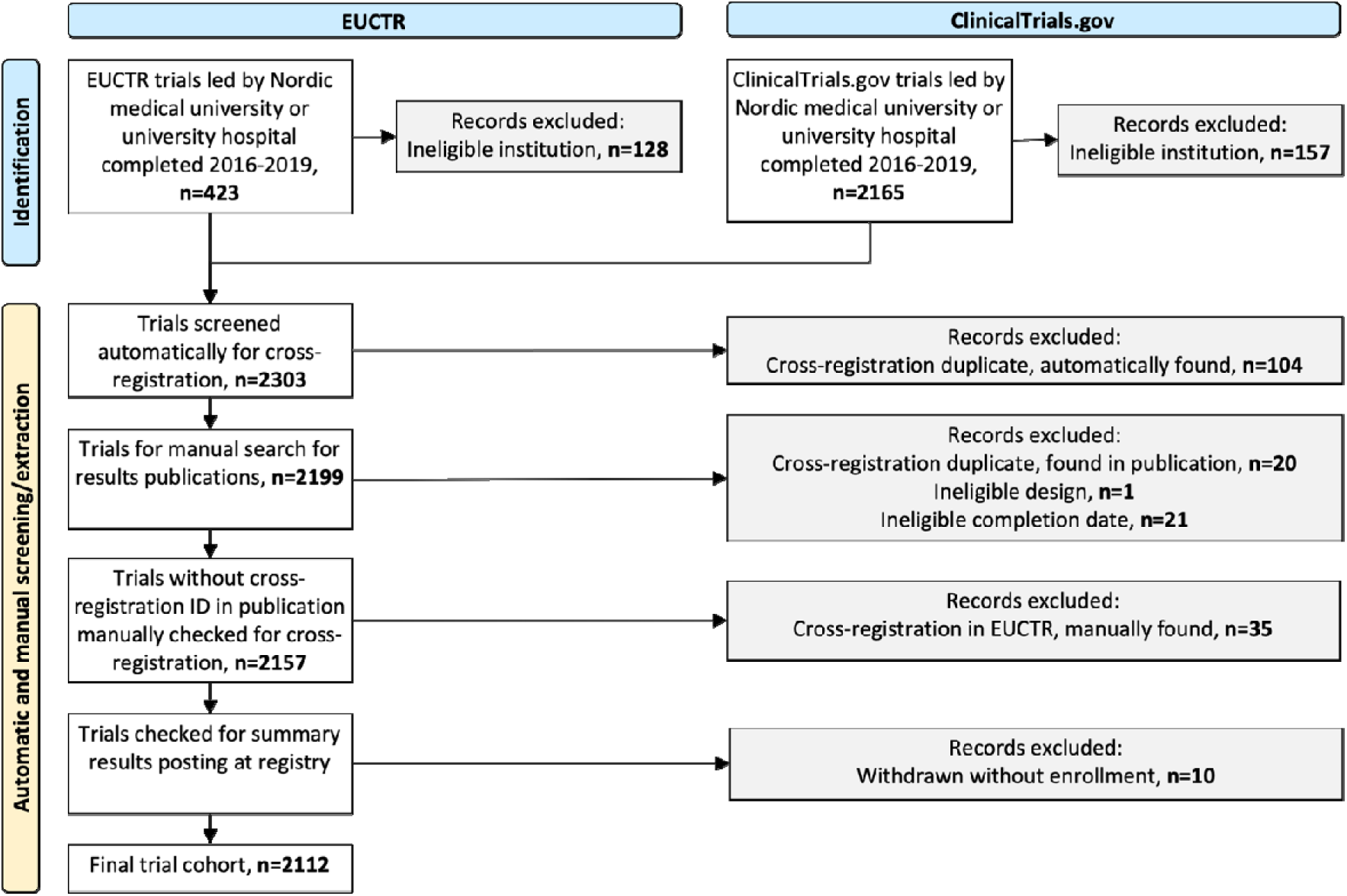
Flowchart for the search process. Note. EUCTR trials were identified in the EU Trials Tracker dataset (which excludes EUCTR records without any linked EudraCT protocol). ClinicalTrials.gov trials were identified in the AACT dataset (after excluding duplicate records and observational studies). Automatic screening for cross-registrations was done in the statistical program R (and manually verified). Manual search for cross-registration IDs was done in all results publications. Additional manual searches for cross-registrations were done for all EUCTR-registered trials and for ClinicalTrials.gov-registered medicinal product trials. Cross-registered records were merged to use the most updated information. Summary results were manually verified for all EUCTR trials to exclude trials withdrawn without any enrollment.

## Primary outcomes

Just more than half of all trials were reported within 2 years of completion (1,097/2,112, 51.9%, 95%CI 49.8-54.1%), most often only as a journal article (946/1,097). Few trials had summary results posted in a trial registry within one year of completion (48/2,112, 2.3%, 95%CI 1.7-3%). At any time, across a follow-up time of 2.9 to 7.1 years after completion, summary results were still uncommon (296/2,112, 14.0%, 95%CI 12.6-15.6%) and just over three-fourths of trials reported any results, whether as summary results or a results publication (1,650/2,112 trials, 78.1%, 95%CI 76.3-79.8%). A median of 22.7 months passed between completion and first results reporting (690 days, IQR 1,103 days). The performance of individual institutions varied widely (Figure 2; Appendix Tables 4–7). Time to first results reporting is displayed in Figure 3 and per country in Appendix Figure 2. For 174 trials, results were reported before the global completion date (i.e., the day before or earlier).

**Figure 2.**
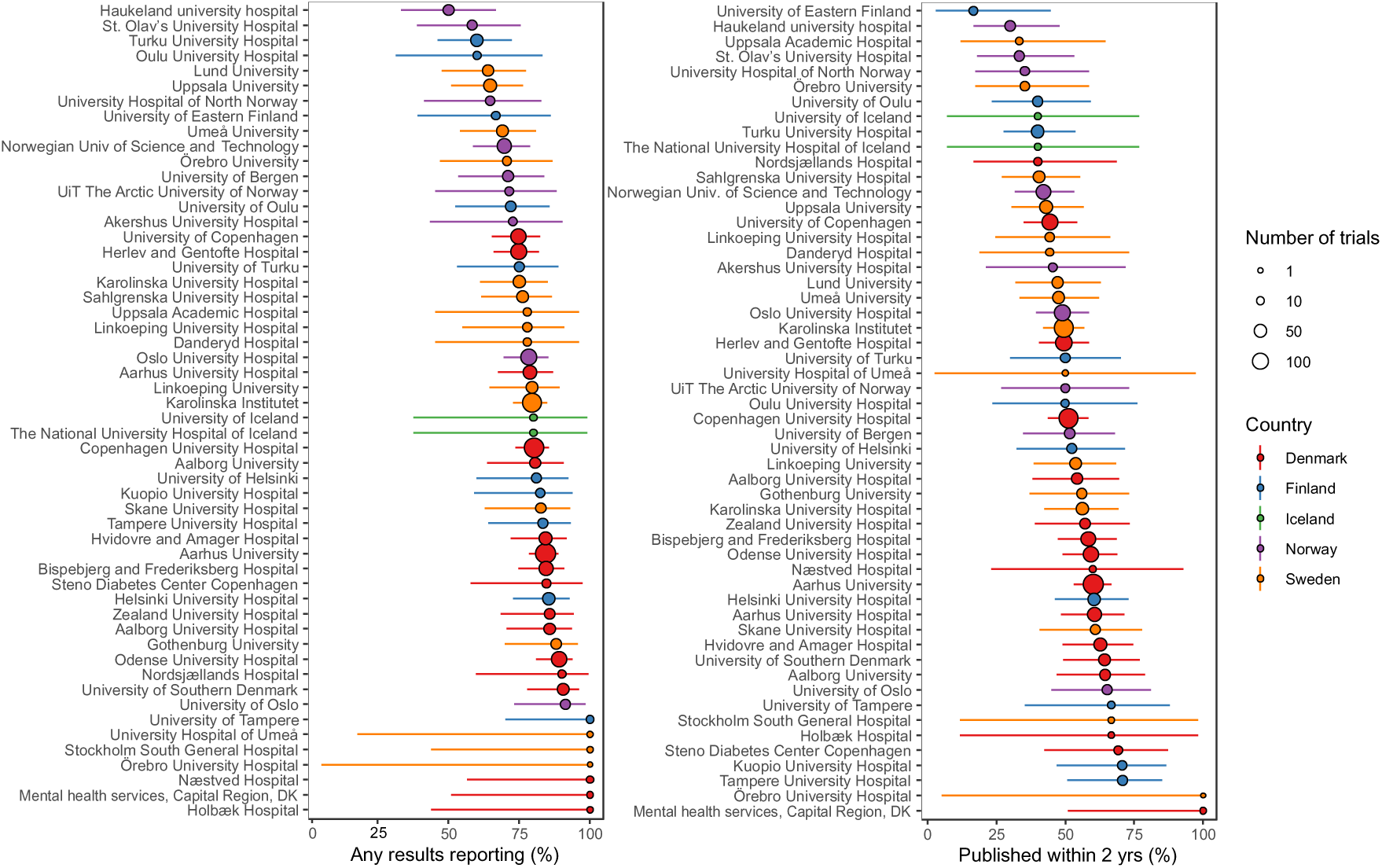
Share of trials with any results reporting (summary results or results publications) at the end of follow-up, and within two years of completion. Note. Confidence intervals calculated with Wilson score (using modified Wilson score if the numerator, or the denominator minus numerator, was less than 3).

**Figure 3.**
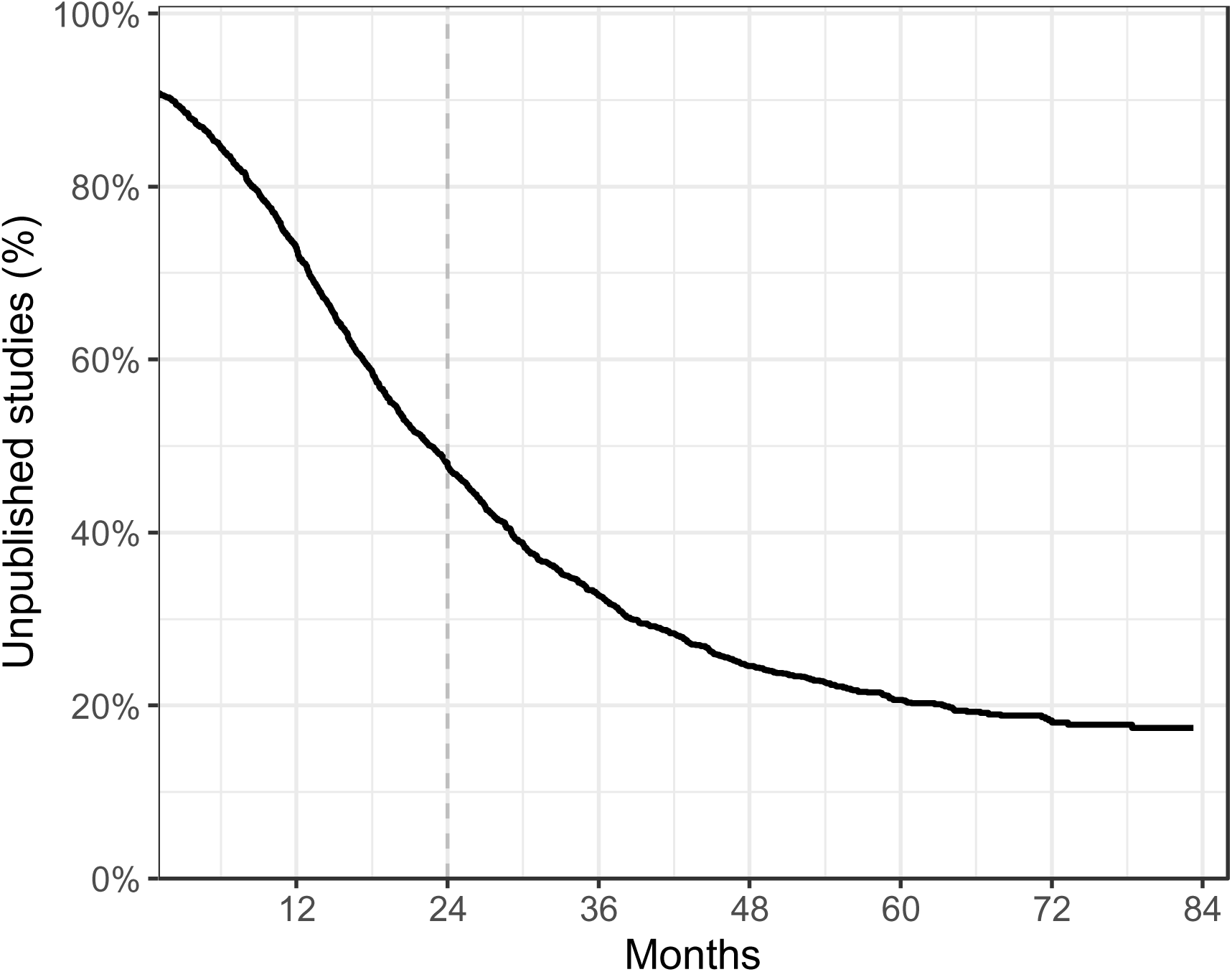
Time to first reported results (summary results or results publication, total sample, N=2,112). Note. For 174 trials (8.2%), results were reported before the completion date. The lowest point of the Kaplan-Meier curve is not the same as the overall proportion that remained unpublished (21.9%) since the curve shows the proportion of unpublished trials among those followed up to that specific point. The curve is restricted to 84 months due to limited data beyond this follow-up (3 trials, one of which had a publication at 85 months).

## Secondary outcomes

Unreported trials (n=462) had a median planned sample size of 42 (IQR 80, range 1-15,030) with a potential total of 83,489 individuals (Denmark n=51,020, Finland n=5,878, Iceland n=1,573, Norway n=11,972, and Sweden n=13,046). For reported trials (n=1,650), median sample size (actual or planned) was 58 (IQR 116.75, range 2-150,000), with a total of 865,781. Prospectively registered trials among those only registered at ClinicalTrials.gov were overall 50.9% (856/1,681) and varied widely between institutions (Appendix Figure 3, Appendix Table 8). Another 14% (n=236) were registered within 60 days, and 35% (n=589) more than 60 days, after their start (Appendix Figure 4). Among trials registered at ClinicalTrials.gov likely falling under the requirement for registration in the EUCTR (non-phase-I medicinal products trials), less than two-thirds were found also in the EUCTR (195/333, 58.6%, 95%CI 53.2-63.7%).

**Figure 4.**
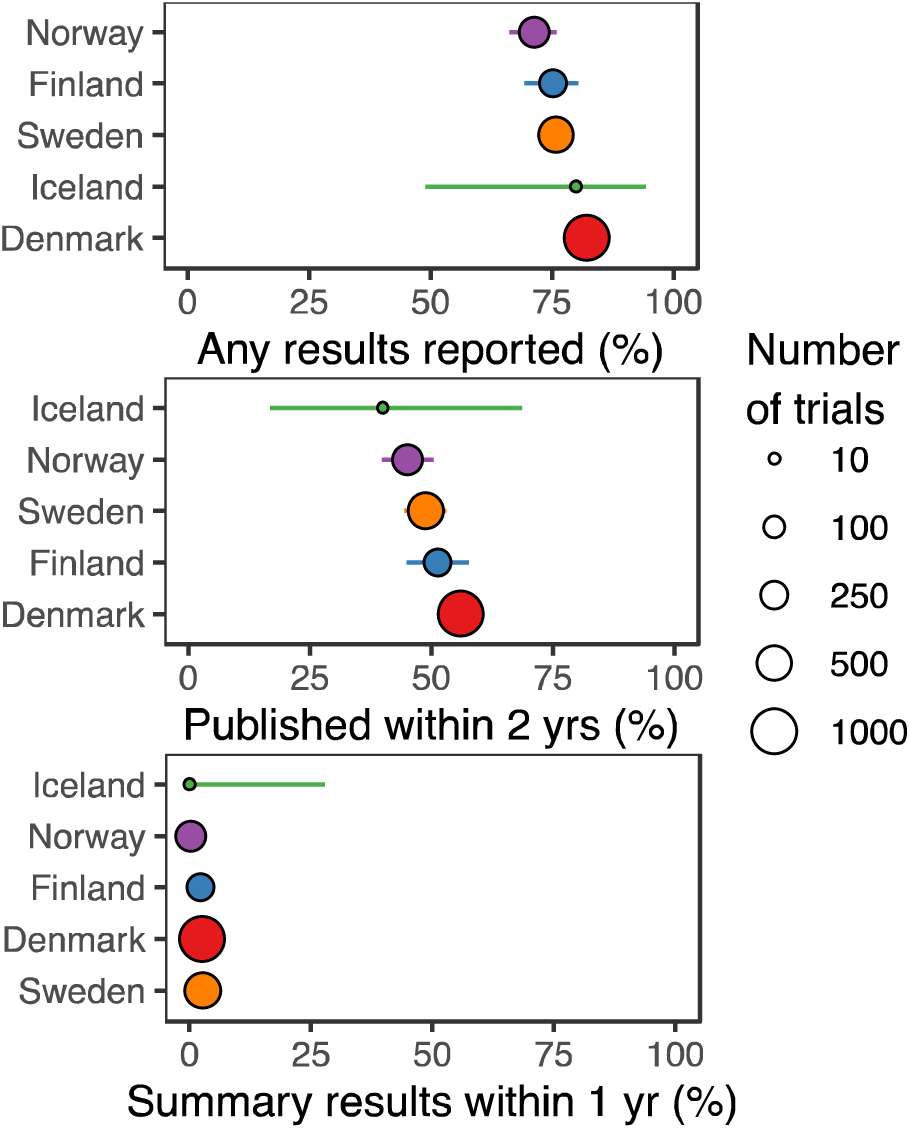
Reporting of trial results per Nordic country. Note. Confidence intervals calculated with Wilson score (using modified Wilson score if the numerator, or the denominator minus the numerator, was <3).

## Subgroup analyses

Denmark presented the highest percentage of reported trials among the Nordic countries (Figure 4), and Norway the lowest. Large trials (vs. small trials) were more likely to be reported at all, and within 2 years of completion; and medicinal product trials (vs. non-medicinal product trials) were more likely to have summary results. We found no differences per multicenter status or industry-sponsored status (Appendix Table 9).

## Sensitivity analyses

Reporting was better for EUCTR-registered trials, and worse for only ClinicalTrials.gov- registered trials, than the overall sample (Appendix Table 10). Reporting of any results, and within 2 years of completion, was higher among completed trials than those terminated or

with unknown status (Appendix Table 10). Other sensitivity analyses were consistent with the main results (Appendix Tables 10–13).

## Discussion

### Summary of findings

Only 52% of clinical trials led by Nordic medical universities or university hospitals were reported in adherence to WHO recommendations (11). Timely summary results reporting in the registries was rare, with only 2% of registrations showing results 12 months after completion. Among EUCTR registrations, for which this practice is now mandatory, the proportion was 8%. For 22% of trials (planned to involve 83,489 participants), no results could be located regardless of time limit. Contrary to established recommendations, almost half of registrations were made retrospectively, limiting their usefulness (38,39). Among trials registered at ClinicalTrials.gov likely falling under the requirement for registration in the EUCTR, short of two-thirds were found also in the EUCTR. These results show that registration and reporting of clinical trials in the Nordic countries can be considerably improved.

### Comparison to earlier research

Overall, results from the present project are similar to earlier findings in other settings. The IntoValue project followed up reporting of trials from German university medical centers, registered in ClinicalTrials.gov or in the German Clinical Trials Register (DRKS) (14–17). Our present results are similar to theirs, where out of 1,658 trials completed in 2014–2017, 43% published results either as summary results or in journal articles within 24 months after completion, and five years after completion, 70% of the trials were published (15). In Canada, a follow-up of 6,720 trials on ClinicalTrials.gov from 2009-2019 showed that 59% were registered prospectively and 39% reported summary results in the registry (19). This proportion is considerably higher than in the present study, mainly because they included many industry-sponsored studies, among which summary results reporting was more common. Of trials registered 2009-2014, 55% were subsequently published in an academic journal (19). In Poland, among 305 interventional clinical trials at academic medical centers registered on ClinicalTrials.gov and completed 2009–2013, 80% had been published as articles or posted summary results in 2019 (18). Results were posted within a year of study completion and/or published within 2 years of study completion for 43% of trials. In the United States, a sample of 400 trials from ClinicalTrials.gov was followed up for 36 months and results were found for 61% of trials either in the registry or on PubMed (40).

### Strengths and limitations

A strength of this project is that extensive online manual searches were carried out to track results publications, inspired by and further developing previous methods (14–17).

Investigators were trained with sample searches and two independent investigators performed searches for each trial, relying not only on trial identifiers but also other characteristics. Although some results publications may still have been missed, we believe that a high coverage has been reached. A further strength is the careful manual work to identify and validate cross-registrations.

The following limitations should be noted. This project relied on information in trial registries, which can be incomplete and not updated since the initial entry. For instance, completion date and enrollment numbers can reflect what was estimated at the start, rather than actual numbers. A recent study of registered Covid-19 trials found that registrations with actively updated completion status showed a higher proportion of results reported compared to others (41). We did not contact primary investigators nor sponsors to verify trial status; however, a survey in a sample of German registered trials showed only minor changes after status verification by the primary investigator (42). The planned number of participants in unreported trials is likely an overestimation compared to final enrollment (43,44), but the lack of results for their trials still reflects extensive research waste. The registries contain a heterogeneous sample of trials, some of which are intended to inform clinical practice, and some of which are closer to basic research.

In the EUCTR, registration dates and start dates have particular limitations (27). While all trials on the EUCTR should be prospectively registered as they would need approvals tied to their registration before starting, it is impossible to tell from the registration when a trial actually started. Also, primary completion dates (with respect to primary outcome) are not generally registered, and we instead had to use global completion dates (last patient’s last recorded data point). For trials with long follow-up of secondary outcomes, this may be much later, and contributes to explaining the proportion of trials reported before their completion date. Using a stricter cutoff, numbers for timely reporting may be even lower. We expect many trials to erroneously appear as ongoing although they are actually completed or terminated (45); for Norway, this affects a particularly large proportion of trials (24). Timely reporting may have been overestimated, if trials without status update are less likely to have reported results. In contrast to the EUCTR, ClinicalTrials.gov lists anticipated completion dates, which allowed us to include trials without an active status update. However, since some of these trials may be still ongoing, this may partly explain the difference in results between the two registries. In our sensitivity analysis among ClinicalTrials.gov-registered trials with “actual” completion dates results were in-between.

Cross-registration of trials in our examined registries varied in extent and content overlap. If some were without overlapping or similar titles, we may have missed them and hence double-counted some trials. These might appear unreported if summary results were posted in one registration and not the other. Information in cross-registrations was often inconsistent in terms of key study dates and status. We chose to merge cross-registrations to use the most updated information.

Not all trials are registered, and those not registered may be more likely to also not be reported (27,46). Trials that were not prospectively registered may have had a higher likelihood of becoming registered if the investigators were in the process of reporting. Also, we did not scrutinize the quality of the reporting nor alignment with registered outcomes.

These reasons may contribute to overestimating the proportion of meaningfully reported trials.

### Implications and future perspectives

Nordic academic sponsors need to improve clinical trials registration and reporting. Joint work is needed by multiple stakeholders to enable continuous monitoring of clinical trials reporting and to align resource allocation with best practices. The Nordic Trial Alliance Working Group on Transparency and Registration has recommended institutions to implement standard operating procedures to support their researchers and ensure complete registration and reporting (47). Resources summarizing how universities can improve clinical trial reporting are also compiled online (e.g., https://www.transparimed.org/resources).

Possible solutions are already implemented by some sponsoring institutions, e.g., as recently reported by one Nordic institution (Karolinska Institutet) with dedicated support staff for trial registration and active follow-up of reporting (48). In Germany, efforts are ongoing to involve trial funders in creating and implementing strategies for increased reporting (49), and other examples on lobbying activities or infrastructural support structures exist in the United States and United Kingdom (50–52).

There may be several reasons for slow or missing reporting of clinical trials (27). Trials may be underfunded, and institutional support may be lacking, leaving the individual researcher with sole responsibility for trial reporting. Summary results posting can be difficult for researchers unfamiliar with registry interfaces and procedures. Reporting of results in journals typically requires manuscripts to pass editorial and/or peer review, a process largely outside of the researcher’s control. More fundamentally, current models for assessing academic merits may not be well aligned with transparent reporting (53). Researchers may perceive journal publications and journal impact factors as main criteria for funding and promotion. This misalignment may be to the detriment of reporting summary results in registries, or reporting negative results or those contrary to established opinion and therefore more difficult to publish in journals. These obstacles highlight the role of preprints as a vehicle for rapid and unhindered dissemination of research results. Assessments of the merit of articles reporting clinical trials could be improved by taking into account whether studies were prospectively registered and results reported in a timely and complete manner (54).

## Conclusions

We found considerable room for improvement of registration and reporting of Nordic clinical trials. Late and incomplete reporting reduces the accuracy and validity of the evidence base used for clinical decision making and hampers the advancement of knowledge. Future extensions should be made for industry-sponsored trials (55) and trials in other registries.

Joint action by stakeholders is needed, especially at the initiative of academic institutions, to support, monitor, and incentivise trials registration and timely reporting.

## Declarations

### Data sharing

Data and code used for our analyses are publicly available at https://github.com/cathrineaxfors/nordic-trial-reporting and https://zenodo.org/records/10091147.

## Author contributions

**Conceptualization:** G.N., S.W., N.J.D., M.S.-H., S.Y., L.S., M.R., E.R., J.-O.H., M.B., S.M., P.J.,

T.B., K.B., M.C., L.G.H., F.N., S.P., R.M.W., J.P.I., D.S., and C.A.

**Investigation (data collection):** S.W., K.K., T.S., S.Y., N.O., C.C., K.S., L.S., M.W., J.B., M.R., A.S., T.K., T.P., L.A., E.R., L.H., J.-O.H., S.K., R.B., M.B., S.M., K.L., M.S., N.H., S.R., P.J., E.Z., T.B., D.F., and C.A.

**Methodology:** G.N., S.W., N.J.D., M.S.-H., T.B., J.P.A.I., D.S., and C.A.

**Data curation:** C.A. and L.A.

**Formal analysis, Funding acquisition, Project Administration, Resources, Supervision, Visualization:** C.A.

**Software:** N.J.D., M.S.-H., and C.A.

**Validation:** L.A., K.K., and C.A. **Writing - original draft:** G.N. and C.A. **Writing - review & editing:** All authors.

## Data Availability

All data produced are available online at https://zenodo.org/records/10091147
All code used for data processing available online at
https://github.com/cathrineaxfors/nordic-trial-reporting

https://zenodo.org/records/10091147

https://github.com/cathrineaxfors/nordic-trial-reporting

## Acknowledgements

CA reports funding for this project by the Knut and Alice Wallenberg Foundation through the Wallenberg Foundation Postdoctoral Scholarship Program at Stanford (KAW 2019.0561).

FN reports funding outside this project from the French National Research Agency (ANR-17- CE36-0010), the French ministry of health and the French ministry of research; being work package leader in the OSIRIS project (Open Science to Increase Reproducibility in Science) with funding from the European Union’s Horizon Europe research and innovation programme (grant agreement No. 101094725); and being work package leader for the doctoral network MSCA-DN SHARE-CTD (HORIZON-MSCA-2022-DN-01 101120360), funded by the EU.

The title page and author contributions section of this document was made using the free open-source tool *tenzing*, https://rollercoaster.shinyapps.io/tenzing/.

## Appendix

Appendix to “Results reporting for clinical trials led by medical universities and university hospitals in the Nordic countries was often missing or delayed” by Nilsonne et al.

---

**Figure 1.**
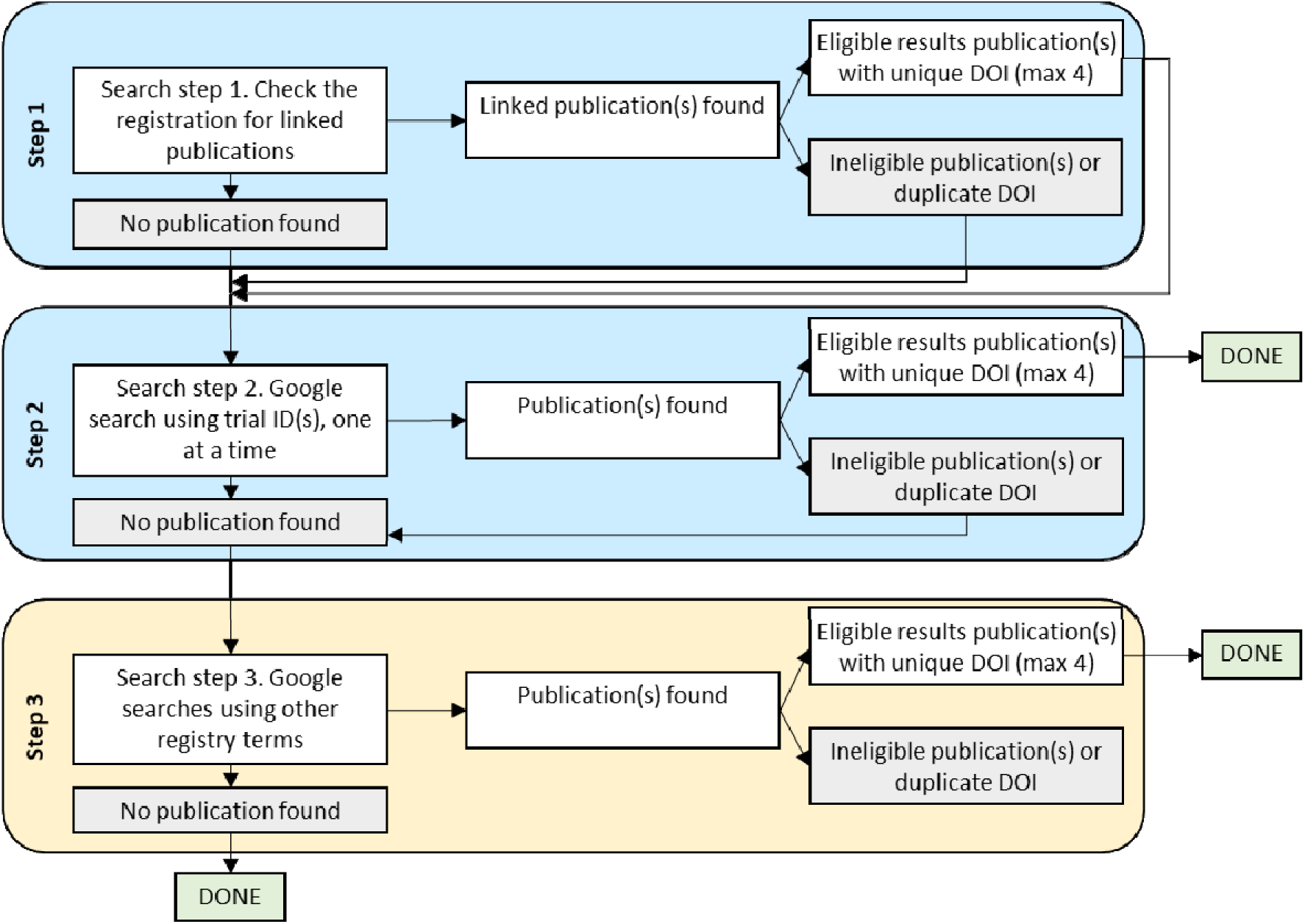
Overview of manual identification of results publications

**Figure 2.**
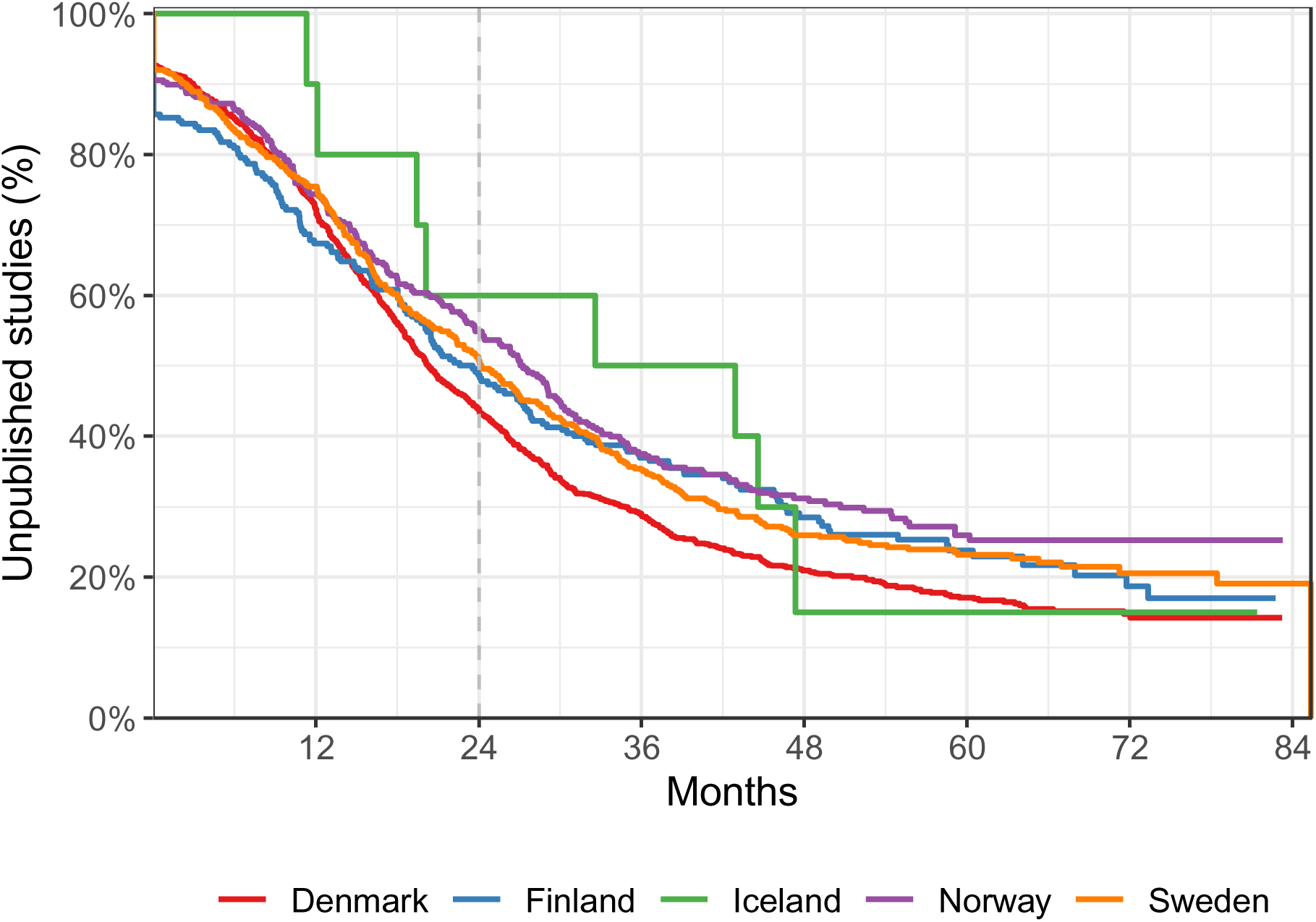
Time to first reported results (summary results or results publication, total sample, N=2,112) per country.

**Figure 3.**
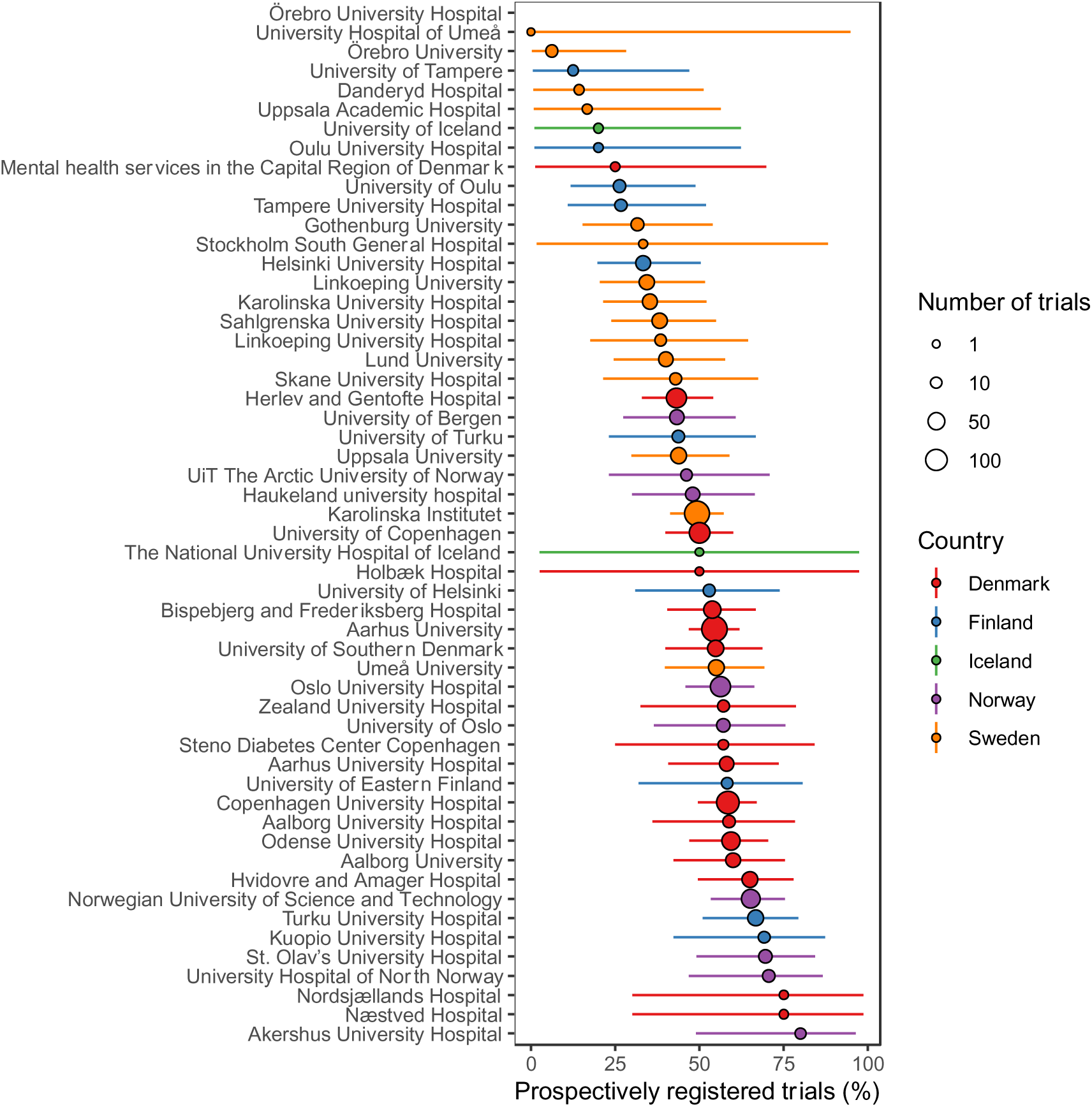
Share of trials with prospective trial registration among those registered at ClinicalTrials.gov only (n=1,681)

**Figure 4.**
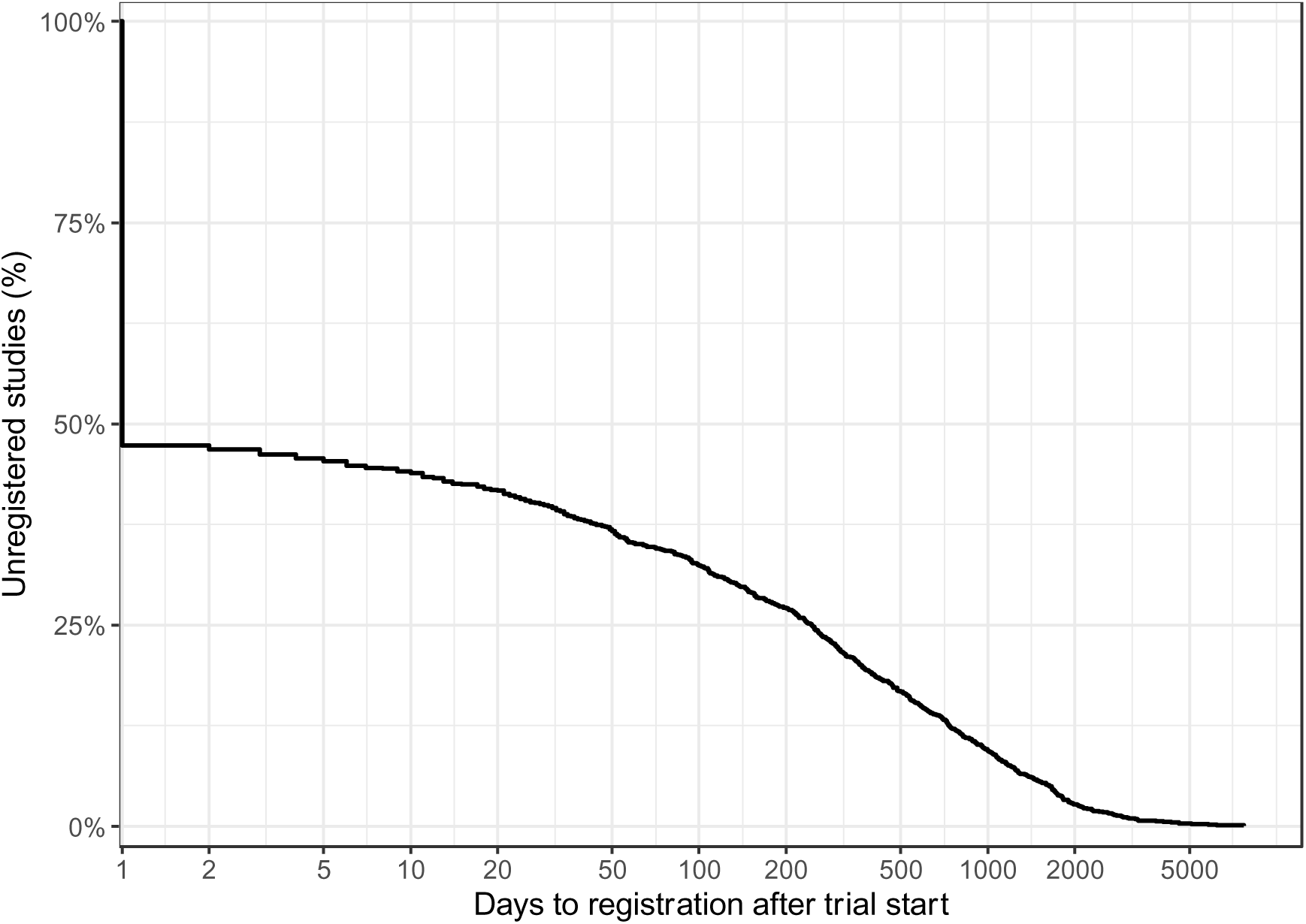
Time to trial registration after start date among trials registered at ClinicalTrials.gov only (n=1,681)

**Table 1.**
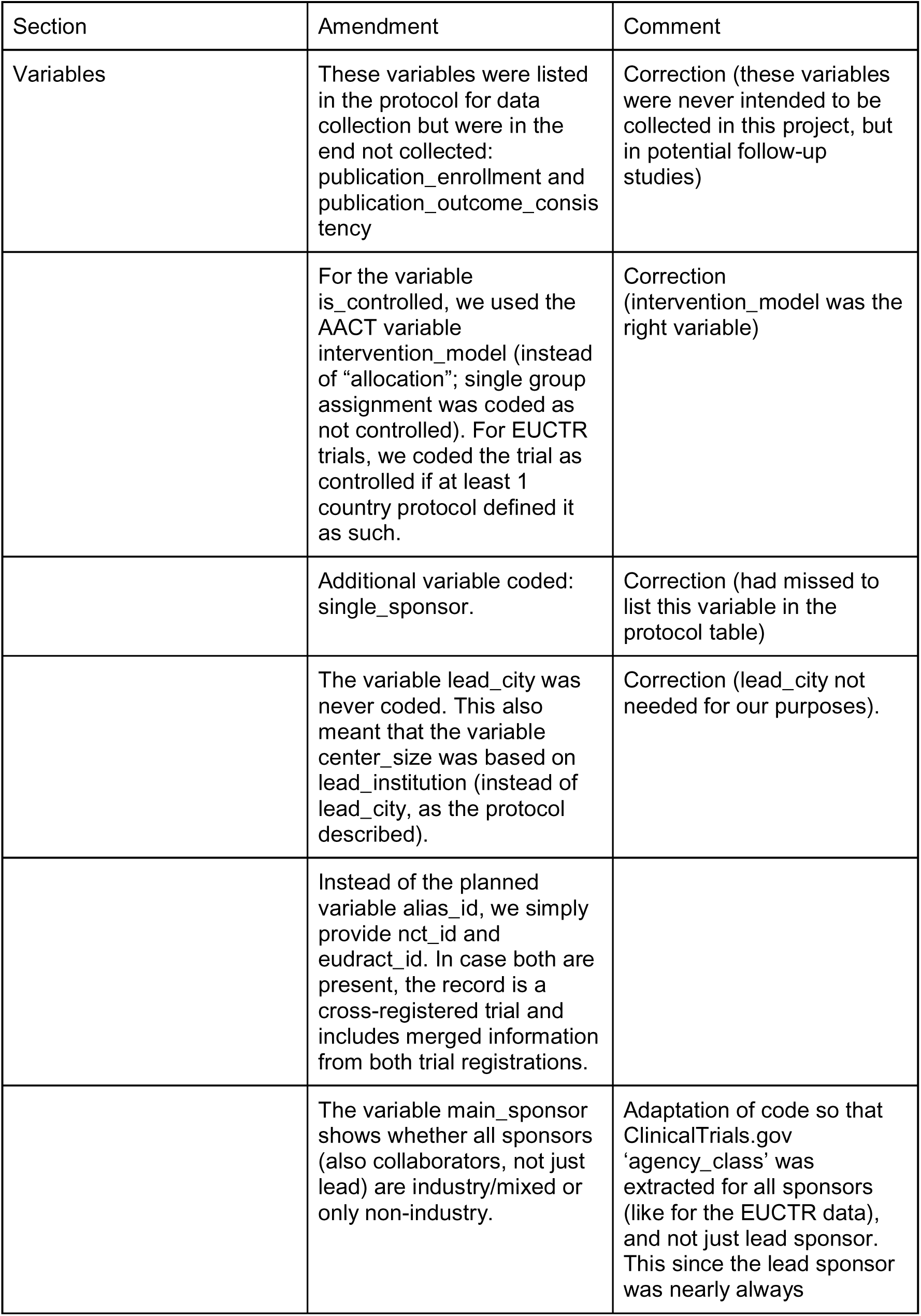
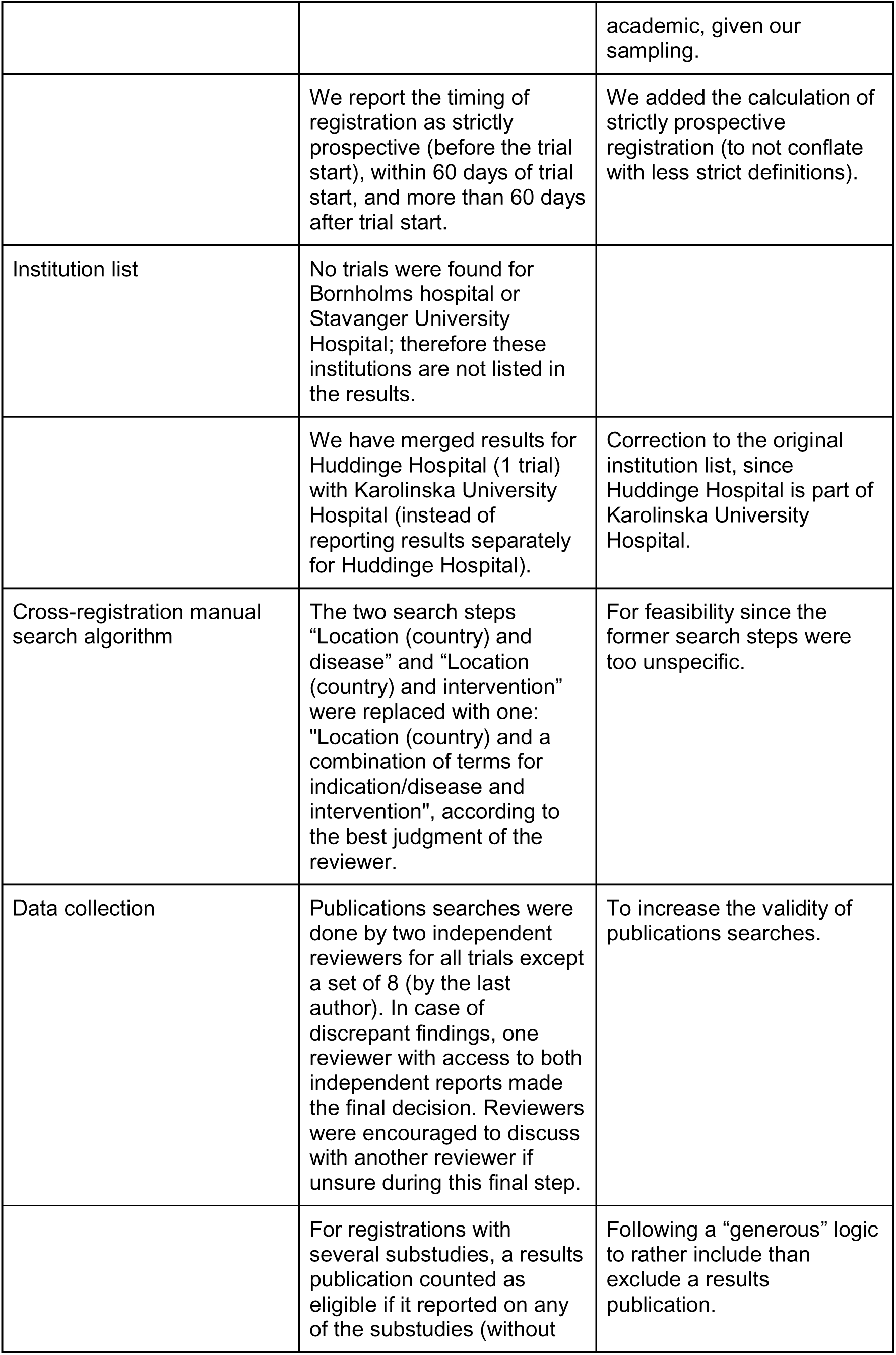
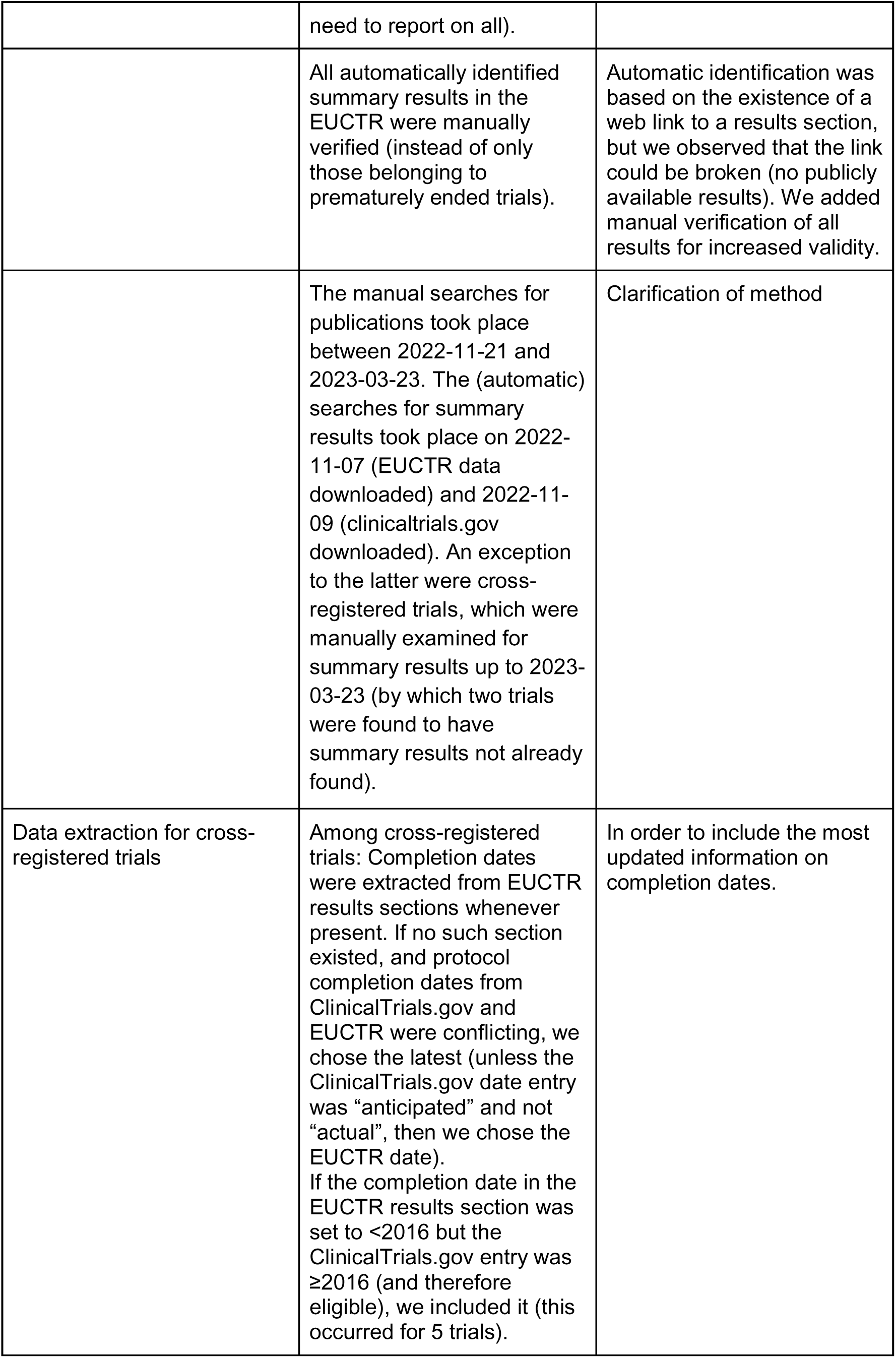
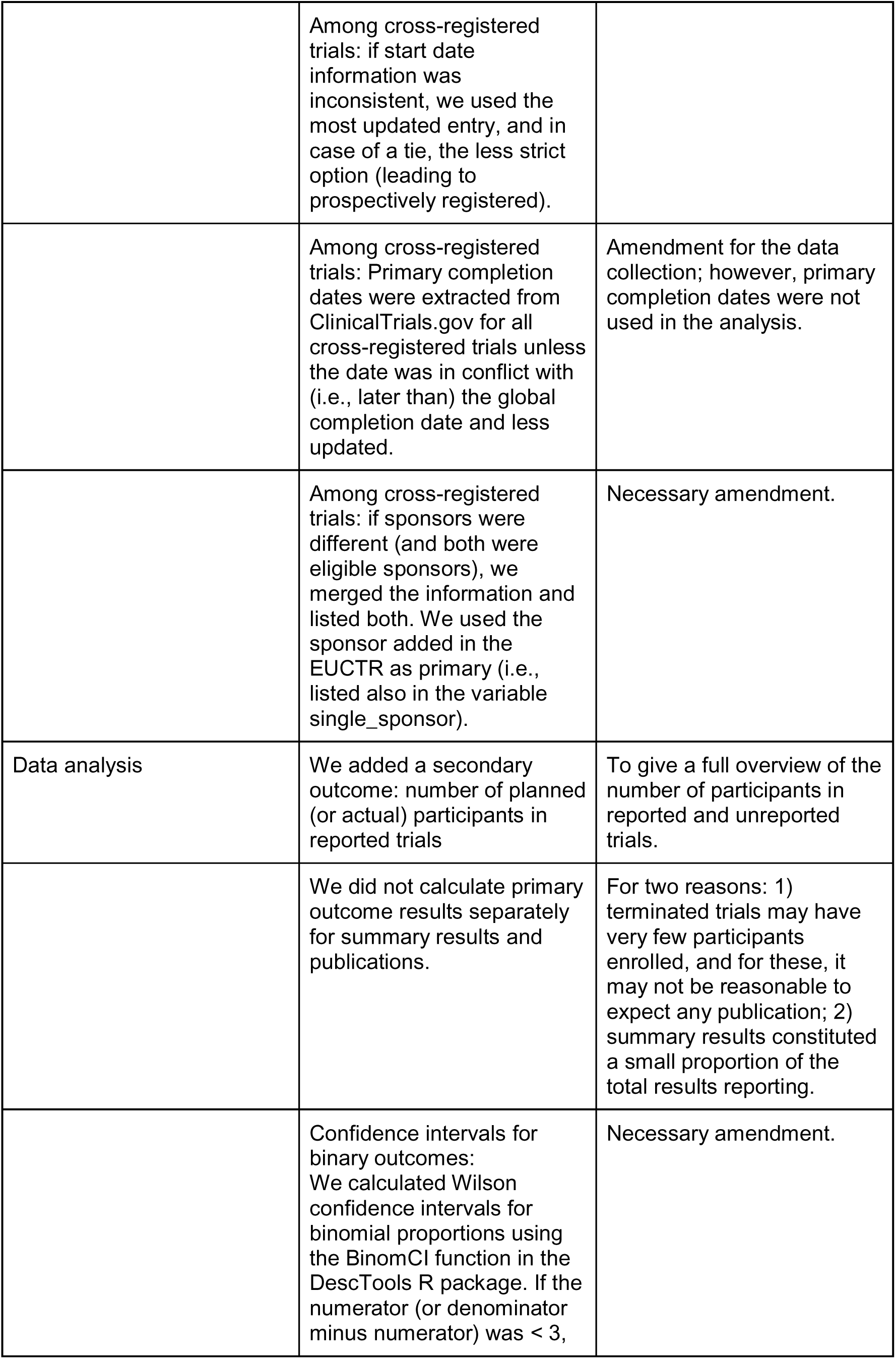
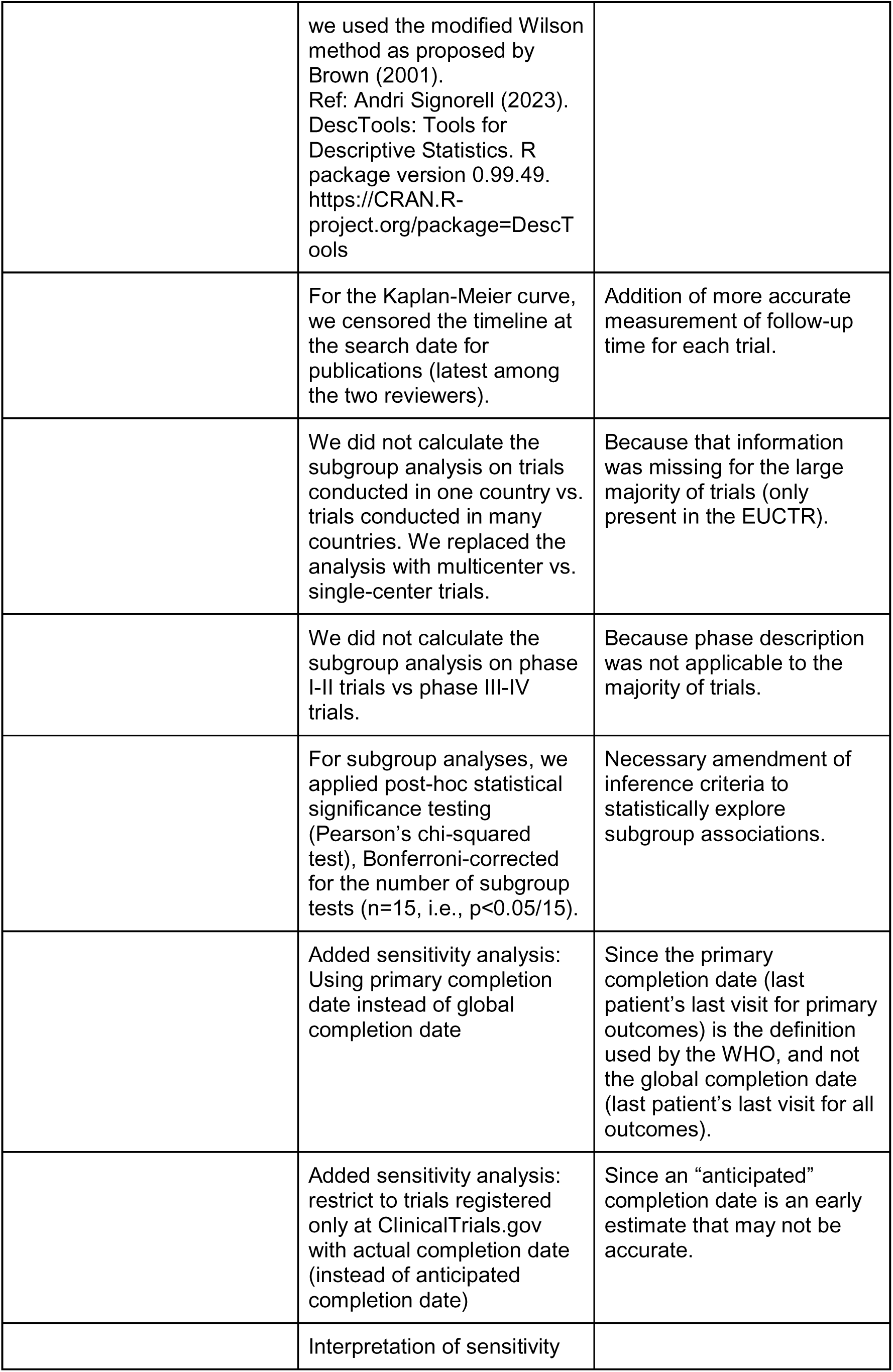
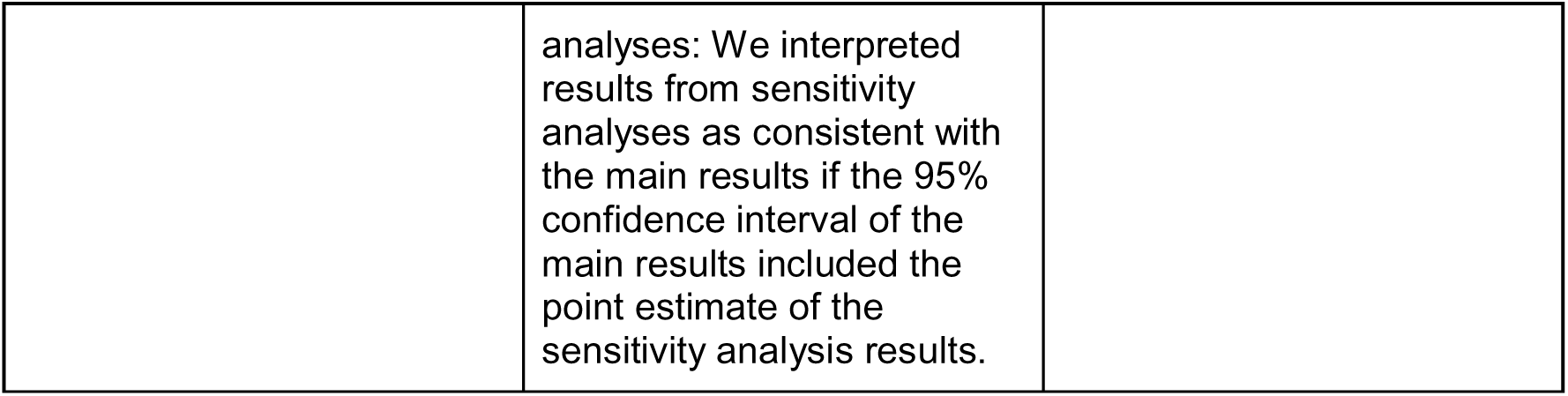
Amendments to the study protocol.

**Table 2.**
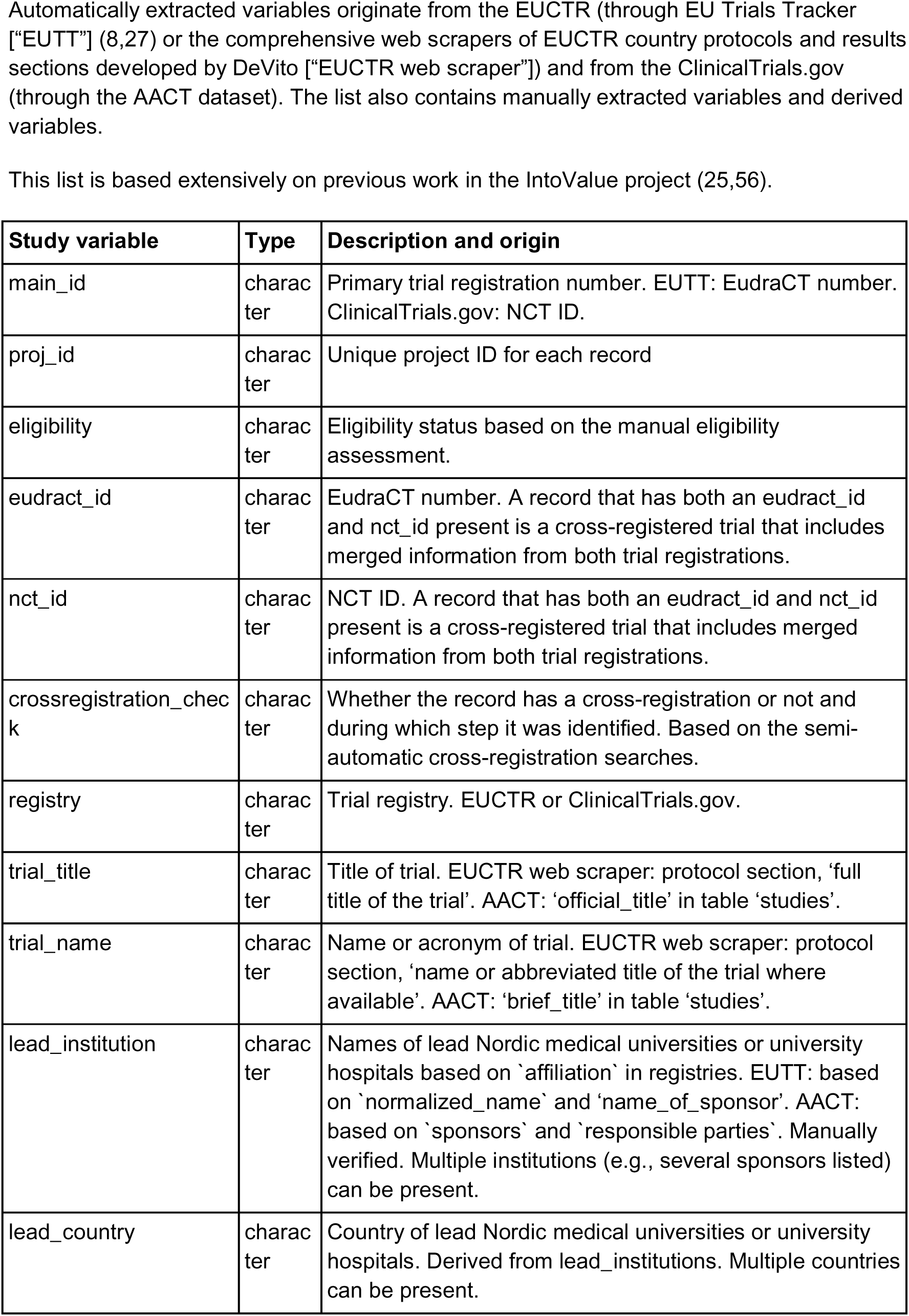
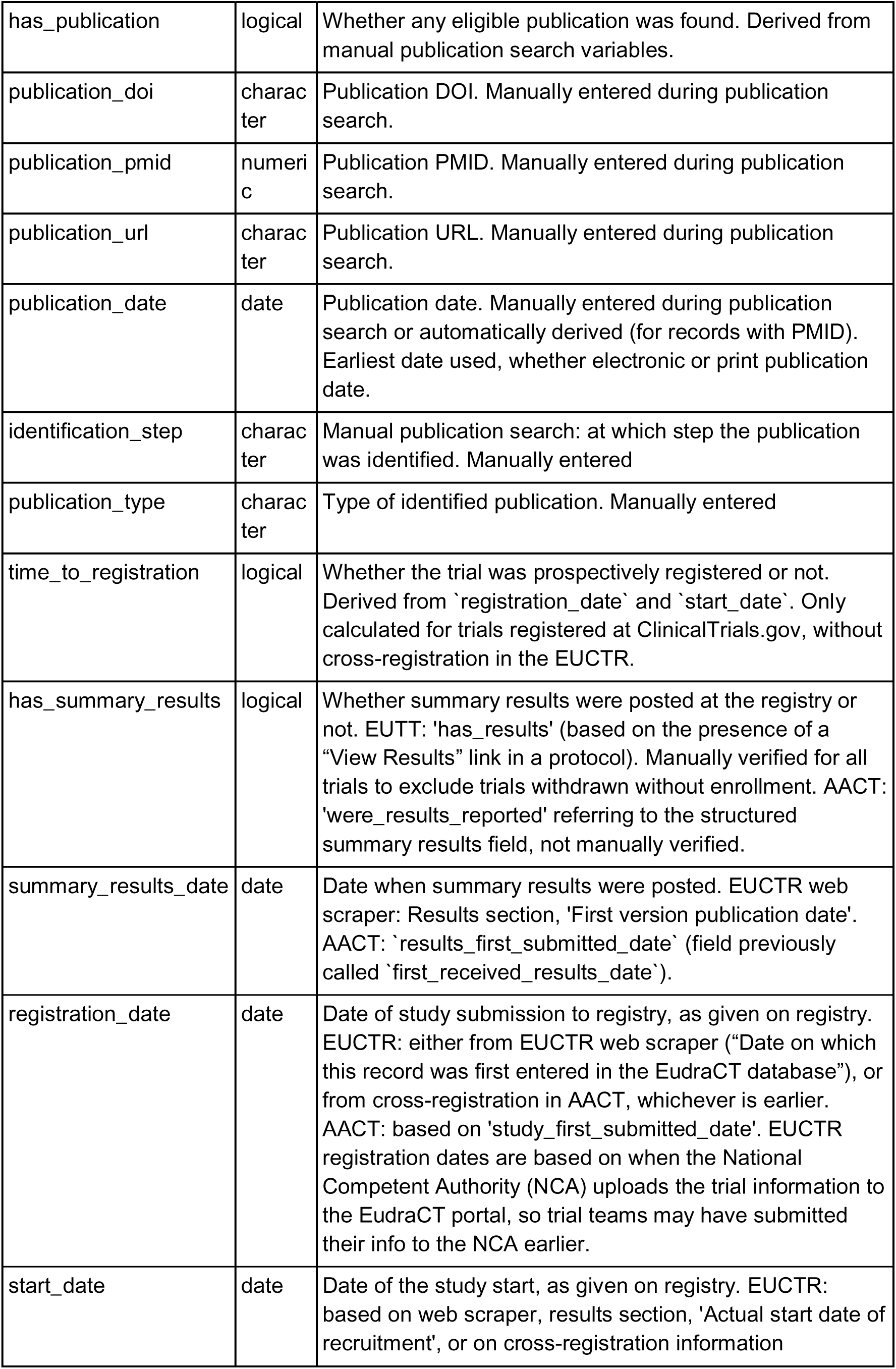
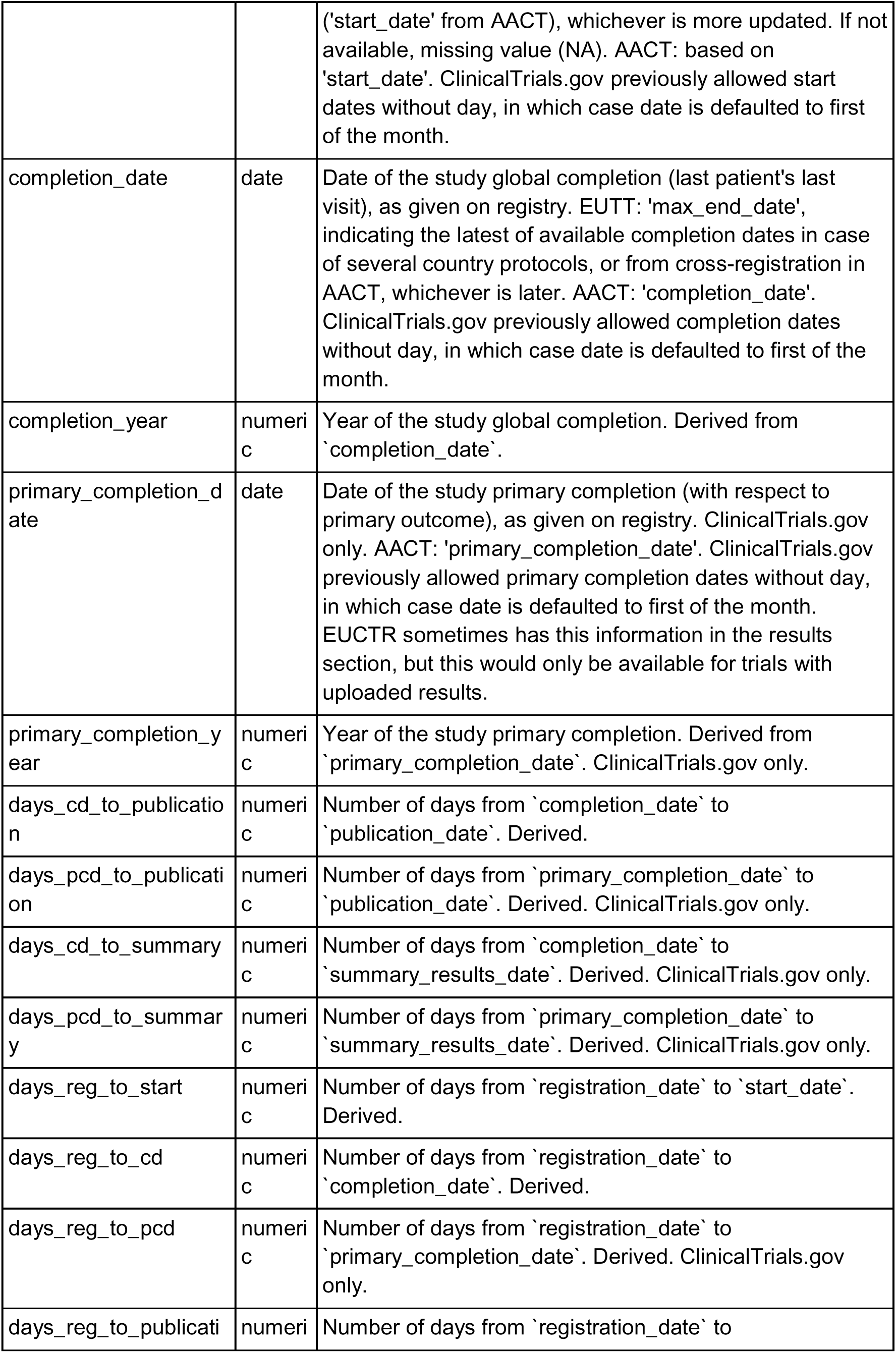
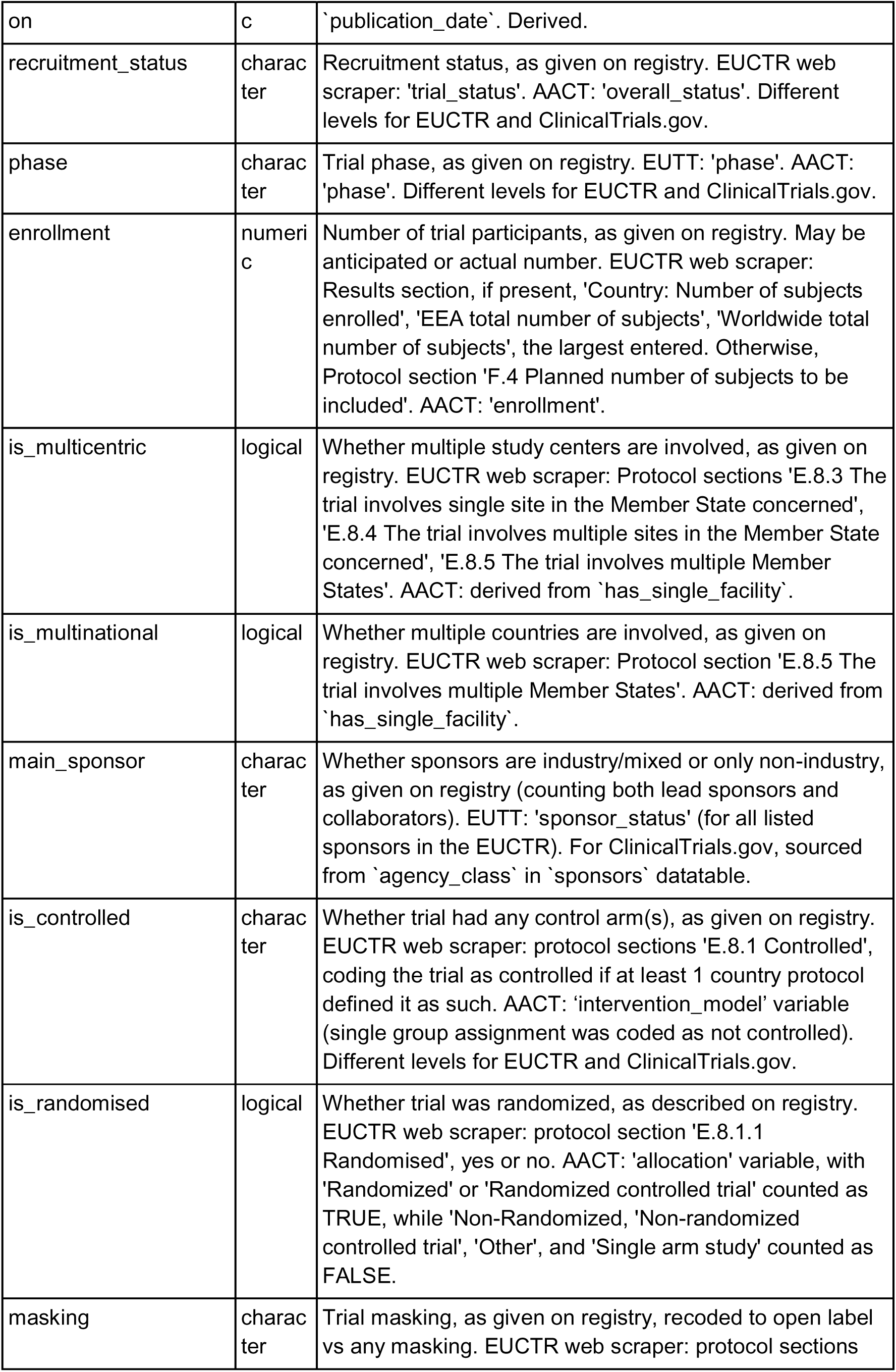
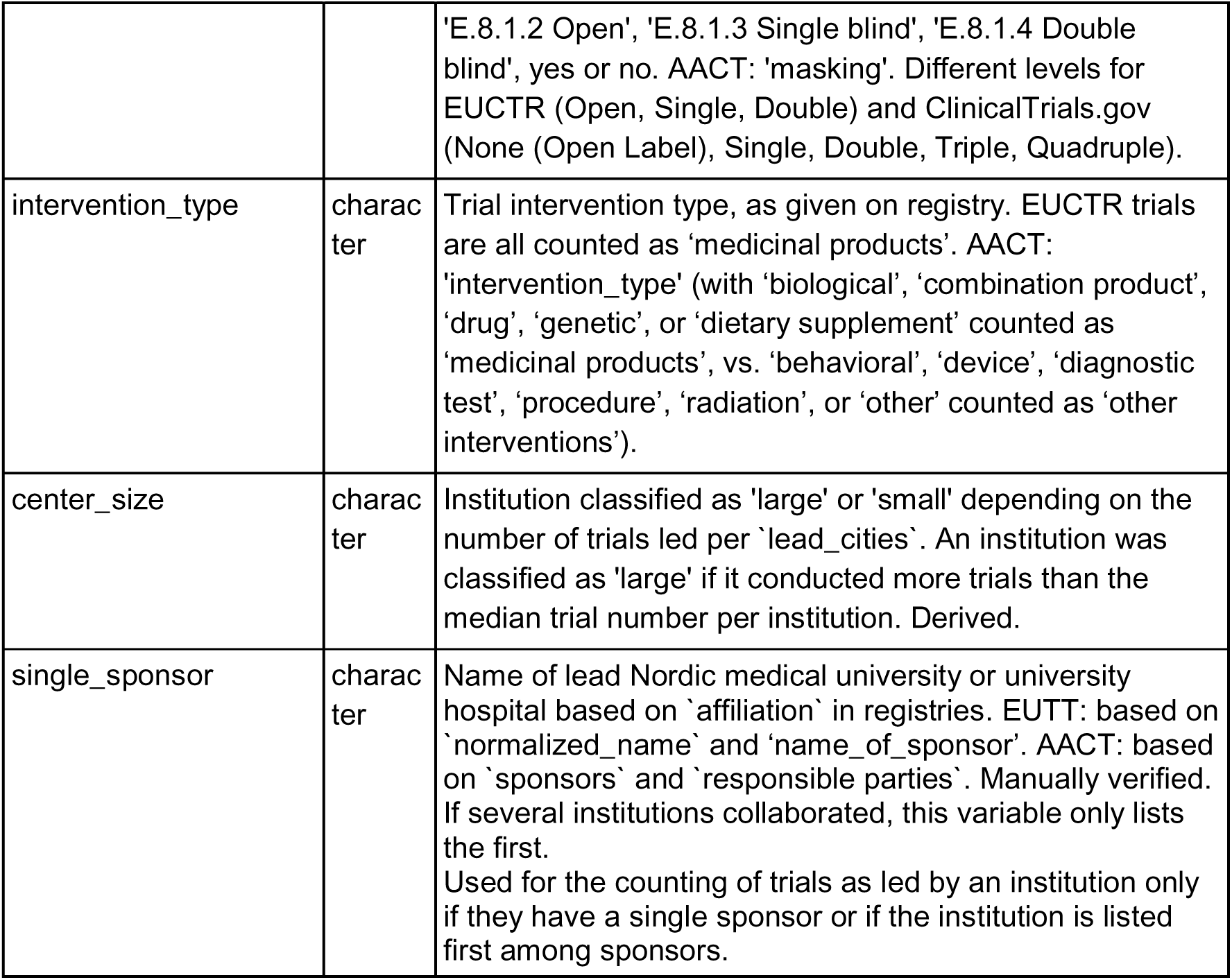
Study variables.

**Table 3.**
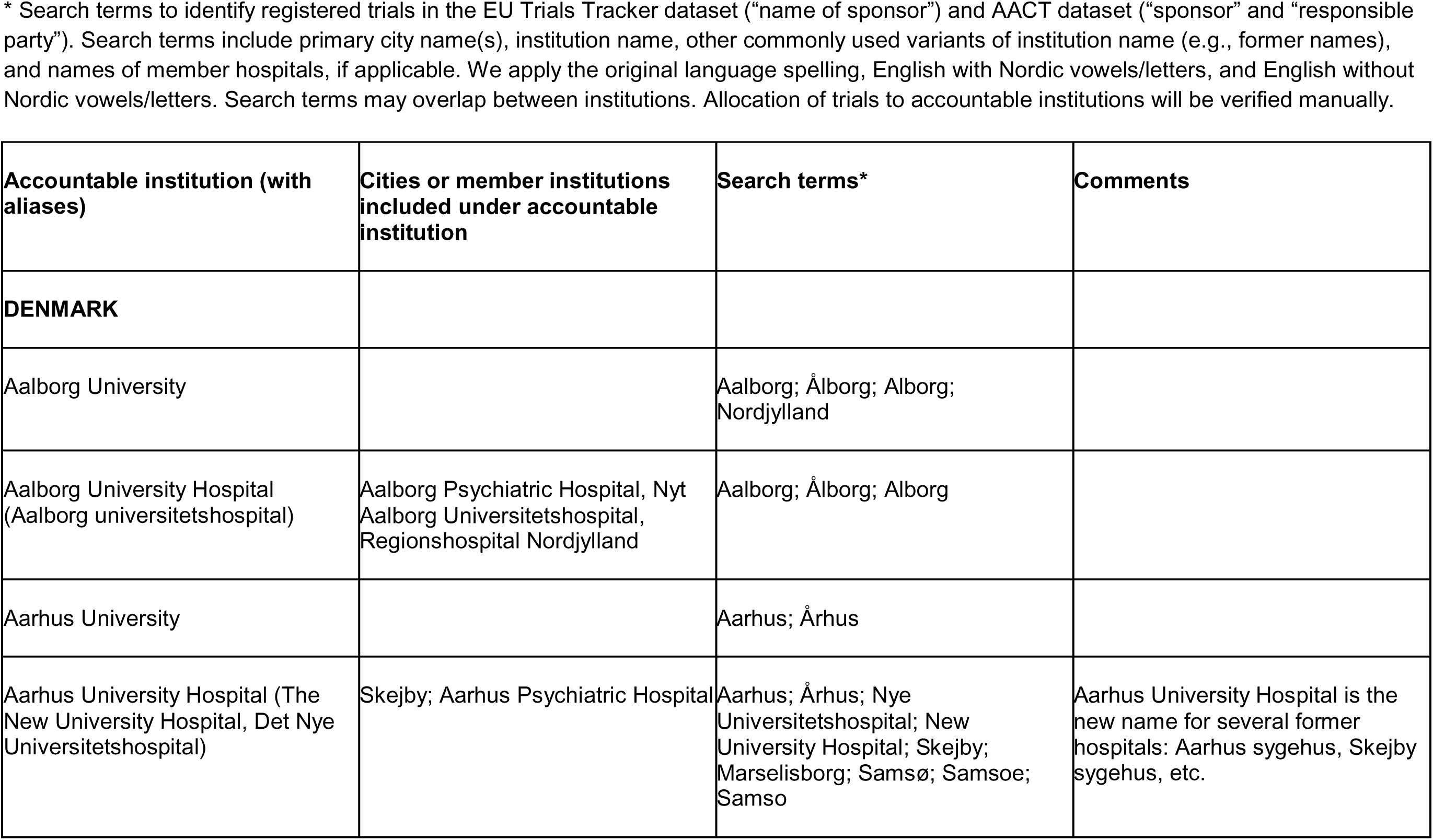
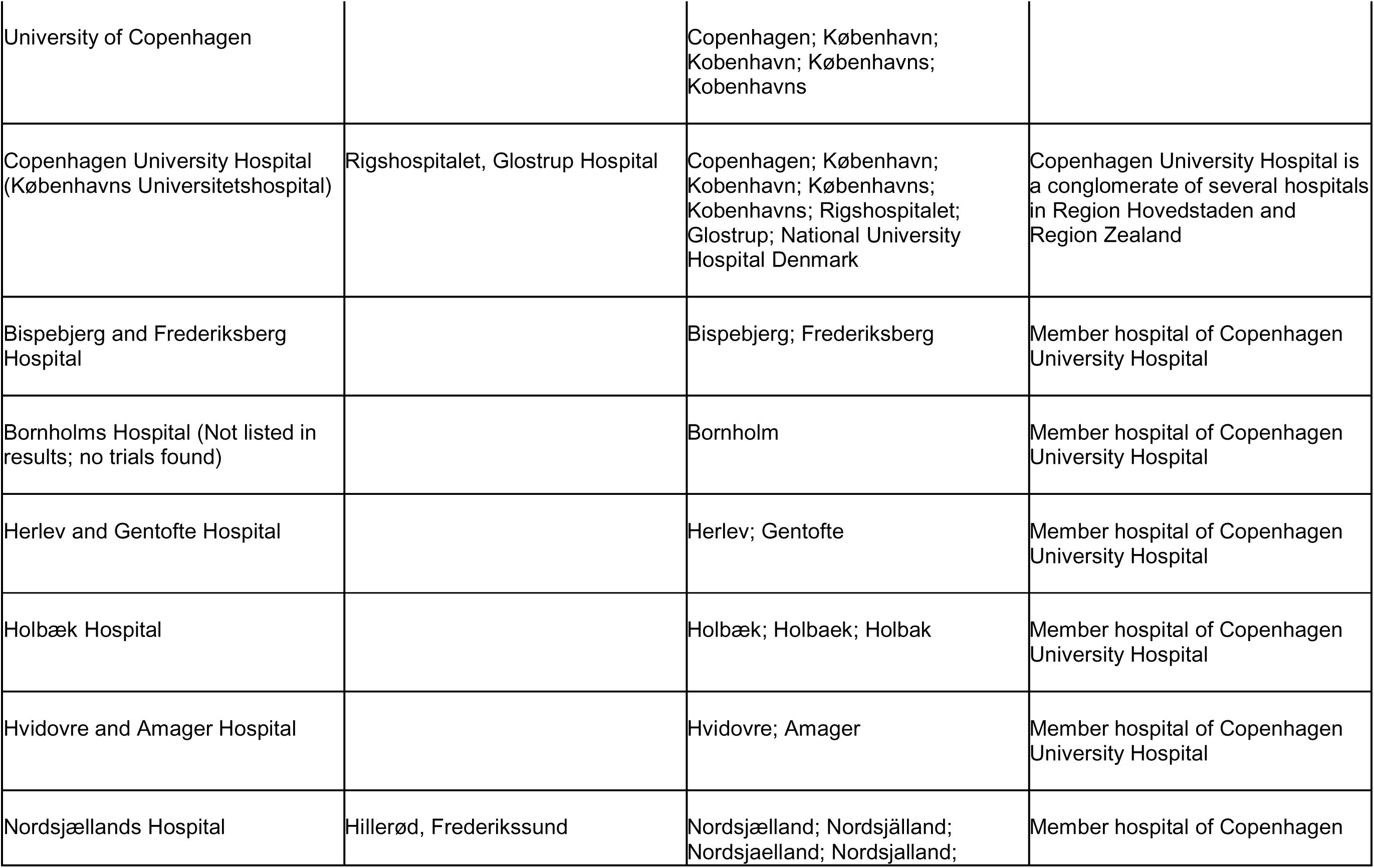
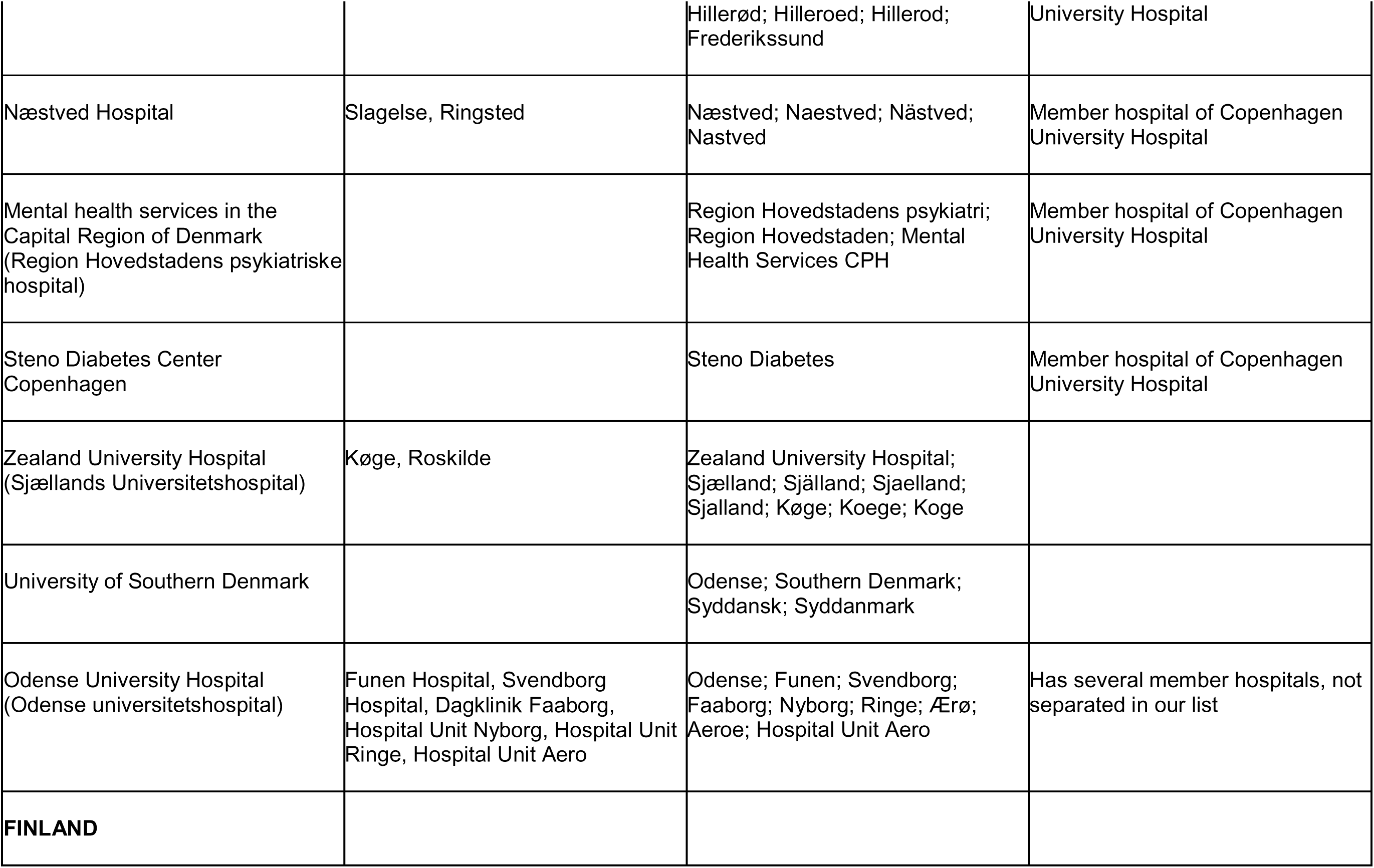
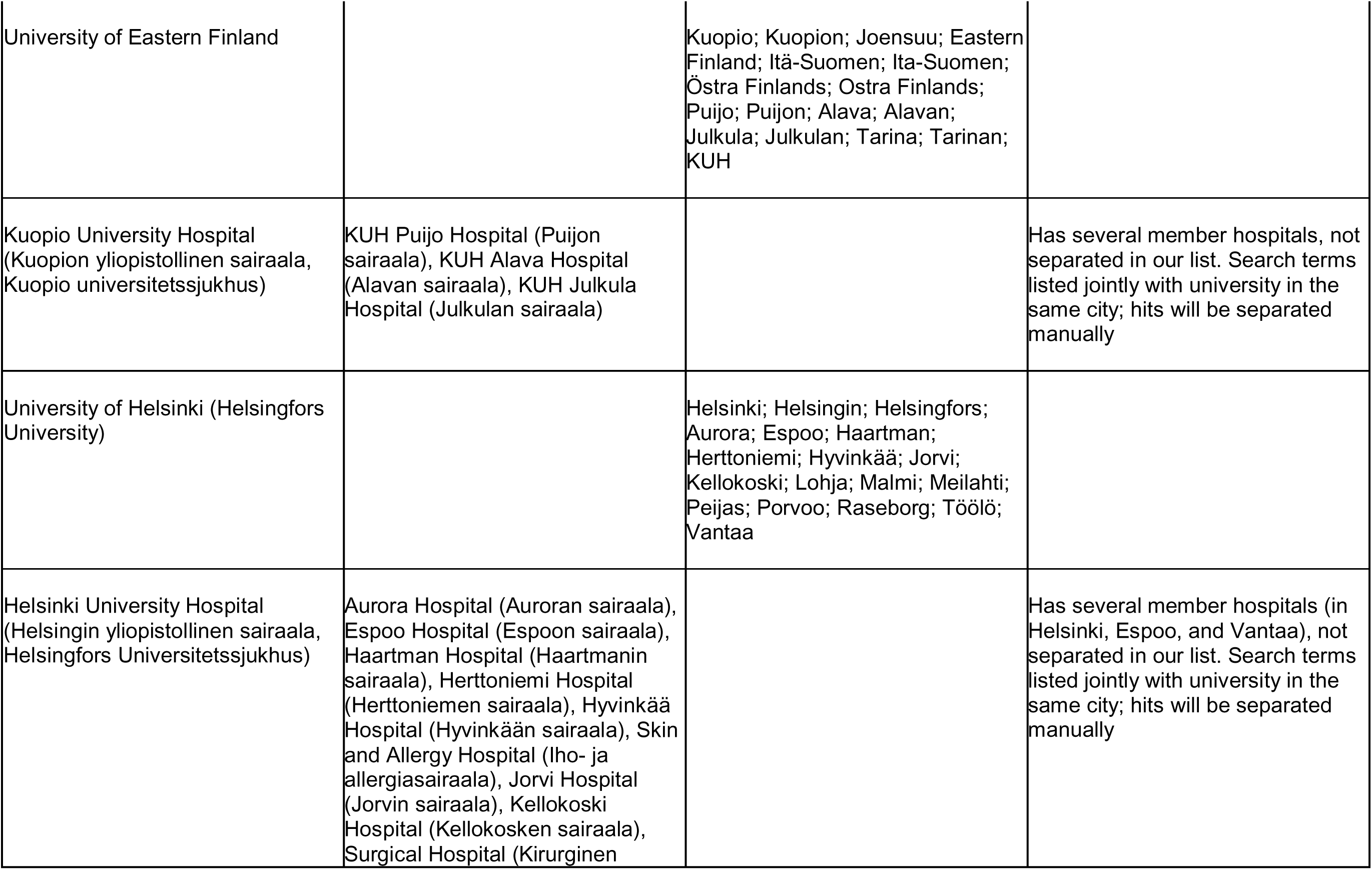
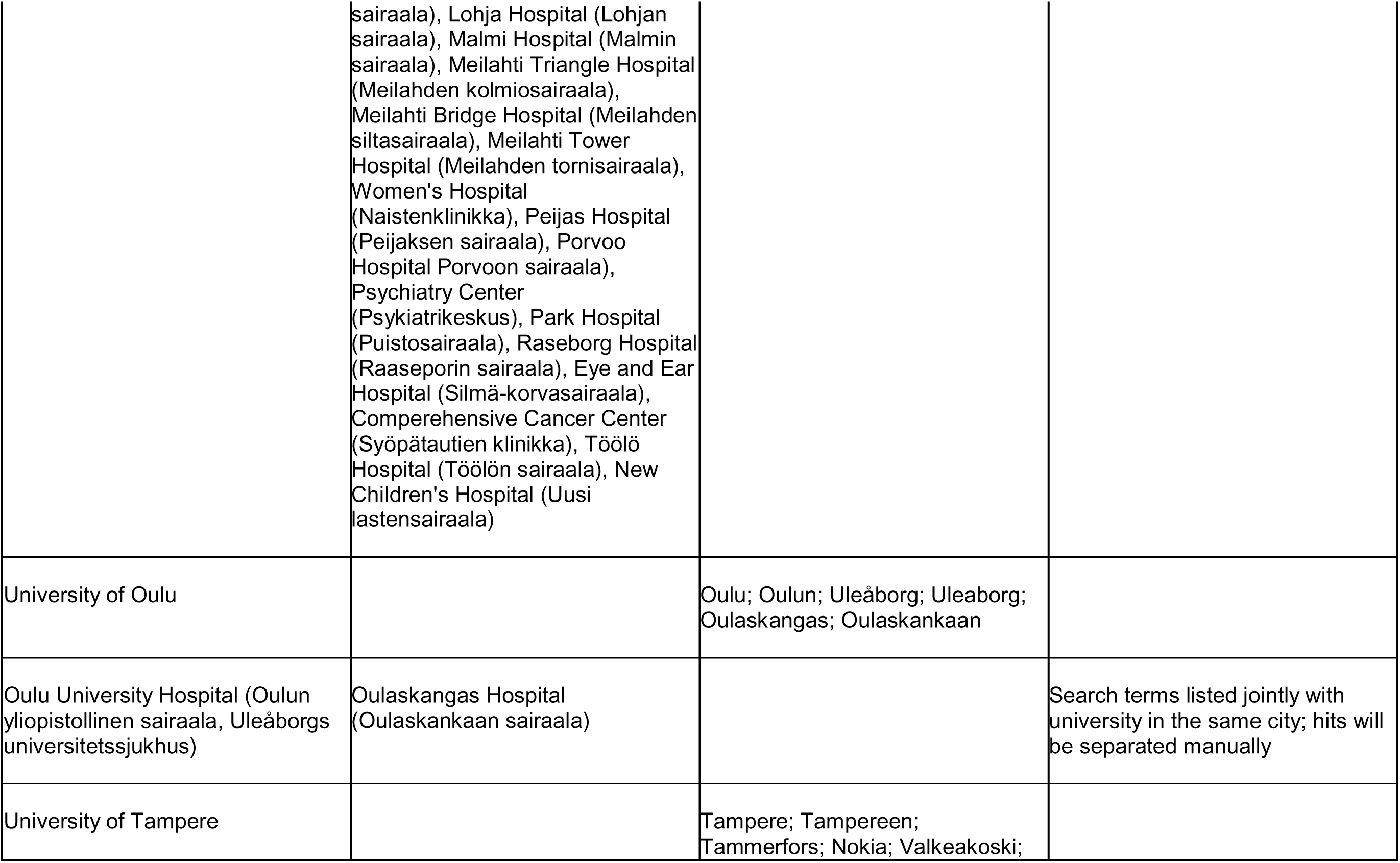
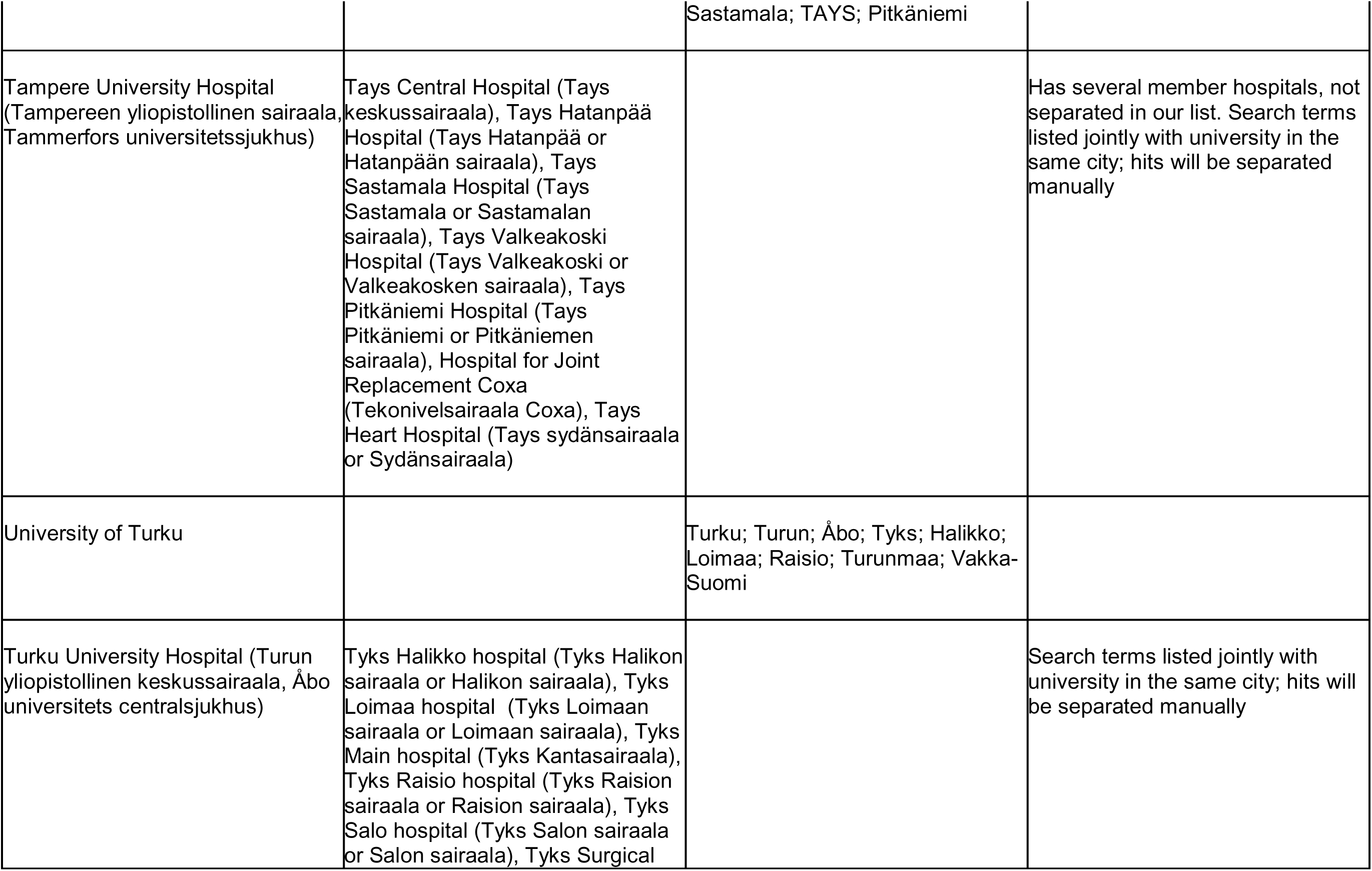
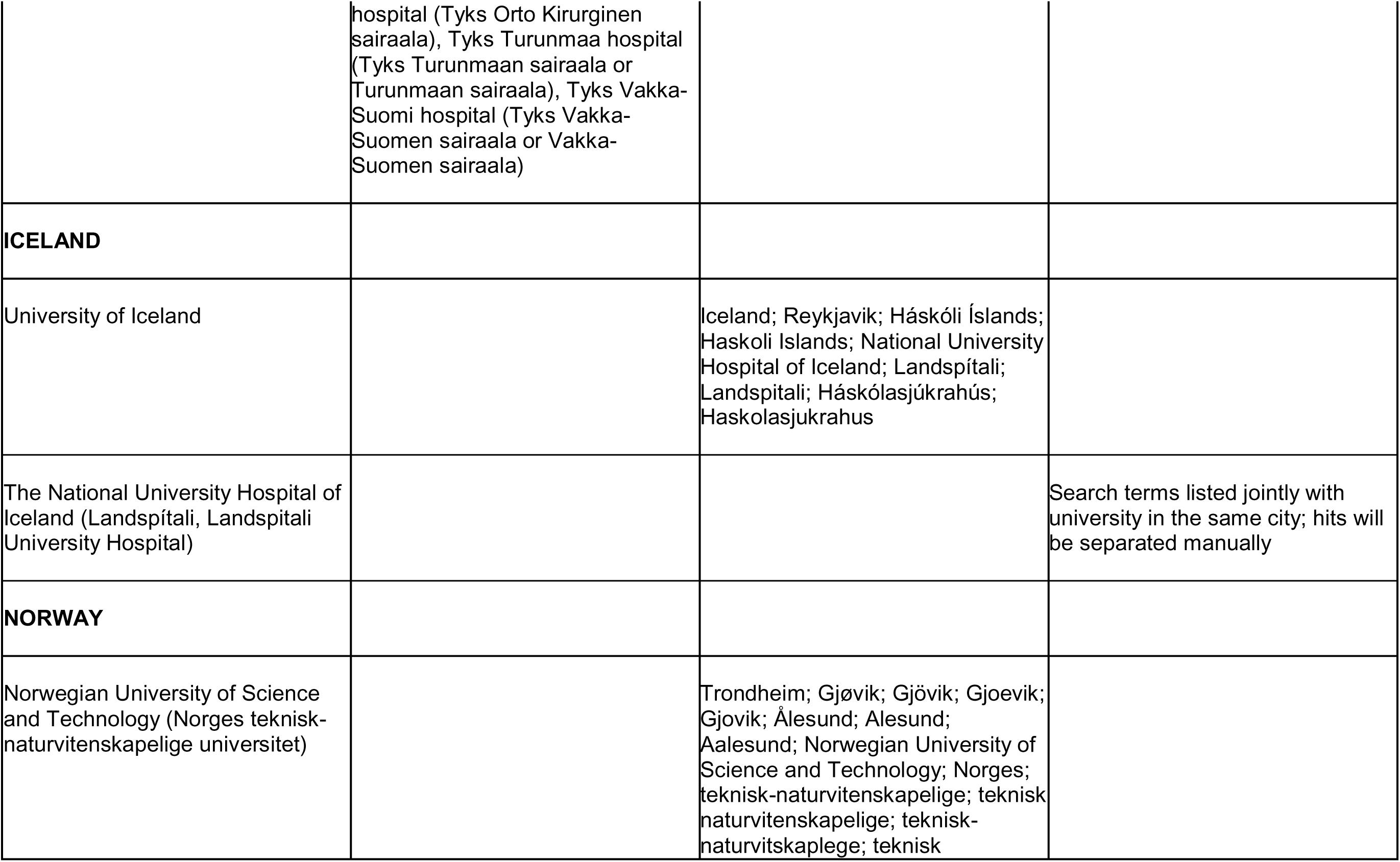
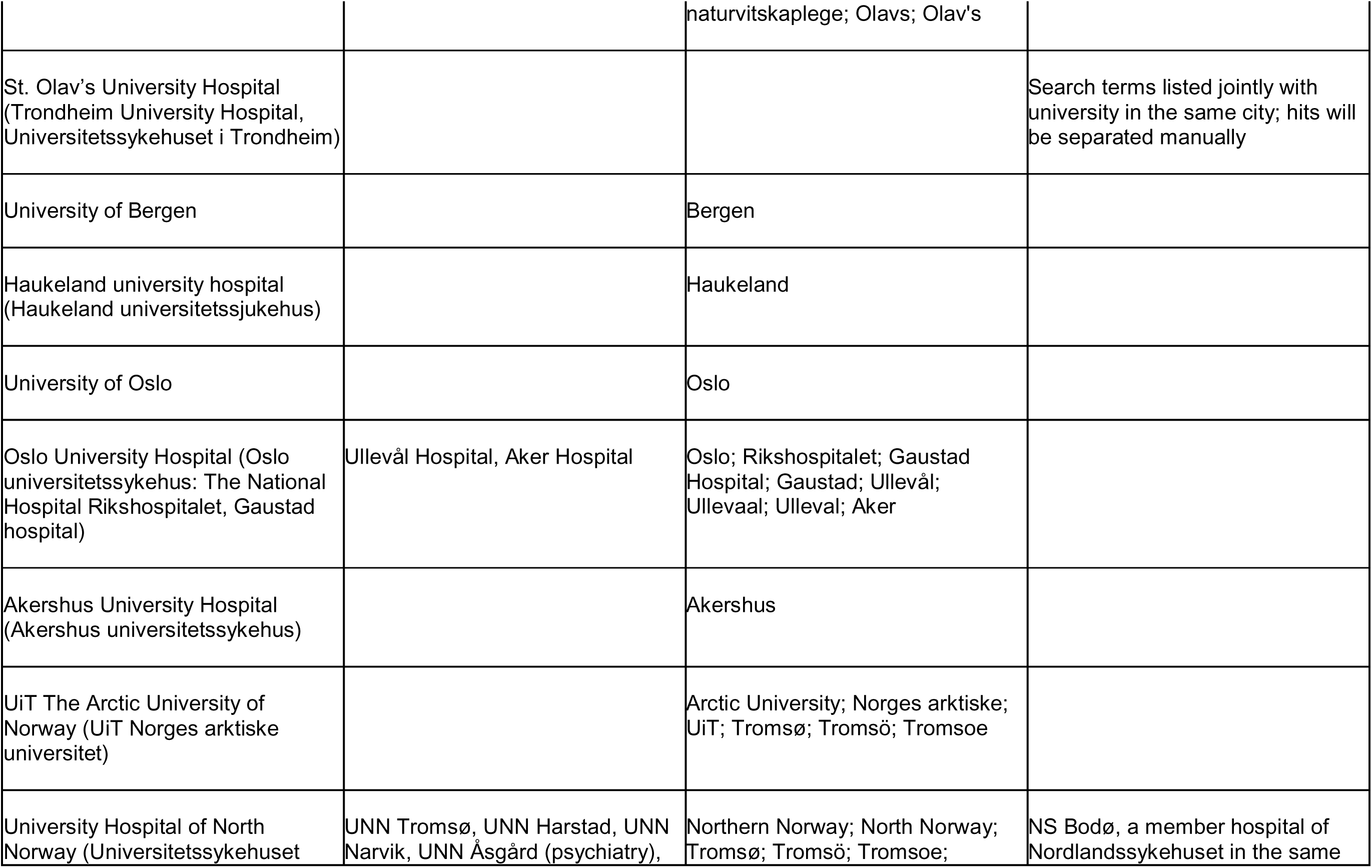
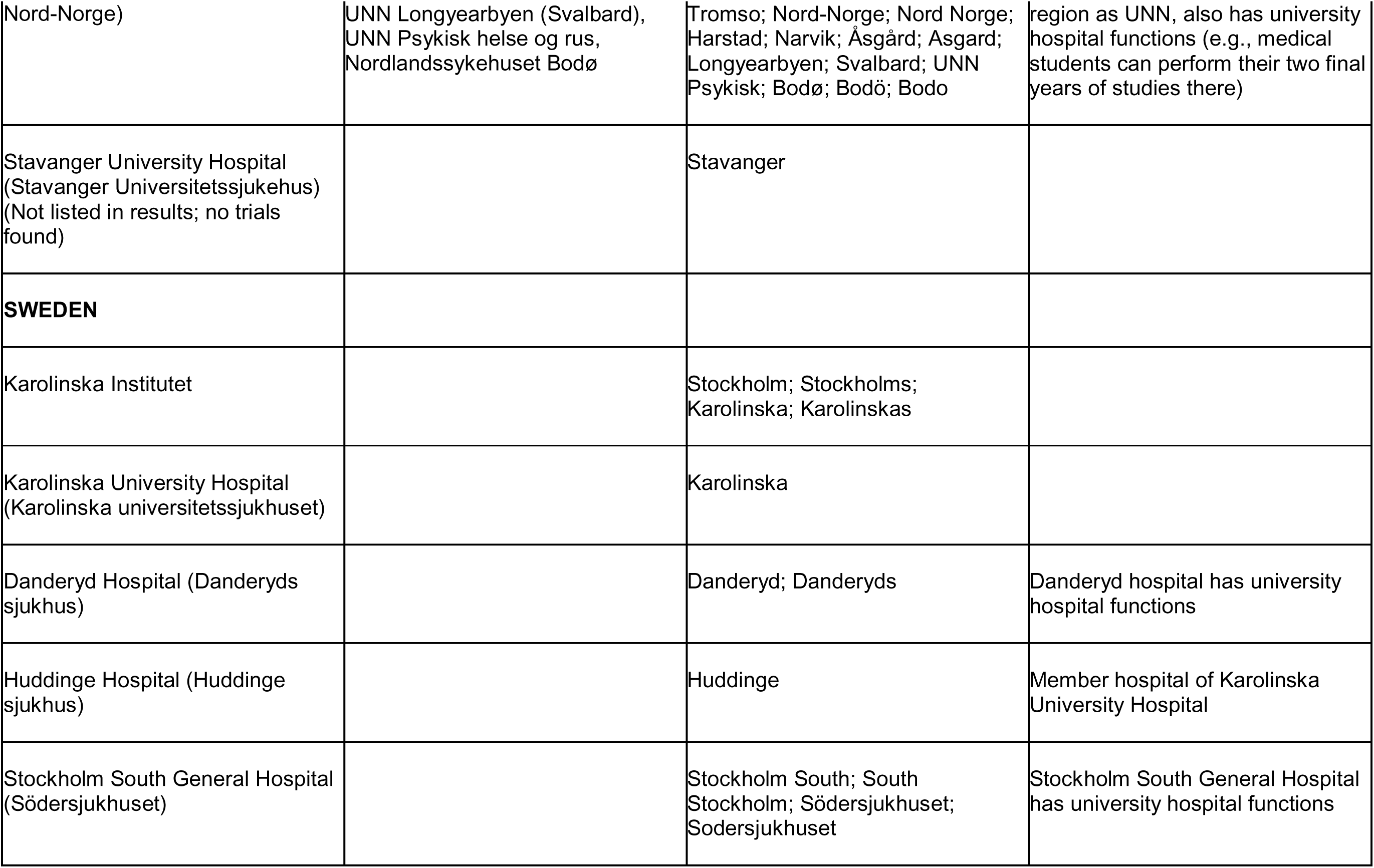
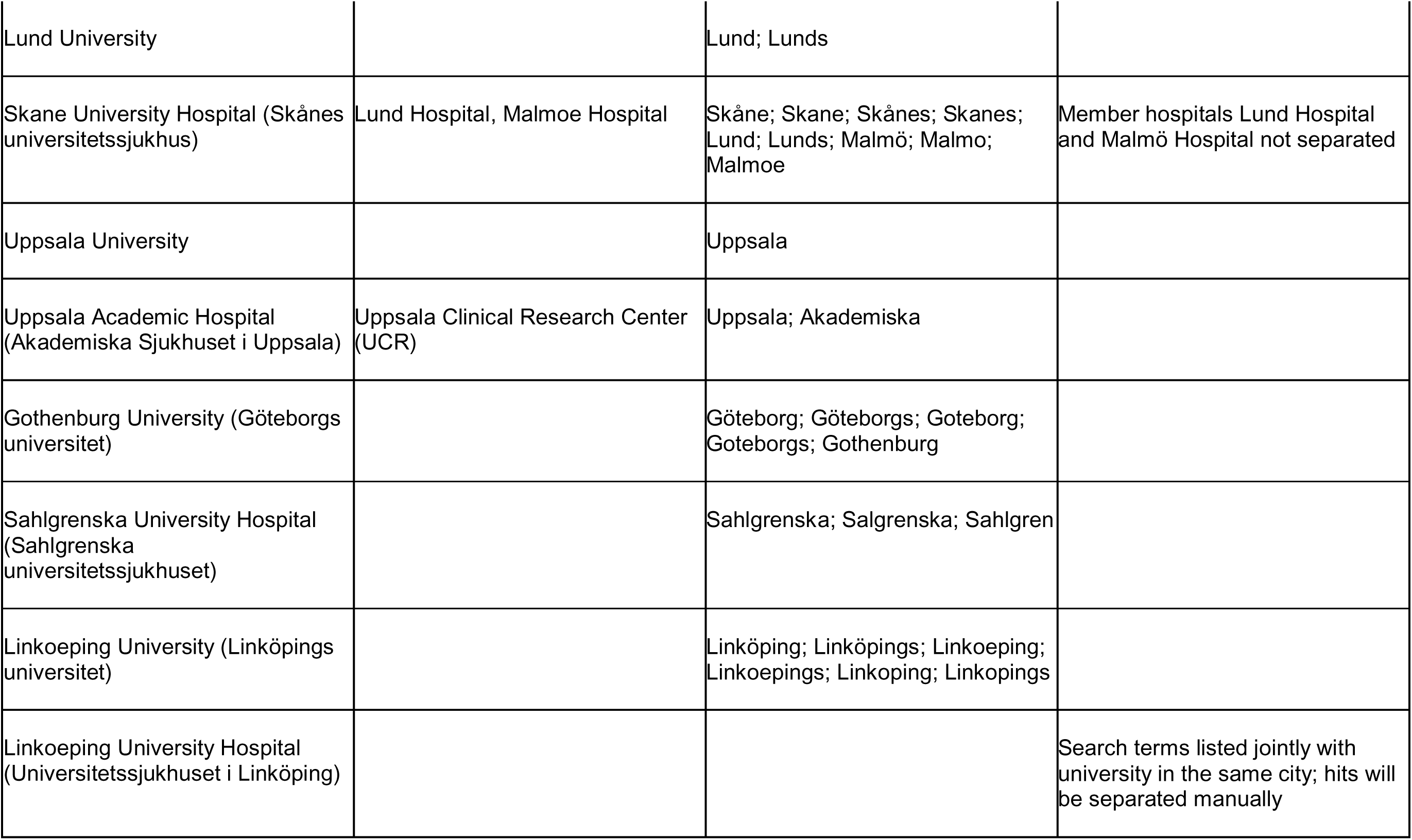
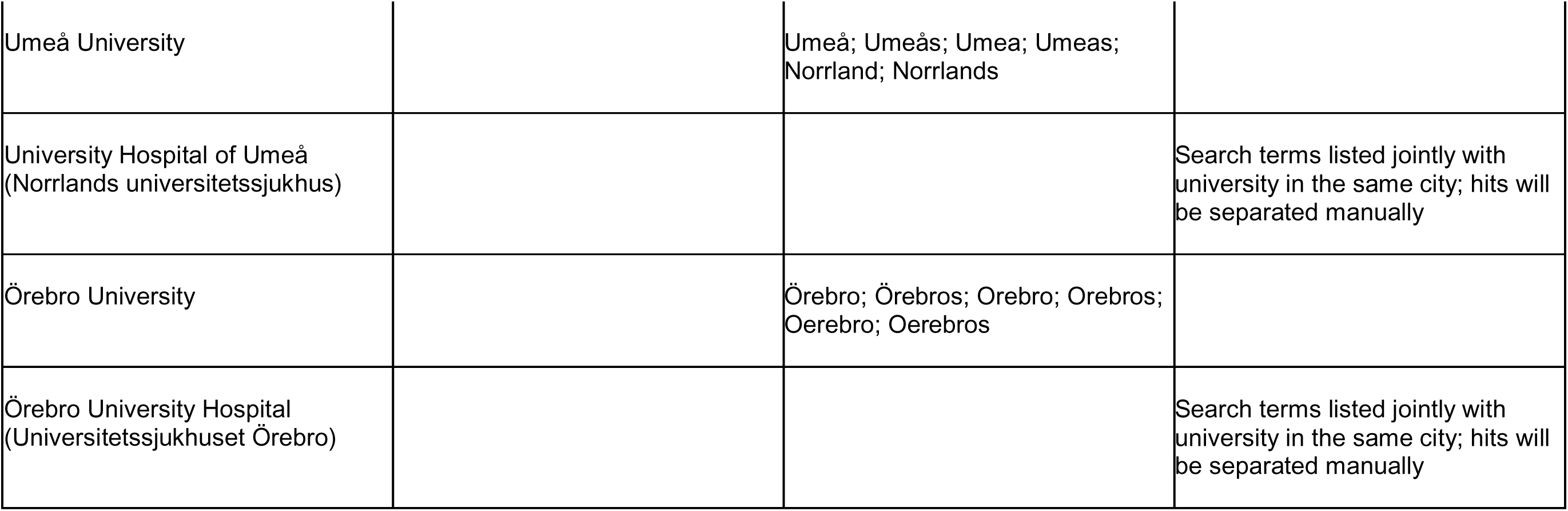
Medical universities and university hospitals in the Nordic countries (and search terms for trial registries)

**Table 4.**
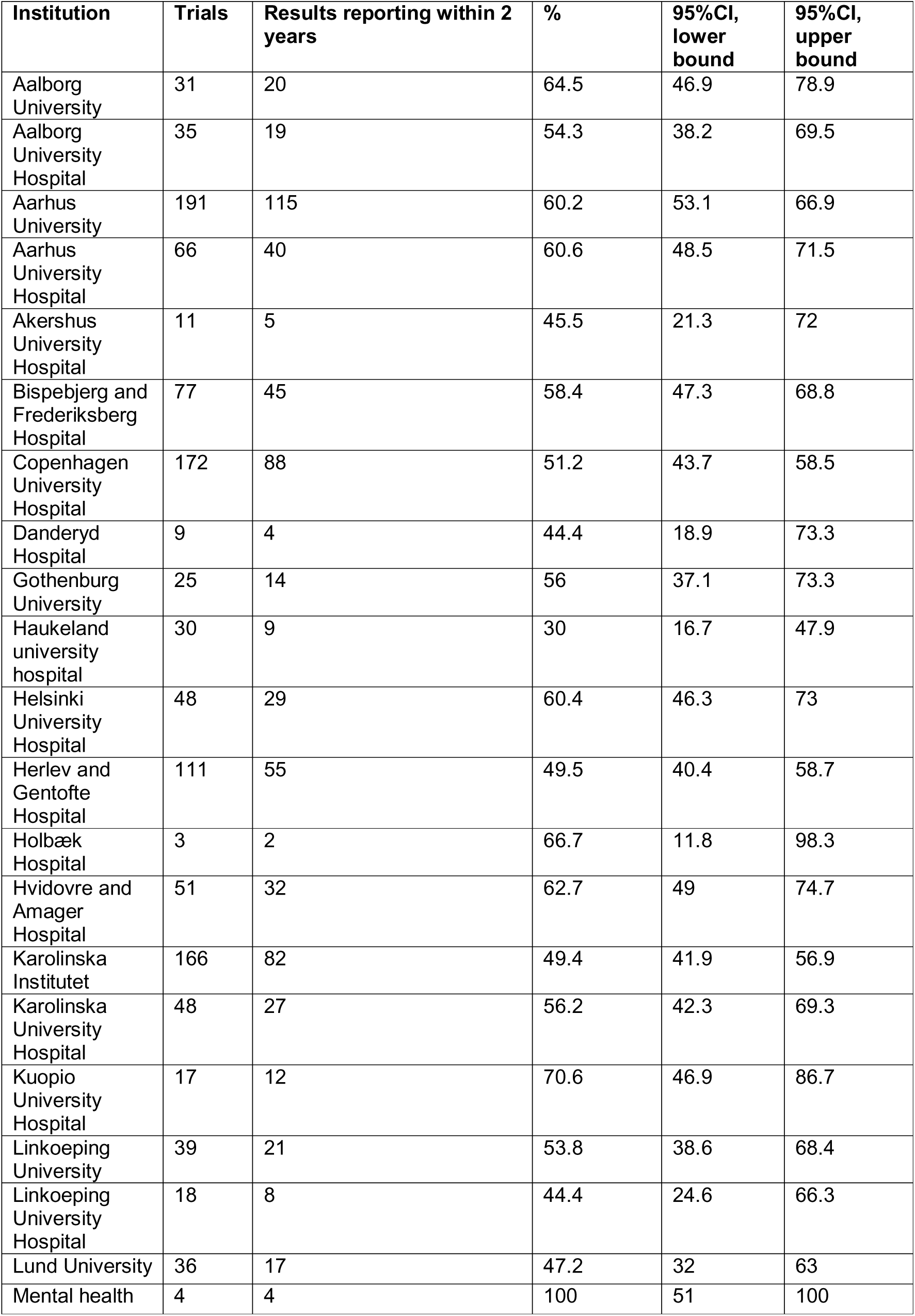
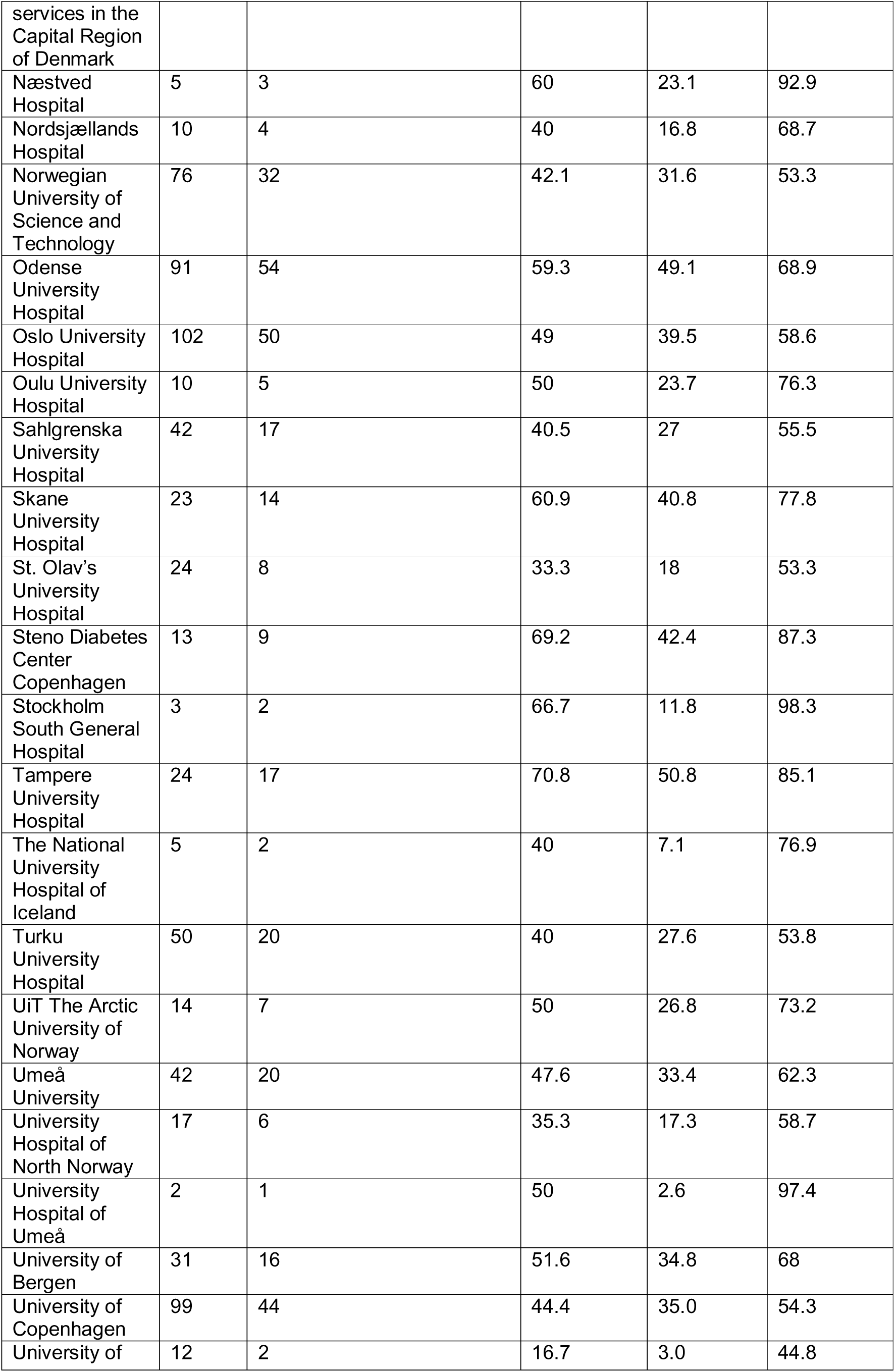
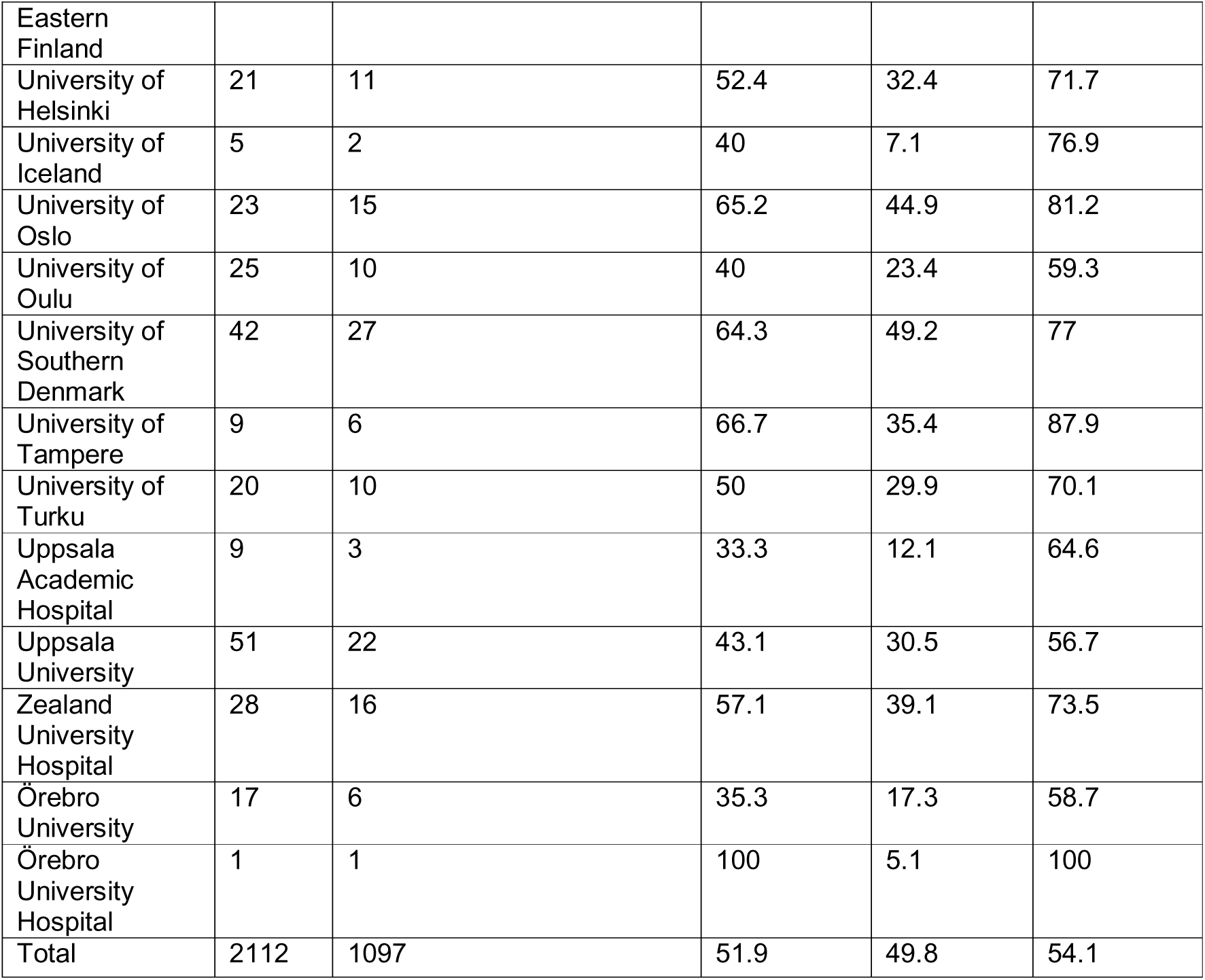
Share of trials with summary results or results publication within two years of completion, per institution.

**Table 5.**
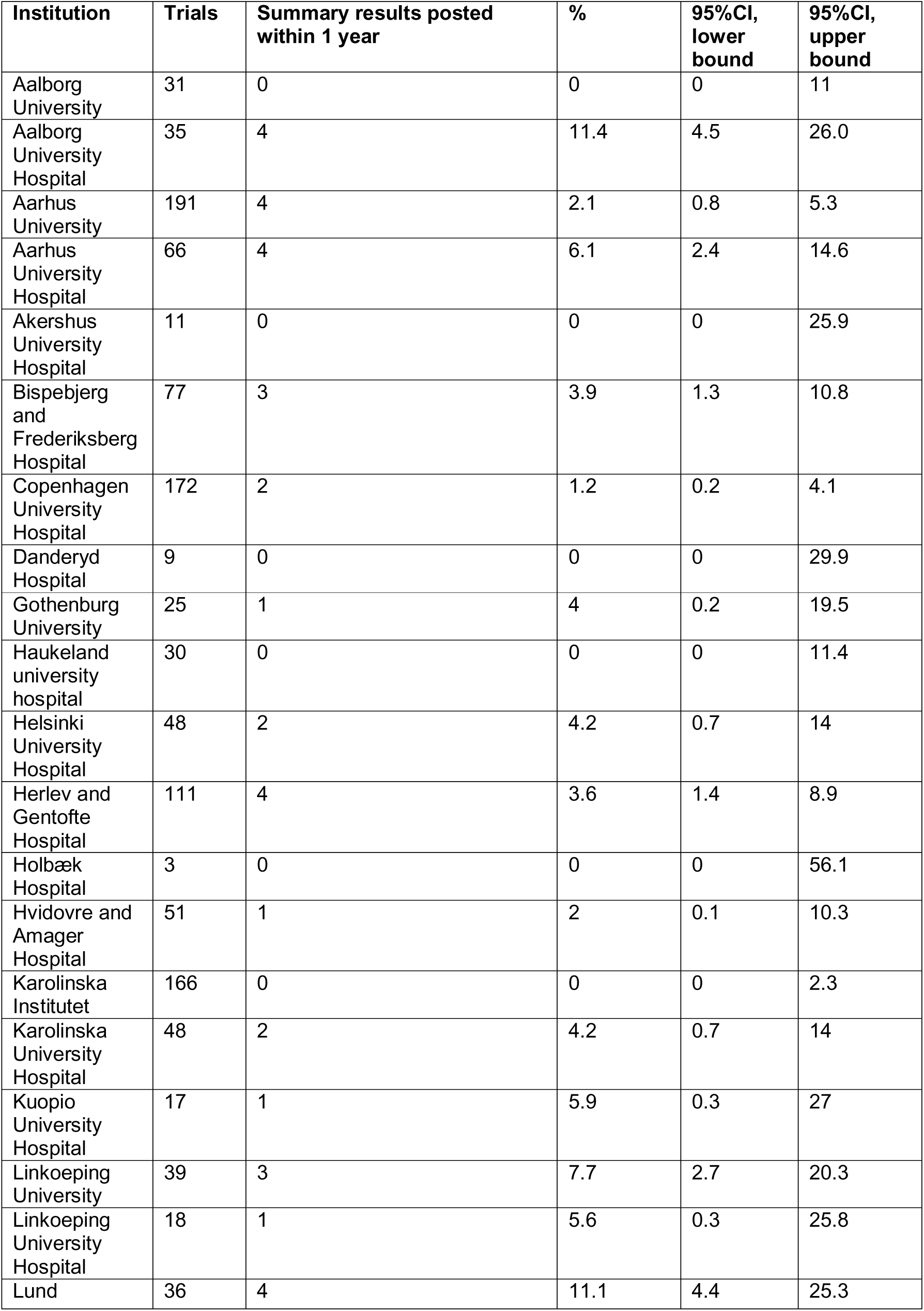
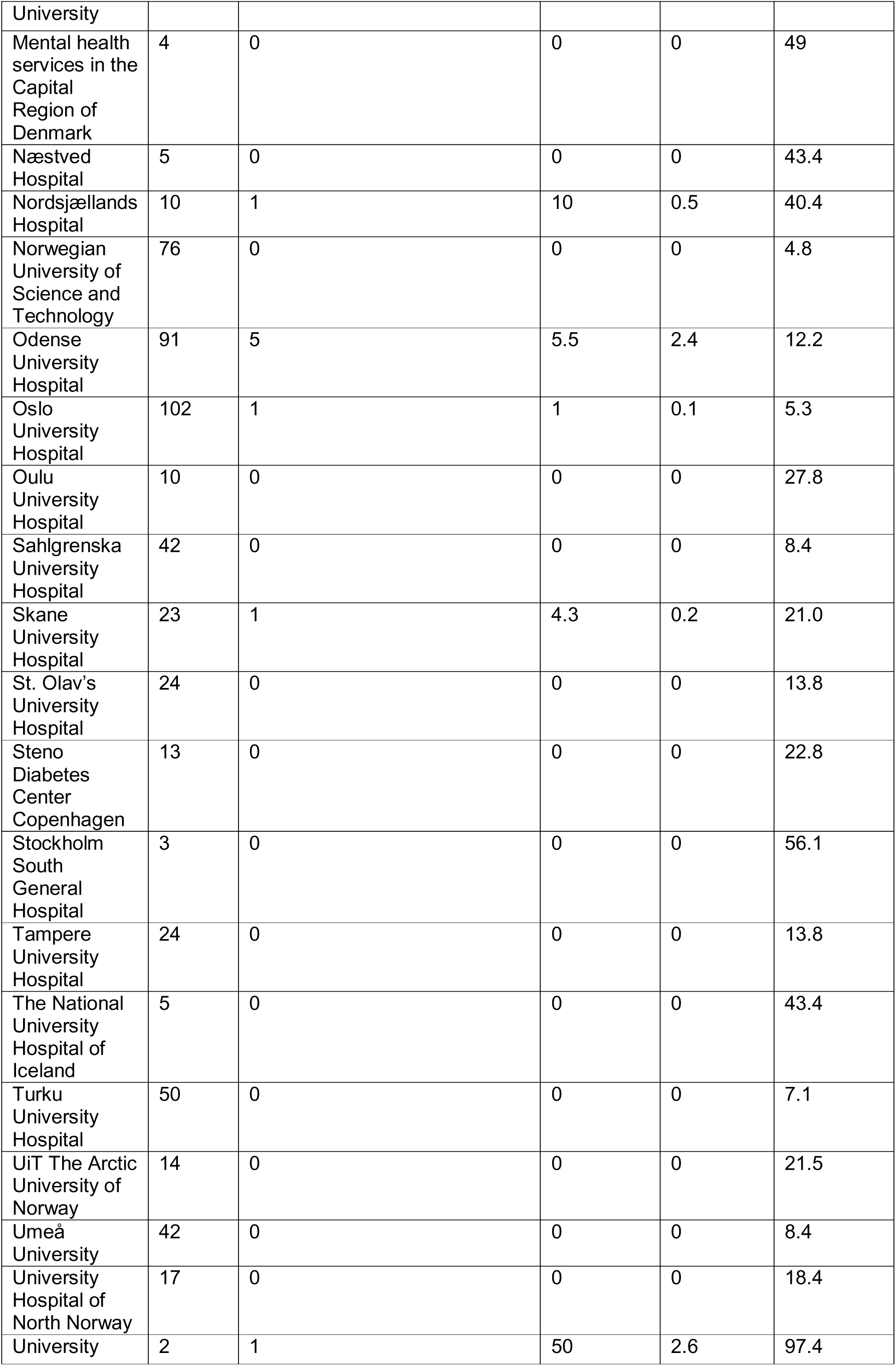
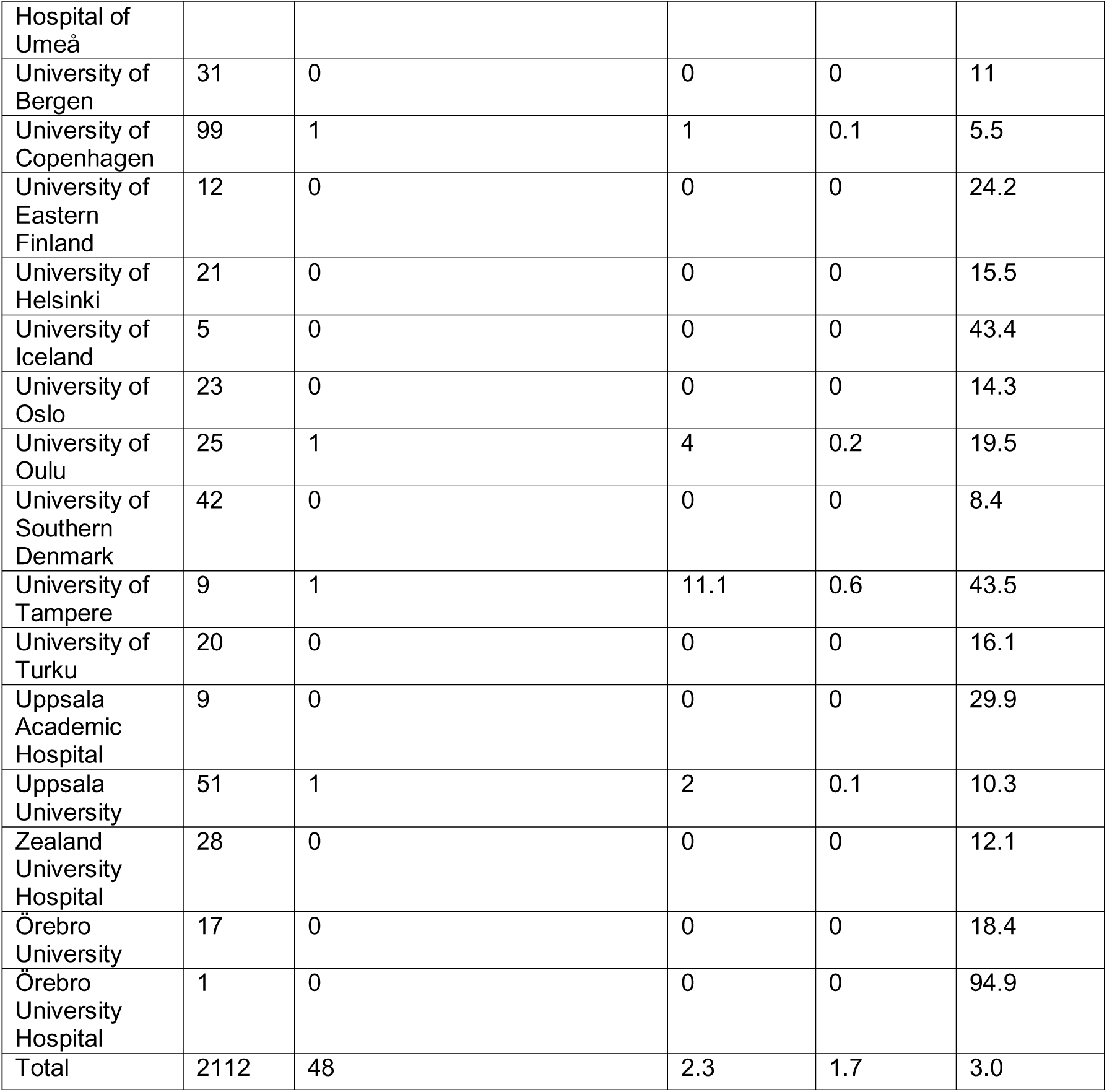
Share of trials with summary results within one year of completion, per institution.

**Table 6.**
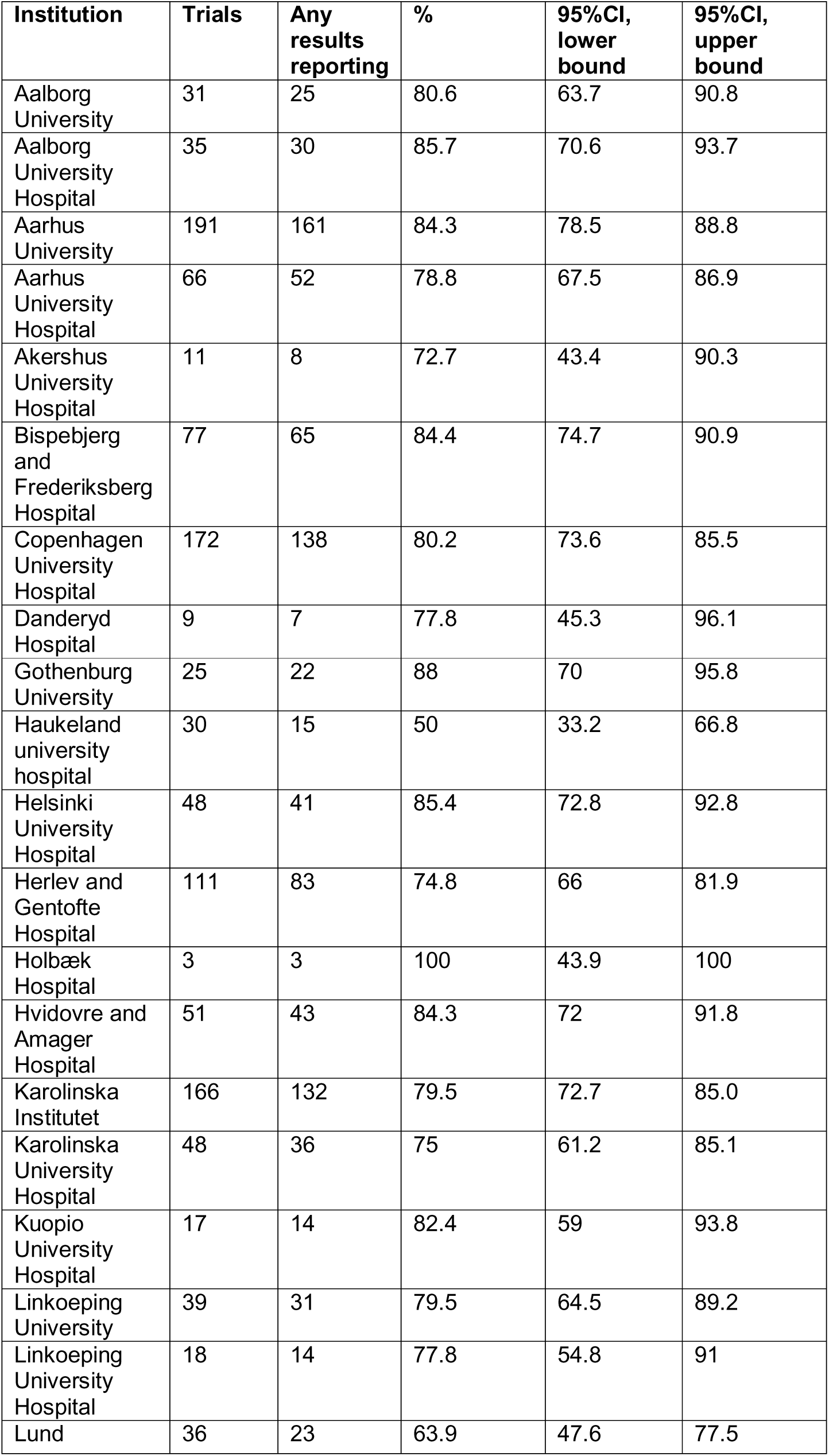
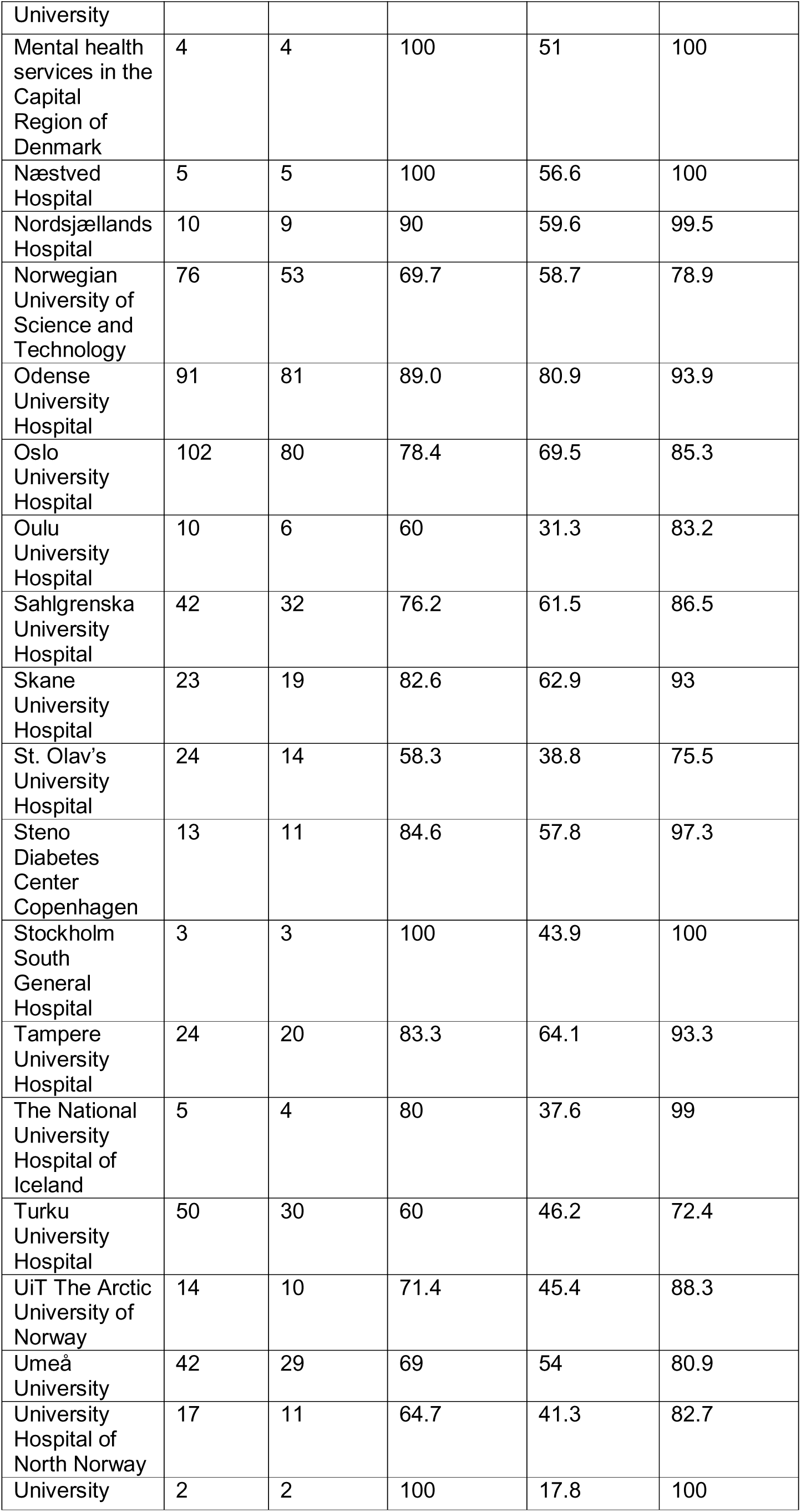
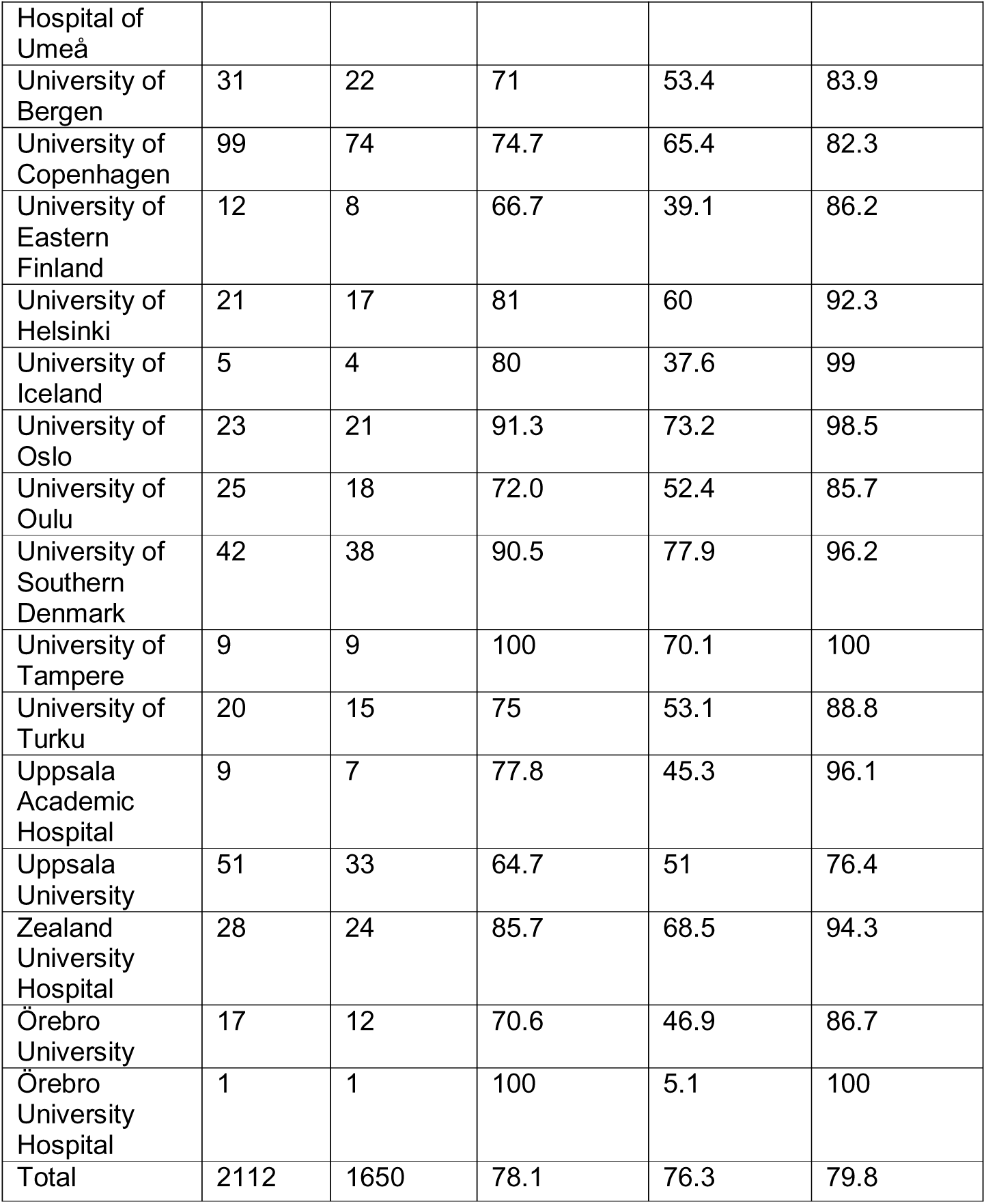
Share of trials with any results reporting (summary results or results publications) at the end of follow-up, per institution.

**Table 7.**
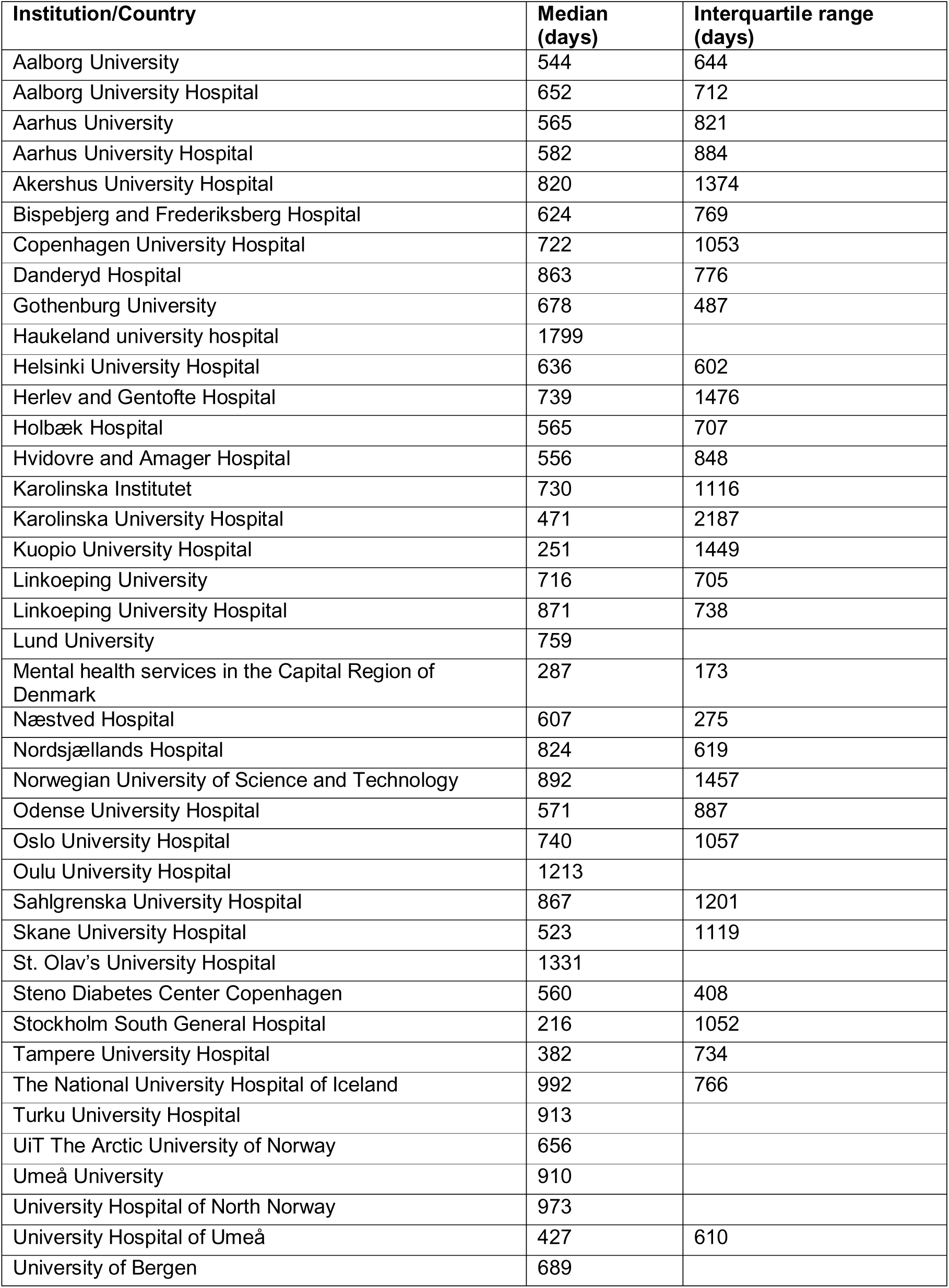
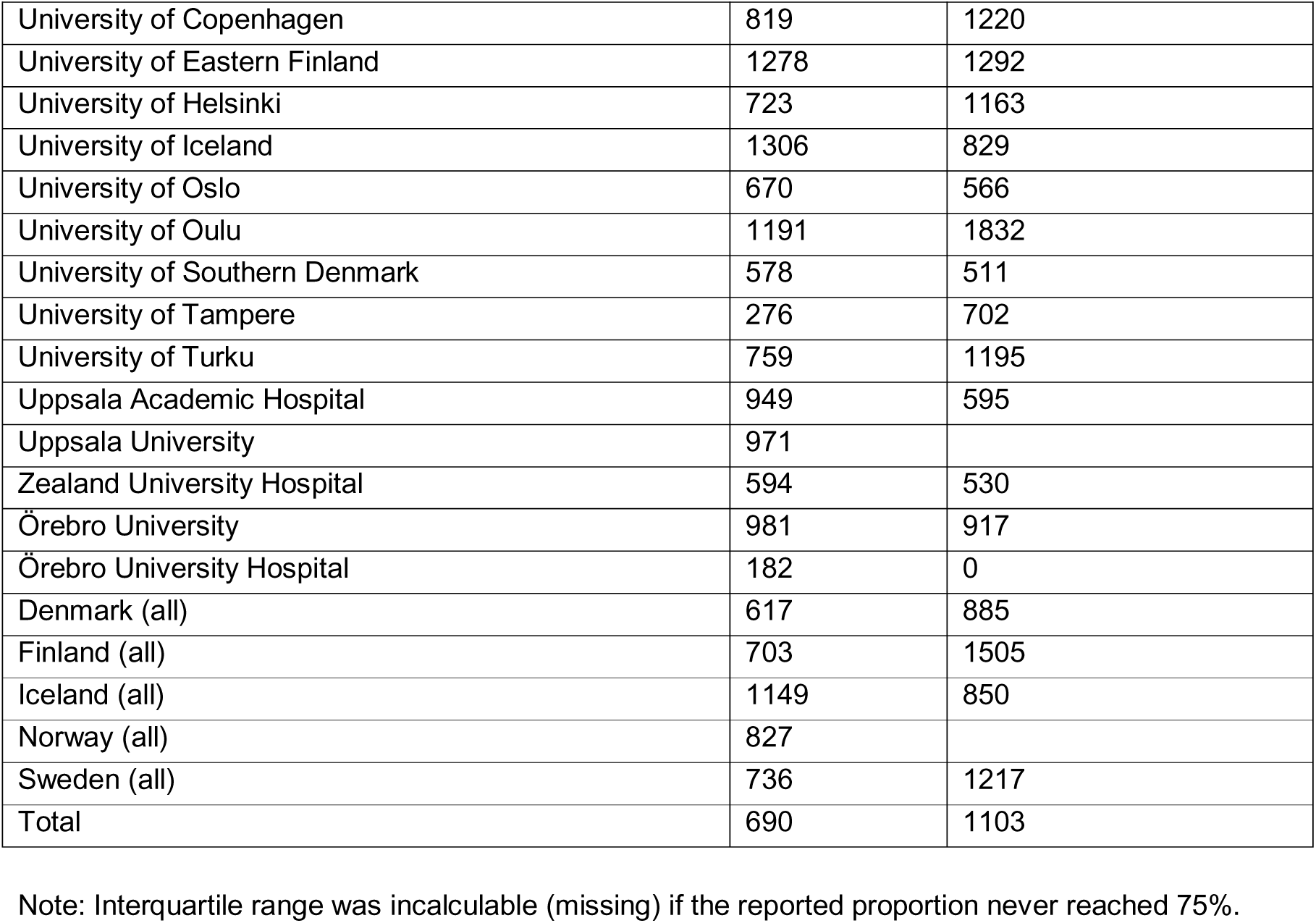
Median time from completion to the first reporting of results, per institution.

**Table 8.**
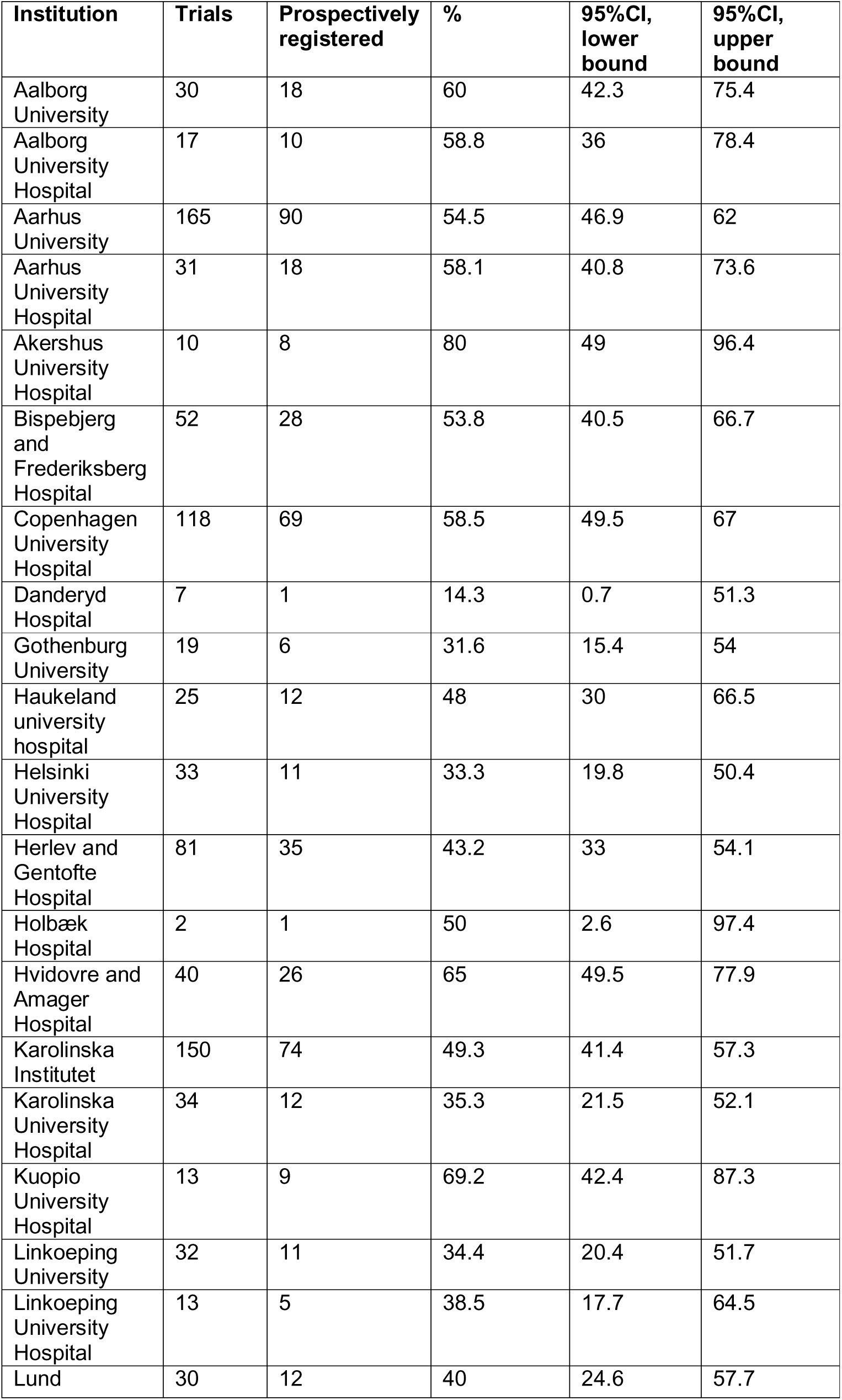
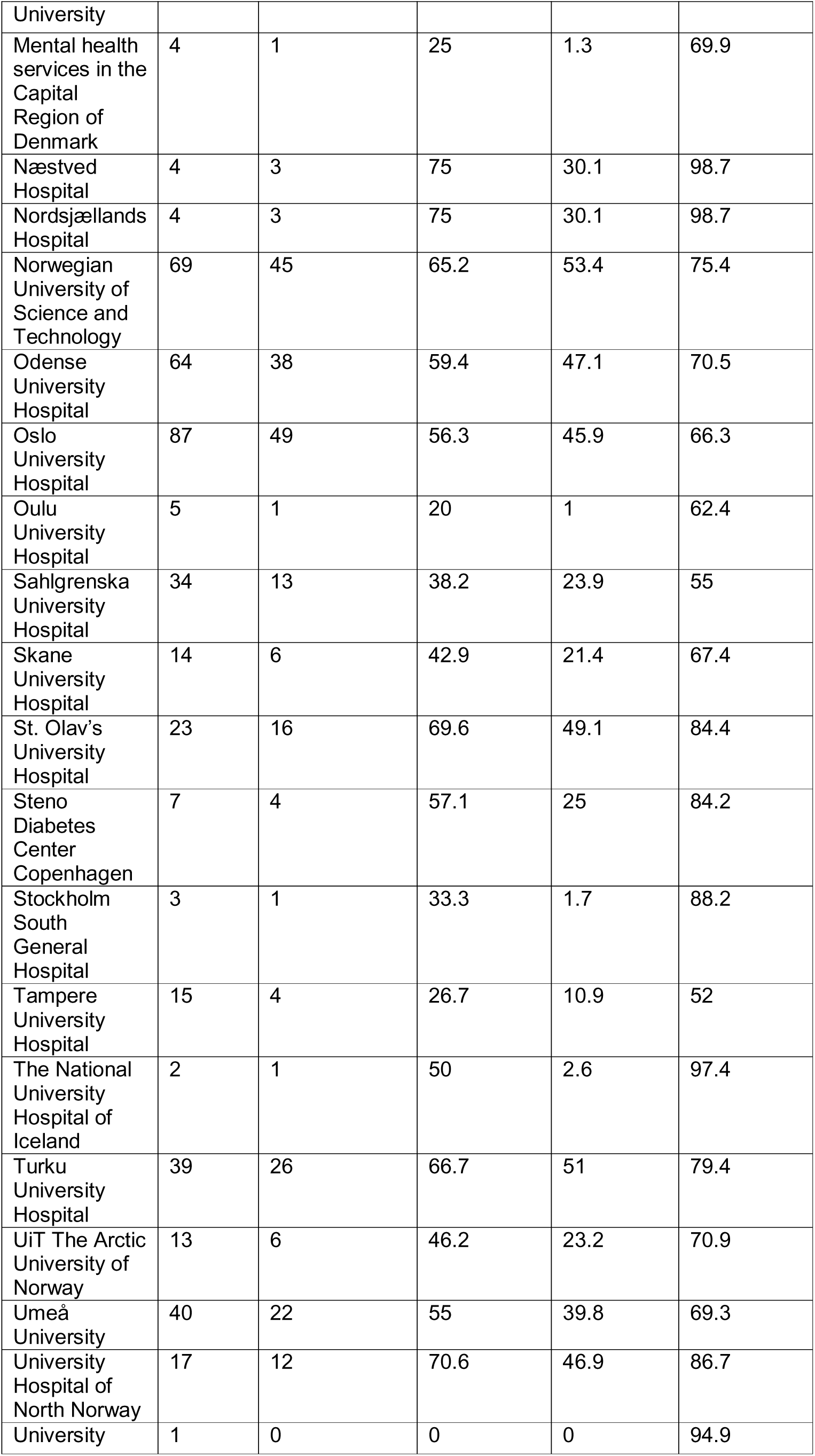
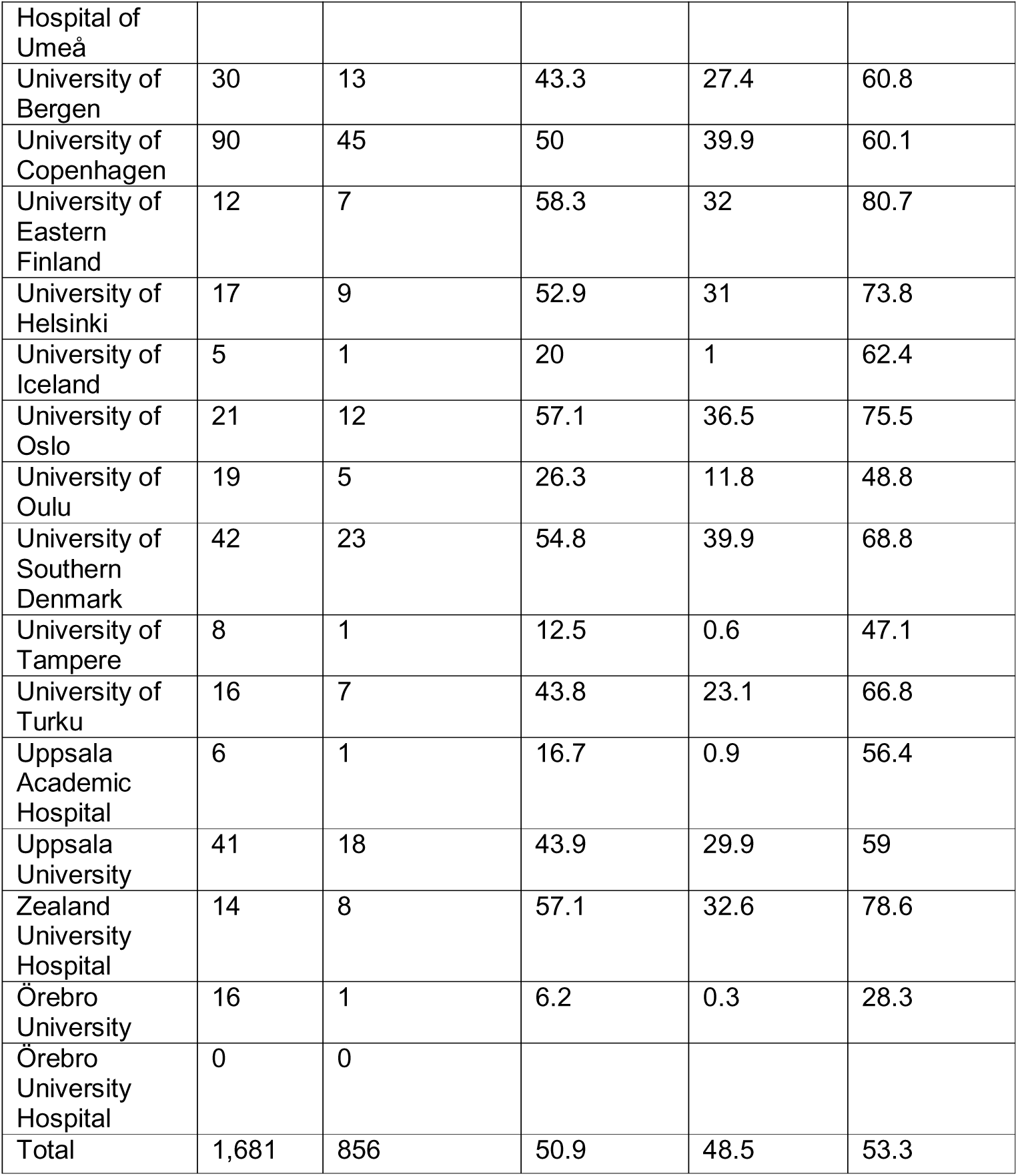
Share of trials with prospective trial registration among those registered at ClinicalTrials.govonly (n=1681), per institution.

**Table 9.**
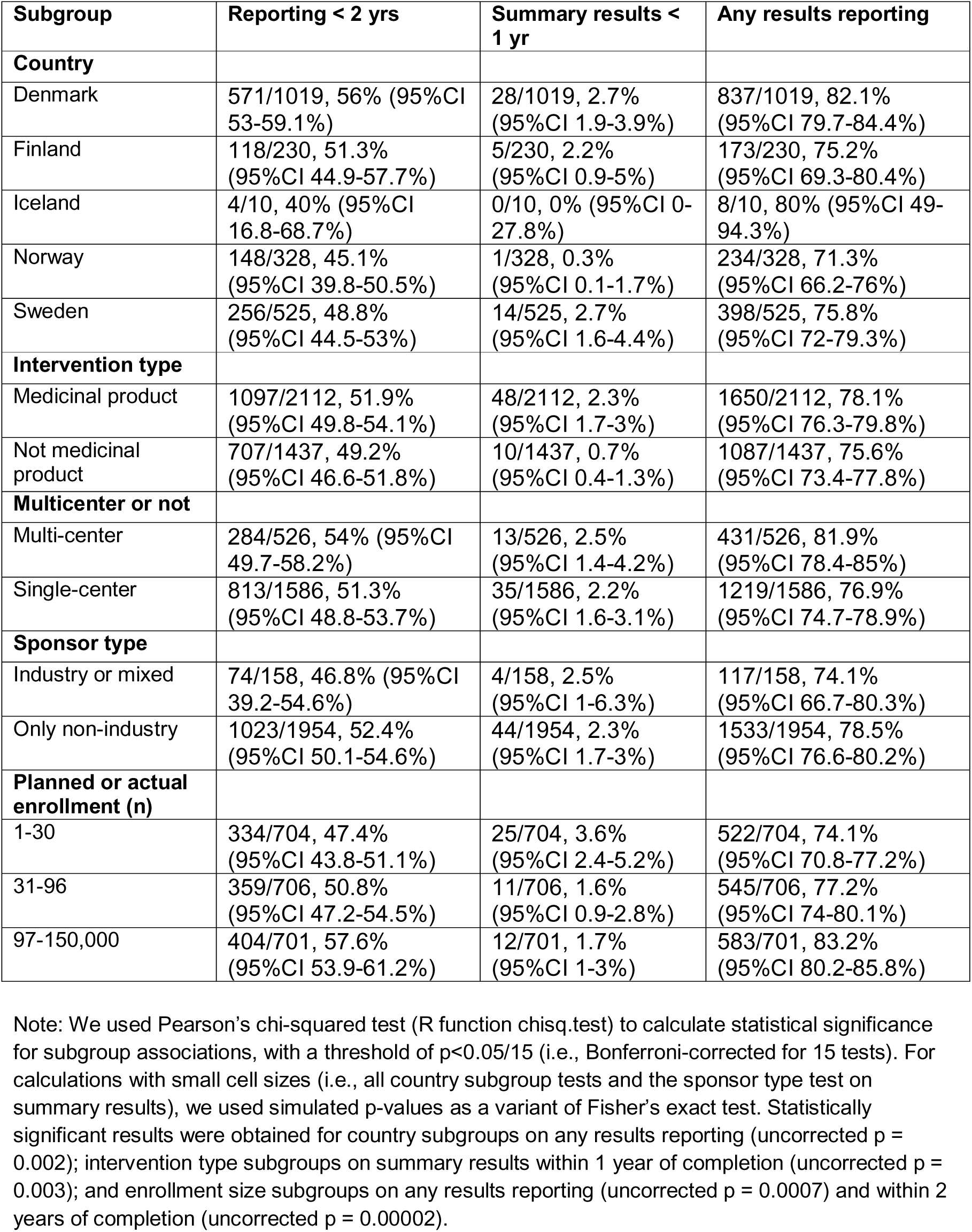
Subgroup analyses.

**Table 10.**
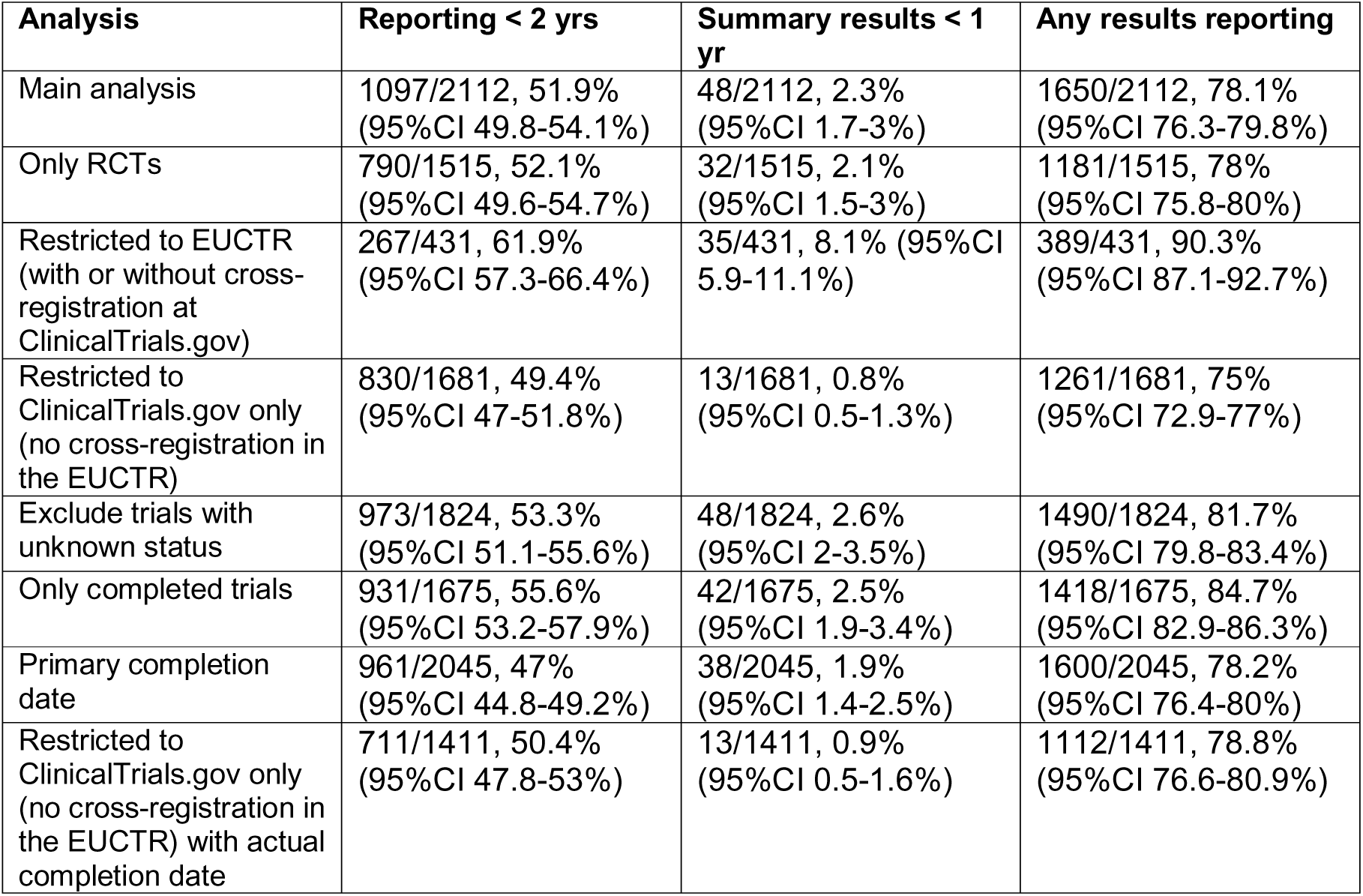
Sensitivity analyses.

**Table 11.**
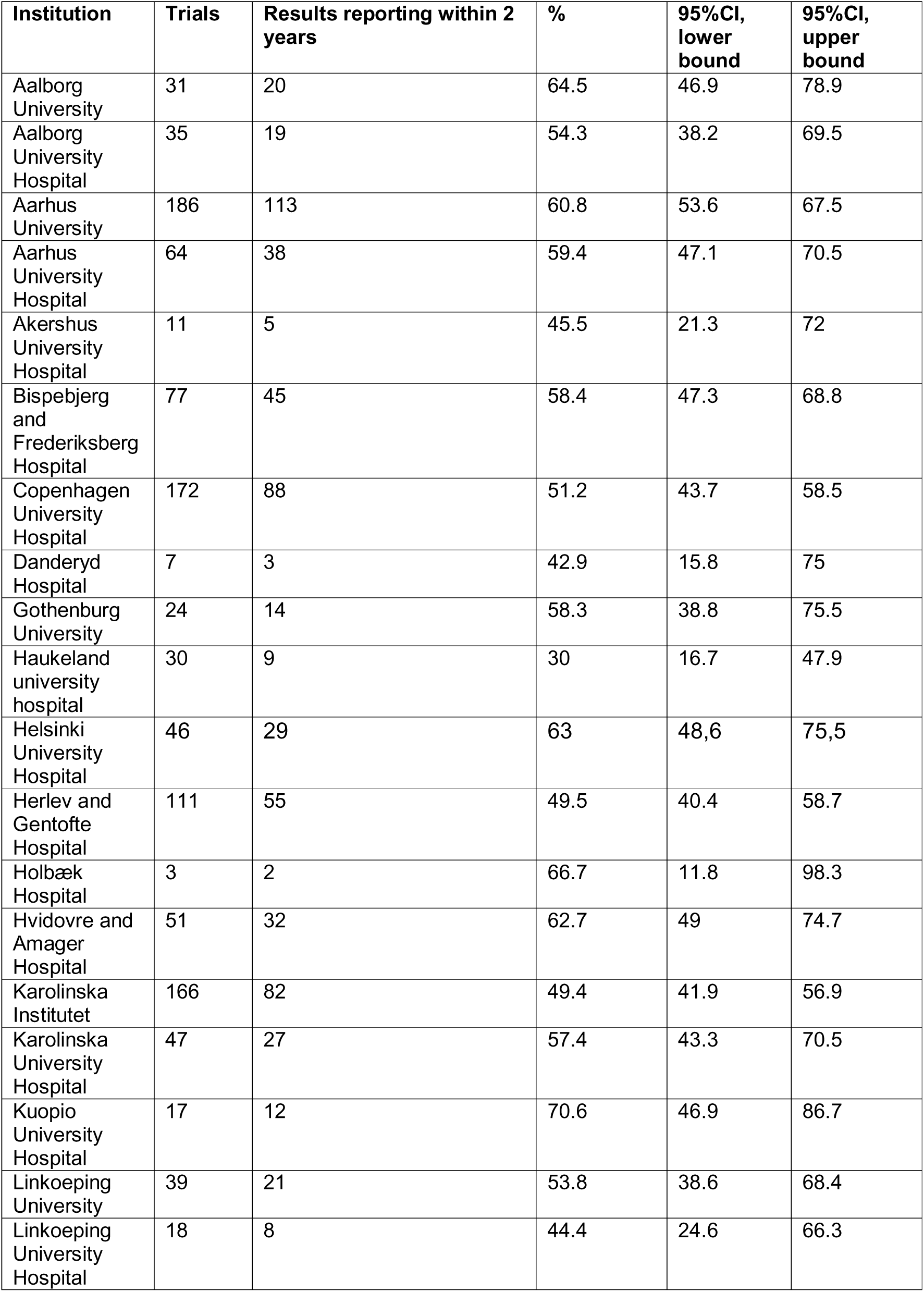
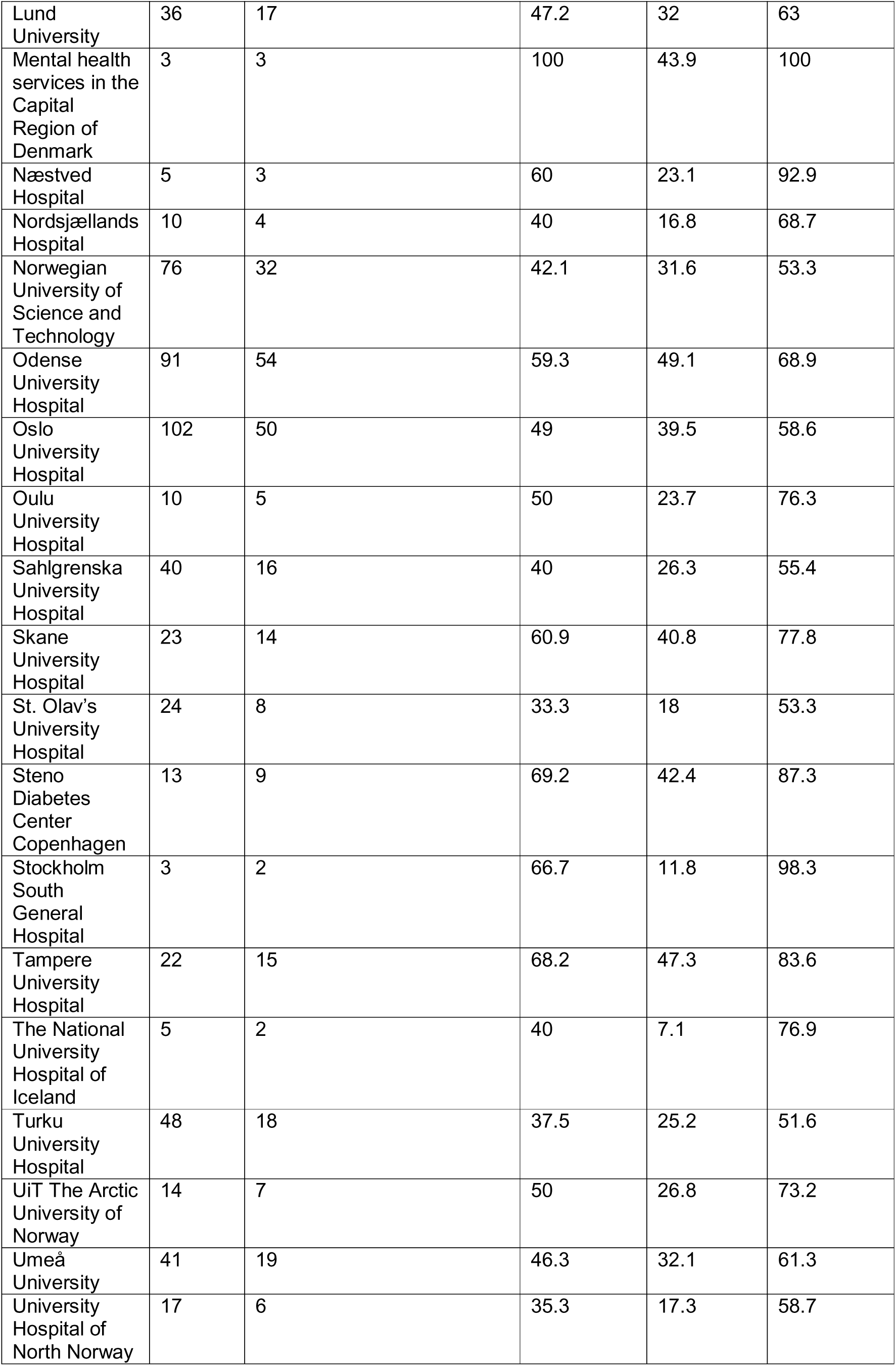
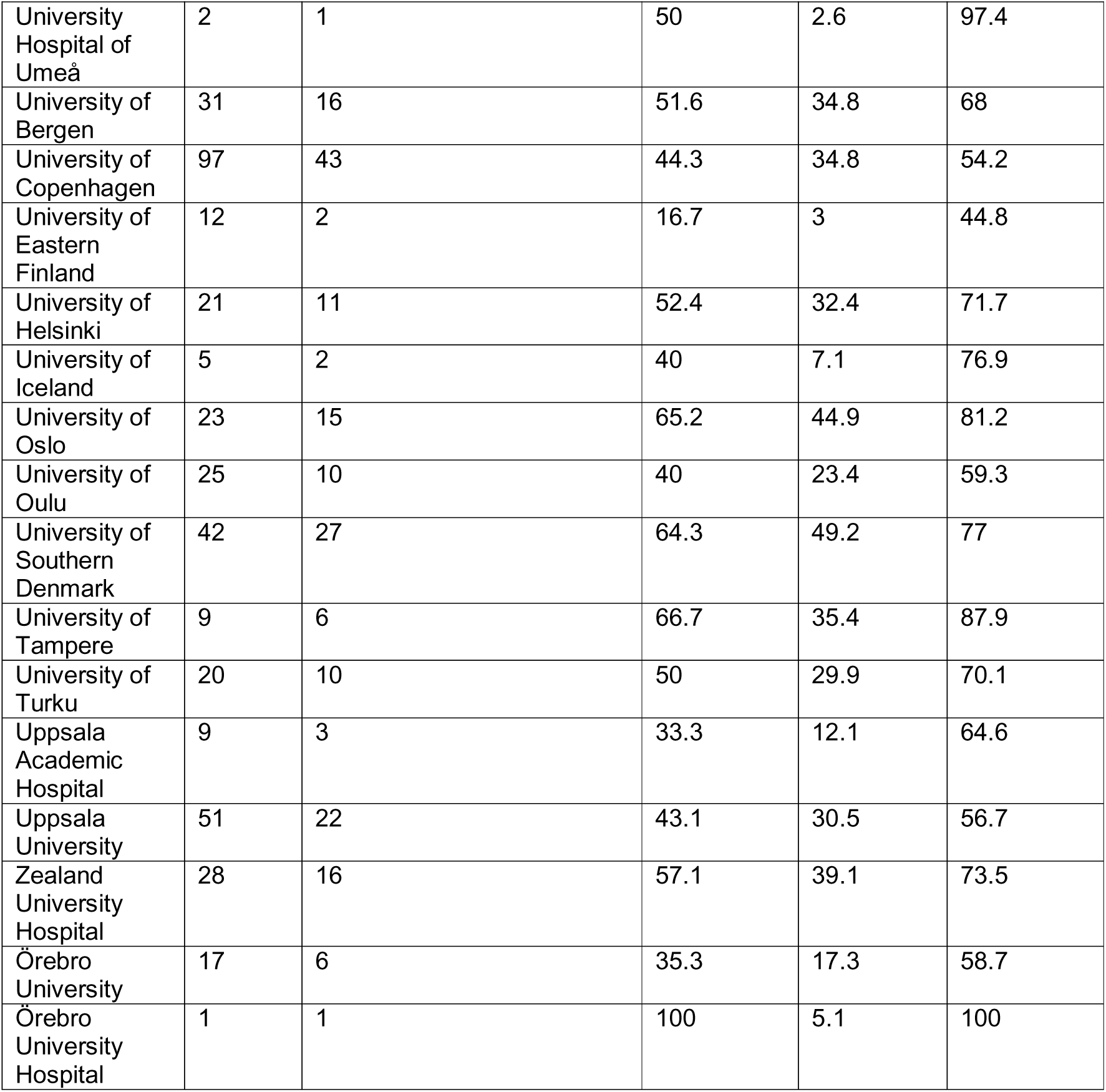
Sensitivity analysis: Only assigning trials to institutions that are the sole sponsor or first listed. Share of trials with summary results or results publication within two years of completion, per institution.

**Table 12.**
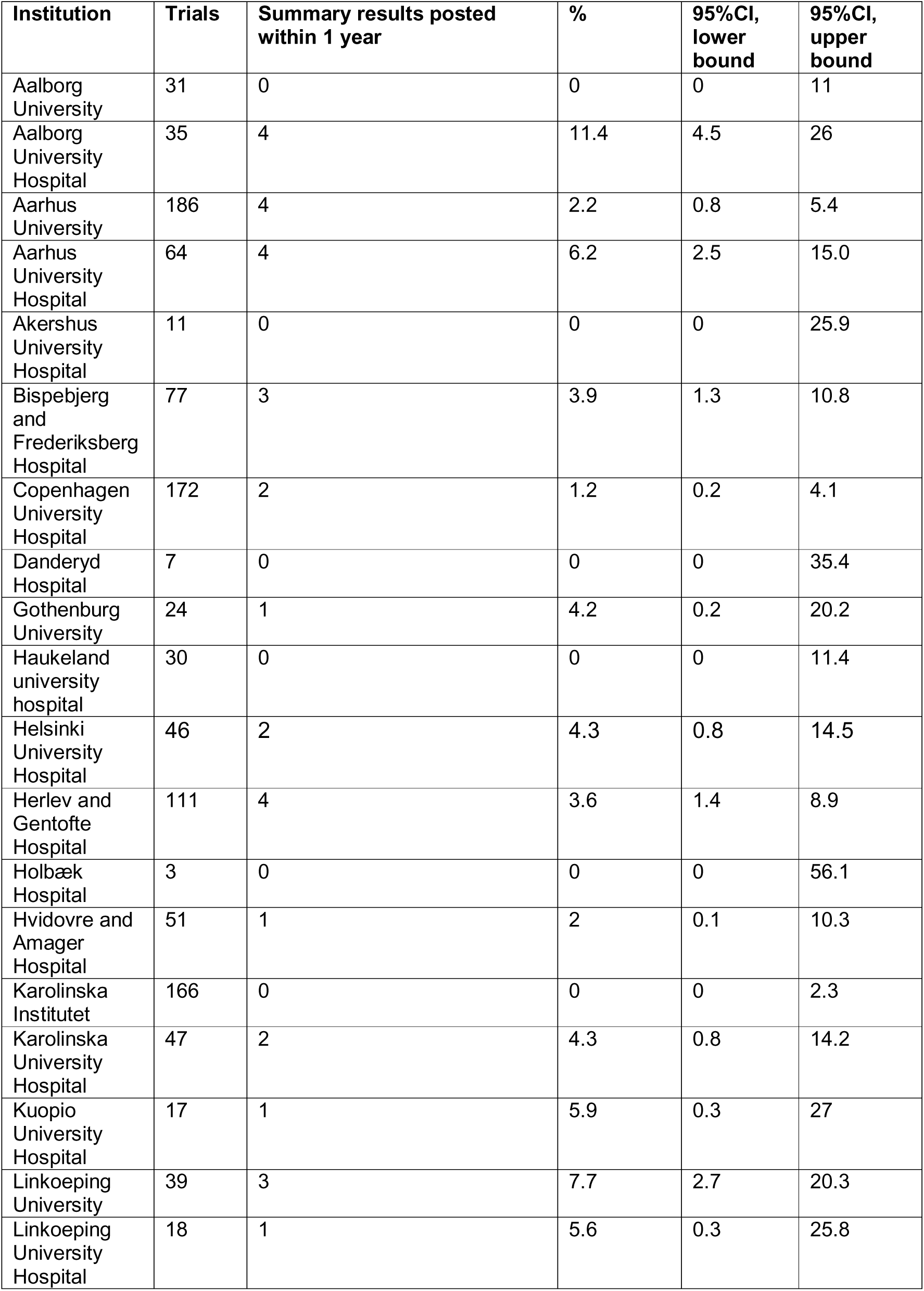
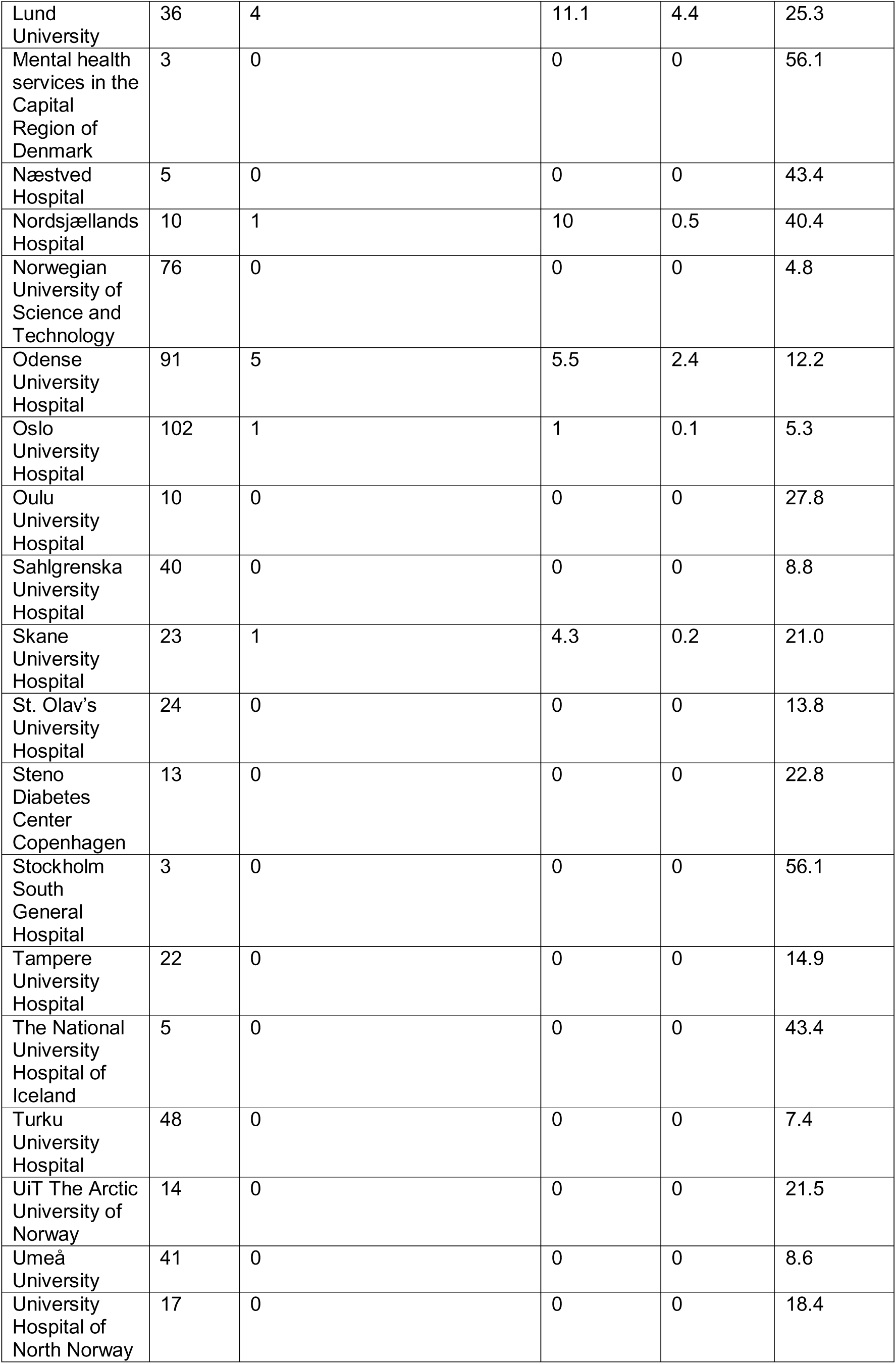
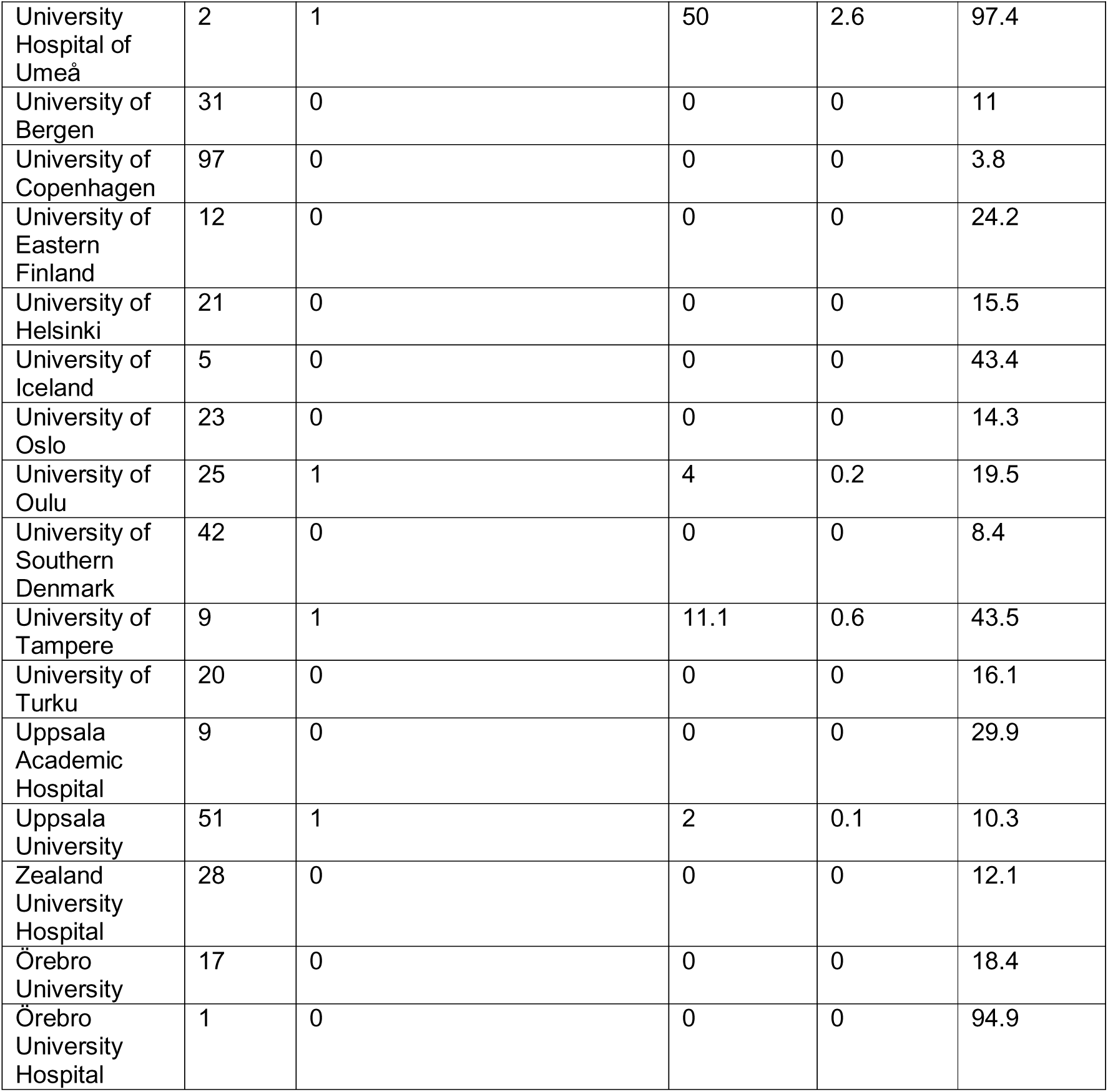
Sensitivity analysis: Only assigning trials to institutions that are the sole sponsor or first listed. Share of trials with summary results within one year of completion, per institution.

**Table 13.**
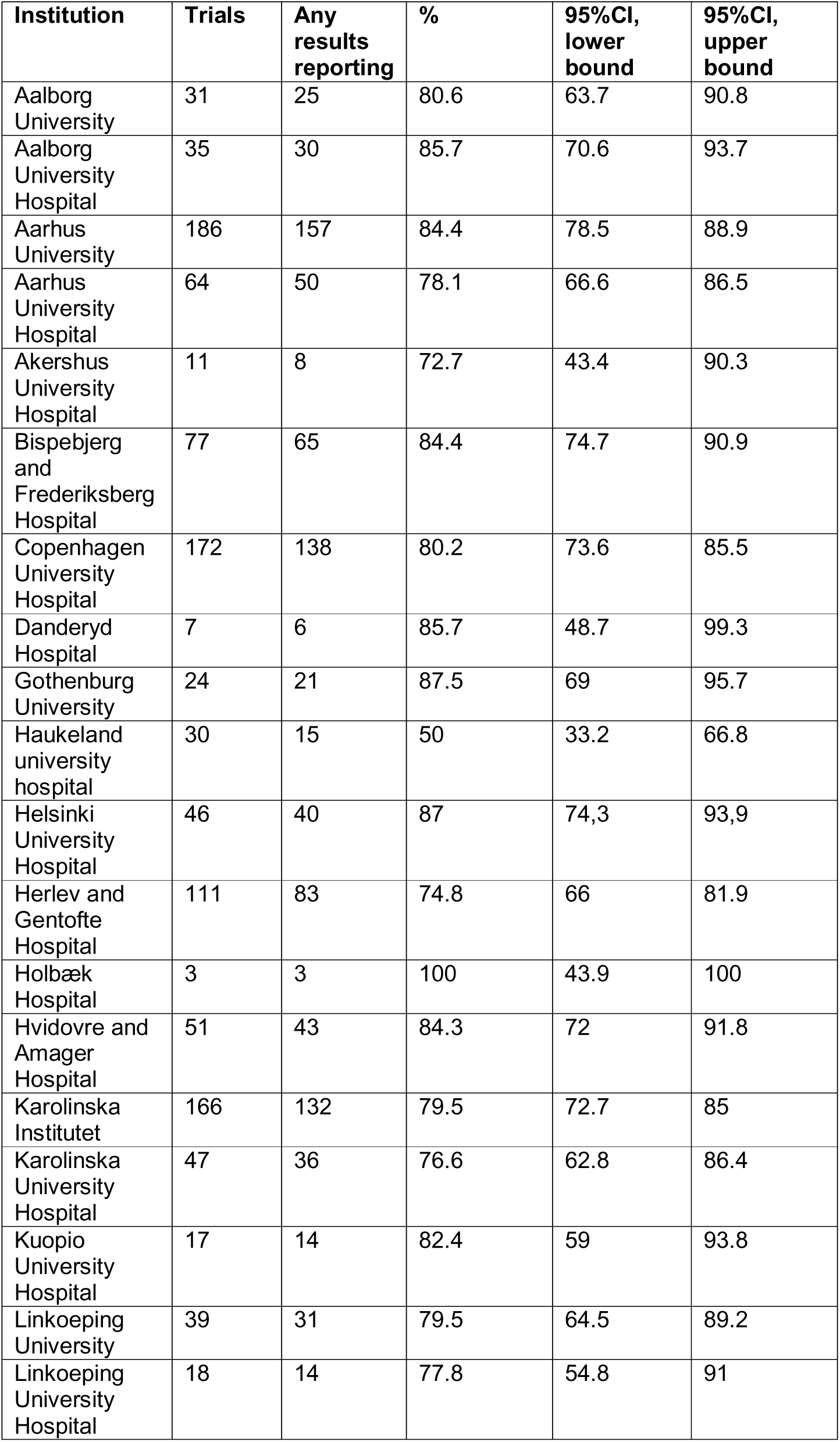
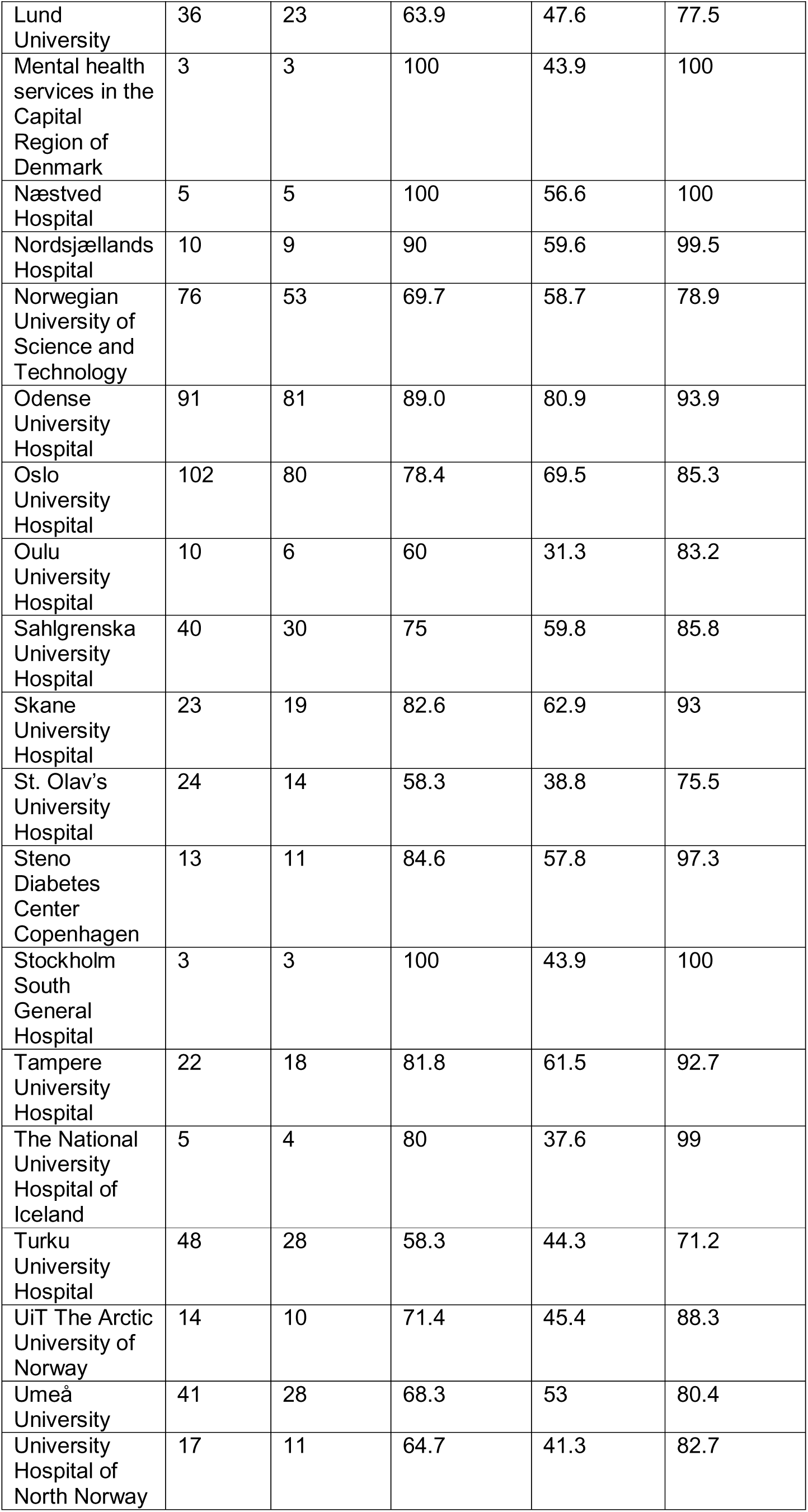
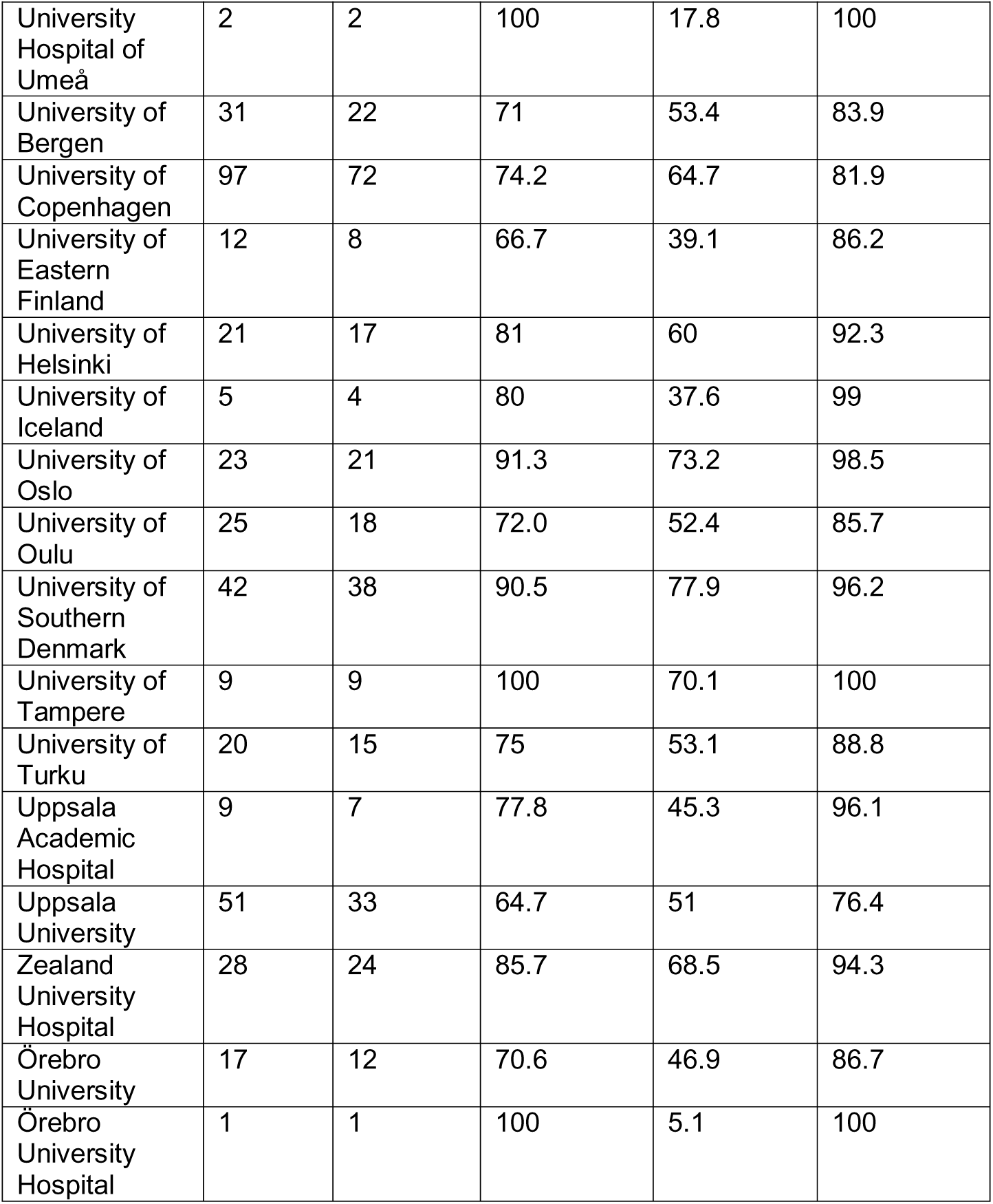
Sensitivity analysis: Only assigning trials to institutions that are the sole sponsor or first listed. Share of trials with any results reporting (summary results or results publications) at the end of follow-up, per institution.

